# A rapid review of interventions to reduce suicide ideation, attempts, and deaths at public locations

**DOI:** 10.1101/2025.04.09.25325515

**Authors:** Meg Kiseleva, Juliet Hounsome, Mala Mann, Abubakar Sha’aban, Riya Reji, Jacob Rees Davies, Rhiannon Tudor Edwards, Adrian Edwards, Alison Cooper, Ruth Lewis

**Author notes:** **Funding statement** The authors and their Institutions were funded for this work by the Health and Care Research Wales Evidence Centre, itself funded by Health and Care Research Wales on behalf of Welsh Government.

## Abstract

Suicide deaths are tragic events and those that occur in public places have an impact not only on the deceased person and their family and friends, but also on members of the public. Having up-to-date information about the effectiveness of interventions allows policymakers and organisations managing locations of concern to choose the most appropriate evidence-based suicide prevention strategies for specific locations. This rapid review was conducted to help inform the development of Welsh national guidance.

The review included literature published since 2014. 24 studies were identified, and these were conducted in the UK, Australia, South Korea, Canada, USA, Denmark and Japan. The studies covered railway or underground stations, bridges, cliffs or other natural heights, tall buildings, and other types of locations.

Surveillance technologies as a means of increasing opportunity for third-party intervention showed the most promise, although the evidence of their effectiveness was limited. In one study, having more closed-circuit television (CCTV) units was associated with fewer suicides at railway stations. Another study that tested a set of interventions including CCTV, infrared security fences, and a suicidal behaviour recognition and alert system, provided some promising initial descriptive data that showed an increase in the number of prevented suicides. Three other studies showed that there was no change in outcomes following the installation of interventions including surveillance technologies. Based on the assessment of the overall body of the evidence, there is a low level of confidence in the findings related to surveillance technologies because of the quality and designs of the studies.

Promotion of suicide helplines as an intervention aimed at increasing opportunities for help seeking was examined in seven studies. Two studies reported that the number of suicides increased after the introduction of the intervention. Three studies, of which two examined a set of interventions including helplines, observed no change. In two studies the effect could not be determined. There is a low level of confidence in the evidence for this outcome.

Other interventions evaluated included staff training; deployment of specialist staff; campaigns encouraging bystanders to intervene; a crisis café; blue lights at railway stations; suicide prevention messages, memorials, or notes other than official crisis line signage; spinning rollers at the top of fences that prevent gripping; and others. The effect of these interventions could not be determined with certainty but some of them appeared promising and warrant further research.

More robust evaluations are needed before any of the interventions reviewed here can be recommended for implementation. To create a better evidence base, high-quality evaluations should be supported and encouraged. Future research should examine which interventions work for who and in what circumstances.

## 1. BACKGROUND

### 1.1 Who is this review for?

This rapid review was conducted as part of the Health and Care Research Wales Evidence Centre Work Programme. The question was suggested by the Welsh National Programme Lead in suicide and self-harm prevention to help inform the development of the Welsh national guidance on how to respond to and manage public locations of concern for suicide. The findings of this review may be of interest to policymakers responsible for suicide prevention and organisations and individuals that manage public locations at risk for suicide.

### 1.2 Background and purpose of this review

It is suggested that in some places in England around a third of suicides happen in a public location (Public Health England 2015). The exact data for Wales is not available, but it is known that between April 2022 and March 2023, there were 356 deaths by suspected suicide of Welsh residents which could occur in or outside of Wales, equal to a rate of 12.6 per 100,000 people, and of these 8.1% happened in woods or forests, 2.2% at railway, underground, or tram stations, 2.2% at rivers, 2.0% at cliffs, 2.0% at sea, and 1.7% at bridges (Public Health Wales 2024). Suicide deaths are tragic events and those that occur in public places have an impact not only on the deceased person and their family and friends, but also on members of the public. They can be traumatic for bystanders who witness the death or discover the body, and especially so for people who inadvertently become directly involved, such as train drivers (Public Health England 2015).

Public locations in which many suicides happen are sometimes referred to as “locations of concern” or “frequently-used locations”. Public Health Scotland (2022a) defines a location of concern as a “specific, usually public, site that is used as a location for suicide and that provides either means or opportunity for suicide”. This can include bridges, tall buildings, car parks, roads, railway tracks, cliffs, woodland, rural, or secluded areas, or locations that provide access to water (Pirkis et al. 2015, Public Health Scotland 2022a). Locations of concern for suicide are sometimes referred to as “suicide hotspots”, but use of this term is discouraged due to it being regarded as sensational and trivialising the issue (Public Health England 2015, Samaritans 2024b).

Public Health England (2015) recommends four areas of action to prevent suicides at locations of concern: restricting access to the site and means of suicide (commonly referred to as physical means restriction), increasing opportunity and capacity for human intervention, increasing opportunities for help seeking by individuals at risk for suicide, and changing the public image of the site to dispel its reputation as a “suicide site”. It suggests a pre-emptive approach to potential locations of concern rather than a reactive one. In Scotland, the national guidance on action to address suicides at locations of concern was published in 2022. It identifies restricting physical means, enabling another party to intervene, signposting to sources of support, and managing the public image of locations as important actions to reduce suicides (Public Health Scotland 2022a).

Physical means restriction interventions have received the most attention in the literature, and they are considered the most effective. A number of systematic reviews of interventions aimed at preventing suicides in public locations published in recent years reported their effectiveness (Barker et al. 2017, Chamberlain & Woodnutt 2024, Havârneanu et al. 2015, Okolie et al. 2020b, Pirkis et al. 2015, Zalsman et al. 2016). There is much less certainty in the literature about the effectiveness of other types of interventions.

Installing physical barriers can be expensive and is not always possible, or practical, and there may be objections to how they change aesthetic characteristics of the location (Shin et al. 2024b). Having up-to-date information about the effectiveness of other intervention measures allows policymakers and organisations managing locations of concern to choose the most appropriate evidence-based suicide prevention strategies for specific locations.

This is what the present rapid review sets out to provide. It aims to answer the following question:

*What is the effectiveness of interventions other than physical means restriction to reduce suicide ideation, attempts, and deaths at public locations?*

## 2. RESULTS

We searched for international academic literature and supplemented it with grey literature from the UK. The methods used to conduct the review and eligibility criteria for selecting studies for inclusion are detailed in Section 5 of this report. A total of 24 studies reported in 29 documents were identified. The documents included both academic journal articles (n=19) and grey literature/pre-prints (n=10). The studies were conducted in the UK (n=8), Australia (n=7), South Korea (n=3), Canada (n=2), USA (n=2), Denmark (n=1), and Japan (n=1). Twenty studies used quantitative methods, two qualitative methods, and two mixed methods which included both a quantitative and qualitative element. Three of the included studies (Lockley et al. 2014, Ross et al. 2020, Torok et al. 2023) evaluated the same combination of interventions at the same location. Although these three studies were independent of each other, unless stated otherwise, we refer to them together and only count them once because they provide evidence on the same set of interventions, and may also be reporting on the same data or events. Reporting them together means that we avoid double-counting the evidence for this set of interventions.

The included studies covered multiple types of locations, including railway or underground stations (n=10), bridges (n=8), cliffs or other natural heights (n=3, covering the same location and set of interventions), tall buildings (n=1), and multiple types of locations with no breakdown of data by type of location (n=2). The interventions described in the studies were categorised as: interventions aimed at increasing opportunity for third-party intervention (n=12); interventions aimed at increasing opportunity for help seeking (n=8); physical means restriction interventions with an additional or innovative element (n=4); interventions aimed at creating a calming atmosphere (n=1); and suicide prevention messages and memorials excluding official crisis line signage (n=1). We also included studies that reported on interventions initiated by a bystander (n=3) to gather insight about what may happen if formal interventions aimed at encouraging bystanders to step in are successful. The same study could include multiple types of locations and interventions.

More information about the studies is available in Table 1 below. A detailed summary of each included study and the assessment of its methodological quality is provided in Section 6 of this report.

**Table 1.**
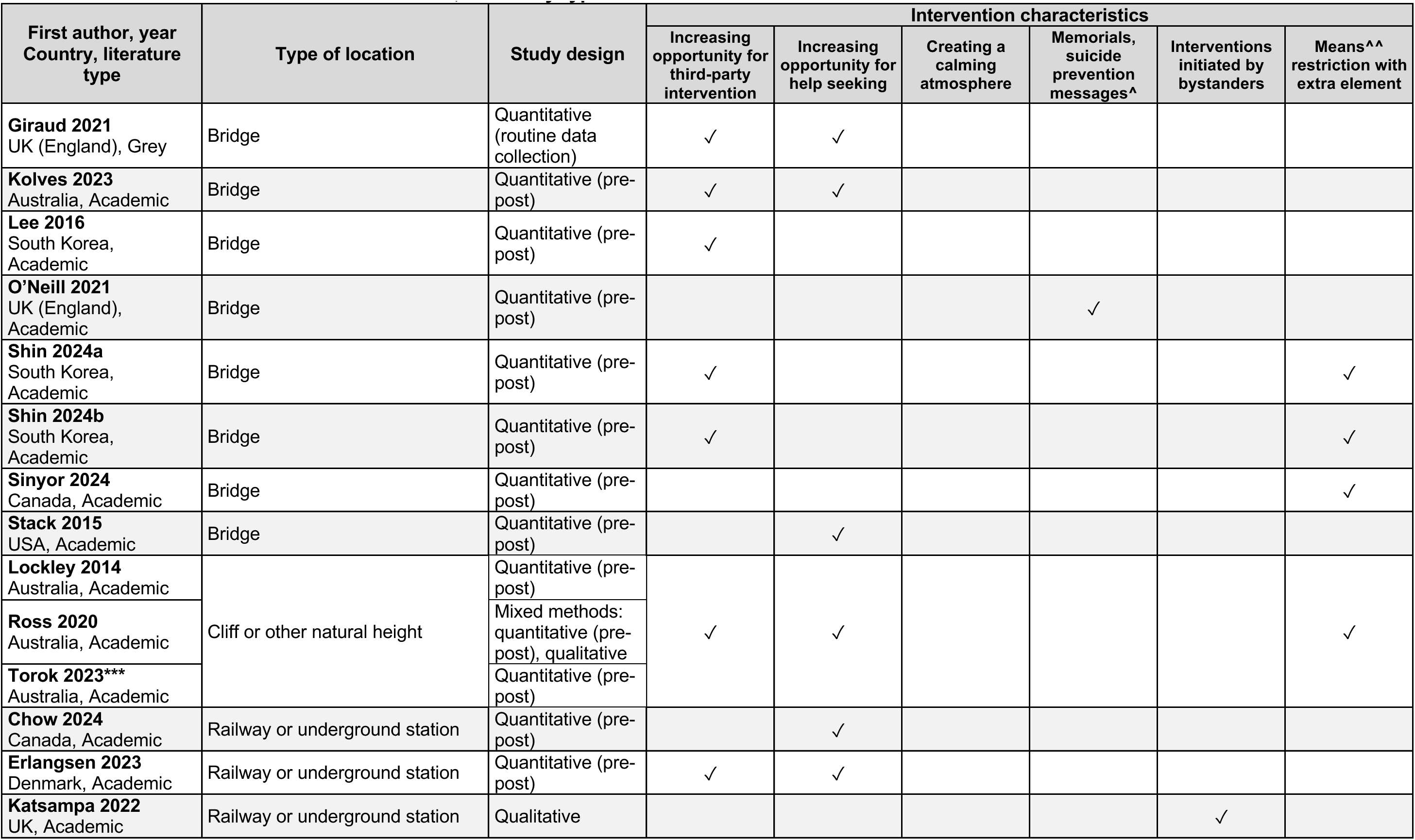
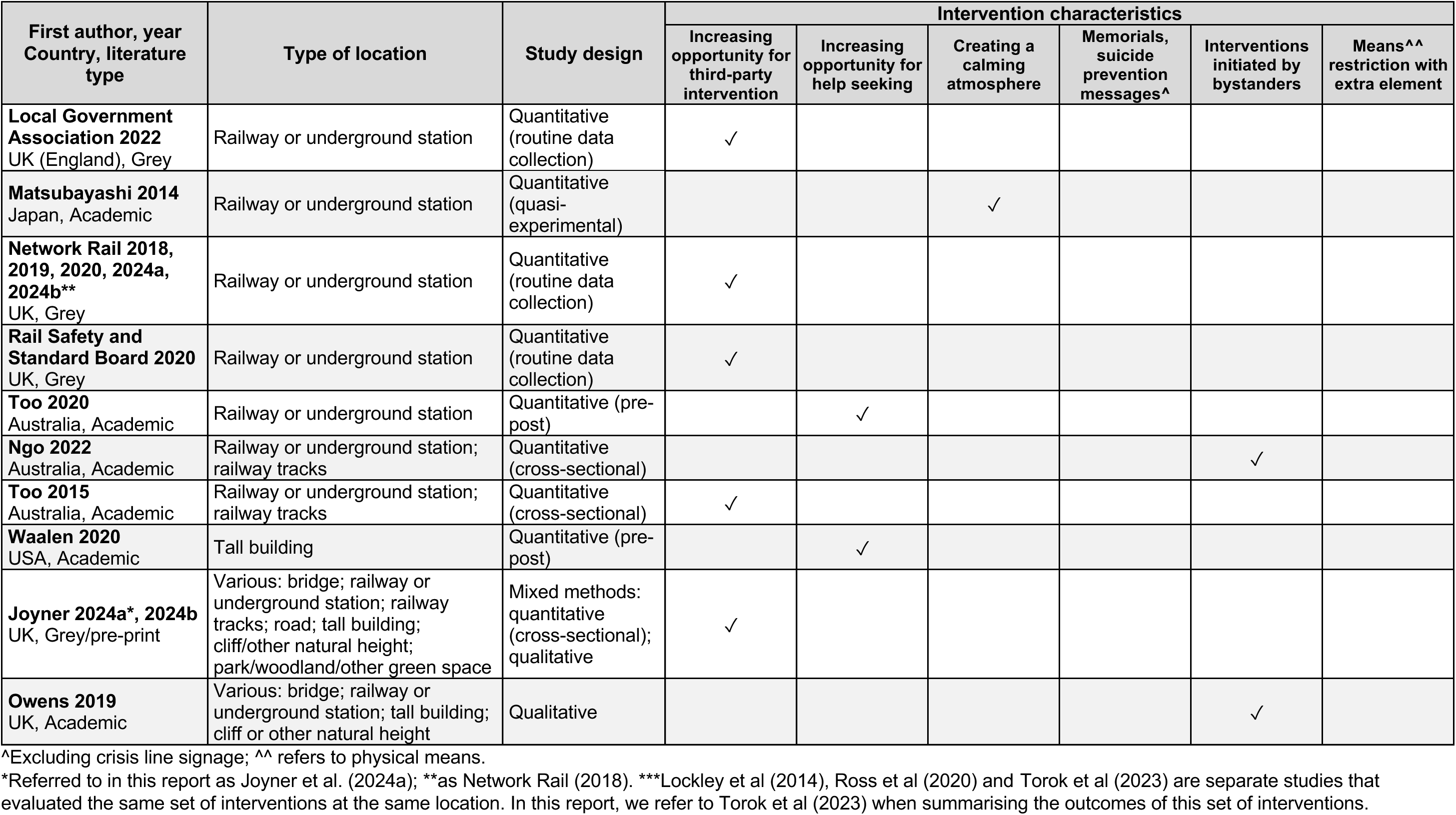
Characteristics of the included studies, sorted by type of location.

The results regarding the effectiveness of different interventions grouped by each category are presented separately for each type of location. An overall summary of the effectiveness of included interventions follows in a later section.

In Appendix 3, we also report a list of relevant systematic reviews identified during the literature searches which may be of interest to readers who want to further explore this area of research.

### 2.1 Interventions at railway stations and tracks

#### 2.1.1 Overview

Ten of the included studies described interventions implemented at railway stations, of which two (Ngo et al. 2022, Too et al. 2015) also covered those at railway tracks away from stations.

#### 2.1.2 Increasing opportunity for third-party intervention

This category includes a broad range of interventions, such as closed-circuit television (CCTV) or alert systems that signal to staff members the need to take action, or campaigns aimed at encouraging bystanders to step in. Five studies (Erlangsen et al. 2023, Local Government Association 2022, Network Rail 2018, Rail Safety and Standard Board 2020, Too et al. 2015) covered such interventions at railway stations, of which one (Too et al. 2015) also included railway tracks away from stations. Two of these five studies were published as peer-reviewed academic articles (Erlangsen et al. 2023, Too et al. 2015) and three as grey literature documents. Three studies (Local Government Association 2022, Network Rail 2018, Rail Safety and Standard Board 2020) concern the UK.

Too et al. (2015) analysed the relationship between the number of CCTV units at railway stations and car parks and the number of suicides on the railways within a broader study examining the links between railway suicides and neighbourhood-level social, economic, and physical factors. As part of this cross-sectional study, univariate analysis (which focuses on examining one factor without considering or adjusting for possible relationships with other factors) showed a slight positive association between the number of CCTV units (per 10 units) and risk of railway suicide (IRR=1.04, 95% CI 1.01–1.07, *p*=.009), i.e., it showed that having more CCTV units was associated with more suicides. However, in multivariate analysis (adjusting for other factors), this association was negative (IRR=0.93, 95% CI 0.88– 0.98, *p*=.004). This difference in the direction of effect, after controlling for other factors, may be explained by the high correlation between the number of CCTV units and the number of people using the station. The initial positive association suggests that stations with more CCTV units tended to have higher numbers of suicides, but after adjusting for other factors, the relationship became negative, likely because larger stations experienced more suicides due to their higher patronage. When this was accounted for, the data suggested that having more CCTV units was associated with fewer suicides. However, this was a correlational study and this design does not provide evidence of a causal relationship between the installation of CCTV units and reduced suicide rates.

Some descriptive data on interventions on UK railways was provided in grey literature documents. The UK’s Network Rail published data on suicides on the rail network and interventions to prevent suicide attempts. Data between 2016 and 2020 came from annual reports (Network Rail 2018, Network Rail 2019, Network Rail 2020) and data for 2023/24 was identified from statistics published on the Network Rail website (Network Rail 2024a, Network Rail 2024b). Interventions happening in this time period included training staff in suicide prevention techniques; the “Small Talk Saves Lives” campaign launched in November 2017, which was aimed at encouraging bystanders to support people who appear to be in emotional distress; and multiple other interventions such as the “Make a Connection” campaign starting in October 2023 that encourages people to access free mental health support and various awareness campaigns in partnership with Samaritans since 2010. There may also have been new physical means restriction interventions introduced in this time period. In 2016/17, there were 1,592 interventions by police, rail staff, and the public to prevent suicide attempts. This number rose to 1,711 in 2017/18, 2,270 in 2018/19 (22% of these were by rail staff and 9% by the public) and to over 2,000 in 2019/20, and was 1,937 in 2023/24. By 2020, more than 20,000 staff had been trained in making interventions to support those in emotional crisis. In 2016/17, there were 237 suicides on the rail network, and this rose to 246 in 2017/18, 271 in 2018/19, 283 in 2019/20, 276 in 2023/24. These data do not account for changes in population size and trends in the number of suicides more broadly, so it is not possible to infer from them whether there has been a change in suicide rates on the railways. The data on interventions to prevent suicide attempts published in the Network Rail reports suggest that a substantial number of suicides might have been averted by third parties, including bystanders, and that without such interventions the number of suicides on the rail network might have been higher.

The UK’s Rail Safety and Standard Board (2020) reported on an intervention that involved introducing Trespass & Welfare Officers deployed at 49 static high-risk locations and an additional 48 high-risk locations attended by five mobile teams. These officers, who had attended the Samaritans’ Managing Suicidal Contacts training course, provide support to individuals in distress and, when safe, make a physical intervention to avoid incidents. Between July 2019 and approximately one year later, Trespass & Welfare Officers carried out 130 crisis interventions, defined as “an immediate and short-term emergency responses to mental, emotional, physical, and behavioural distress”. Additionally, they conducted 20 physical interventions preventing individuals from trespassing on the railway and providing assistance, though it is unclear whether these involved people at risk of suicide. However, the report states that since the Trespass & Welfare Officers were introduced, there was a displacement of suicide-related incidents from station platforms to adjacent bridges.

The Local Government Association (2022) published data on the effectiveness of a crisis café in reducing suicidal ideation. In this intervention, station staff guided people in distress to a nearby crisis café where they could receive practical and emotional support from staff and peers, for example, with crisis resolution or building coping strategies, as well as information and advice, signposting, and referrals to health and social care providers, housing and community resources, and specialised mental health services. The railway station where the intervention was implemented also had British Transport Police representatives, Samaritans signage, and Samaritans awareness events, and 58% of its staff had attended the Samaritans Managing Suicidal Contacts training course. The document reported 264 visits to the café in two months. Recorded outcomes included 29 visits resulting in reduced suicidal ideation. Other outcomes included improved coping strategies (n=121), reduced social isolation (n=73), reduced self-harm (n=13), improved daily living skills (n=13), increased crisis management strategies (n=12), and averted statutory police interventions (n=2). It is difficult to make conclusions from these data because it is unknown how many of the visitors to the café needed support for each of these outcomes.

Additionally, the study by Erlangsen et al. (2023) included motion-sensitive lights at some parts of a railway station that could attract attention of staff or bystanders (although the authors explained their potential effect through limiting the appeal of dark spots), but the main focus of the study is on signposting to a suicide prevention helpline which was implemented at the same time, so it will be described in more detail in the next section.

#### 2.1.3 Increasing opportunity for help seeking

Three academic studies (Chow et al. 2024, Erlangsen et al. 2023, Too et al. 2020) reported on interventions at railway or underground stations aimed at increasing opportunity for help seeking. In all three studies, the intervention involved encouraging people to call helplines. In a Canadian study, subway platforms were equipped with posters and phones encouraging people to call a free suicide helpline, which connected callers with a trained counsellor (Chow et al. 2024). The counsellors assessed callers’ suicide risk and if it was deemed to be low, helped them with safety planning, and if it was considered high, liaised with transit control to slow or stop trains. In the 10-year period between 2011 and 2021, 243 calls to the crisis line were made, of which 72% were classified as low-risk, 16% as medium-risk, and 12% as high-risk. There was no significant decrease in suicides in the quarter following the implementation of the intervention (IRR=0.64, 95% CI 0.36–1.12, *p*=0.11), but after that, in each quarter an increase in suicides by around 2% was observed (IRR=1.02, 95% CI 1.00– 1.04, *p*=0.02). This study controlled for such factors as population size, the number of suicides by other methods in the same city, the unemployment rate, consumer price index, and others.

Posters and digital billboards with a crisis helpline number and a message encouraging help seeking, as well as digital billboards with a guided breathing exercise, were installed at Australian railway stations (Too et al. 2020). There was no significant difference in the rate of suicidal incidents pre- and post-intervention (IRR=0.88, 95% CI 0.59–1.30, *p*=.246). The only potential confounding factor that was accounted for was the number of station users, but there might have been other important confounders that were not controlled for. The number of crisis line calls during which suicide was identified as a safety issue did not change significantly (*p*=.169). However, the total number of crisis calls increased significantly from 154,521 before the intervention to 163,916 after (*p*<.001). The authors also surveyed station users and only 26% of the 1,844 respondents said they had noticed the campaign materials. This number ranged from 13% to 48% across stations.

In a study from Denmark (Erlangsen et al. 2023), signs encouraging help seeking with phone numbers for the national suicide prevention and emergency service as well as motion-sensitive lights were installed at a railway station. However, it was impossible to determine their effect because physical barriers were also installed at the end of some platforms during the same time period. In around 16 months since the interventions were implemented in December 2019, no new suicide deaths and one suicide attempt were recorded, compared to 11 deaths in the period between 2012 and 2018.

#### 2.1.4 Creating a calming atmosphere

A study from Japan examined the effect of installing blue lights at station platforms and tested whether it resulted in displacement of suicides to other stations (Matsubayashi et al. 2014). They compared suicides at stations with blue lights to neighbouring stations without blue lights. At the stations where the intervention was implemented, blue lights were installed at the edges of the platforms and sometimes also in the middle of the platforms, and were on from sunset to sunrise. At stations with blue lights, there was an average of 0.435 suicides per year before the intervention and 0.189 after. After accounting for station- and year-specific variables, such as the number of passengers, population size, types of platforms, and macroeconomic conditions, blue lights were associated with a reduction in suicides (IRR=0.258, 95% CI 0.127–0.523). No systematic evidence of displacement to nearby stations was found. However, when only the data from stations one stop away was considered, without adding stations further away to the model, there was evidence of a statistically significant increase in suicides at stations one stop away (*B*=0.718, *p*=0.01). Only data from one railway company was used and the possibility that people might go to a station managed by a different company was not considered, so it was impossible to establish whether installation of blue lights resulted in a reduction of suicides in the area, which was also served by other companies, or displacement to stations managed by other companies (Matsubayashi et al. 2014). This study followed preliminary communication by the authors published a year earlier (Matsubayashi et al. 2013), which attracted criticism from Ichikawa et al. (2014), who questioned the study’s methodology. Ichikawa et al. (2014) argued that it was necessary to control for the time of day at which suicides occurred, since the blue lights were only on at night, and the specific location at the station at which they occurred because the lights would only be visible in their immediate vicinity.

#### 2.1.5 Interventions initiated by bystanders

Two academic studies (Katsampa et al. 2022, Ngo et al. 2022) reported on interventions initiated by bystanders at railway stations or tracks. These are not formal interventions that policymakers or managers of locations of concern can implement, however, we have found that some formal interventions, such as “Small Talk Saves Lives” discussed in Section 2.1.2, aim at encouraging such behaviour. Therefore, we have included studies that addressed interventions initiated by bystanders to gather insight about what may happen if initiatives such as “Small Talk Saves Lives” are successful in encouraging bystanders to step in. In this section, “intervention” refers to an instance of a bystander taking action to prevent another person from suicide.

The UK study by Katsampa et al. (2022) examined the experiences of people intervening to prevent a suicide on the railways. The authors interviewed 21 people, of whom 11 were members of the public, including four with lived experience of suicidality and three mental health professionals, six train drivers, three railway employees, and one police negotiator. Most of the interviewees reflected positively on their experience and considered it the right thing to do, but some questioned if they should have behaved differently. A few interviewees struggled with not knowing what happened to the person they had helped afterwards and considered it an “unfinished story”. The experience of intervening in a suicide attempt was described as having made a lasting impact on some interviewees. Some of those who had previously witnessed a railway suicide described avoiding repeating such trauma as their main motivation for intervening.

Ngo et al. (2022) investigated the prevalence of preventative measures taken by bystanders in Australia at various railway locations, such as platforms, tracks, and level crossings. They also included interventions undertaken by railway staff and emergency services. During the time period between 2011 and 2019, at two heavy rail networks, there were a total of 635 interventions in suicide attempts, and of these, 139 were by bystanders. In 70 cases, bystanders acted as reporters, i.e. they alerted somebody else to the attempt, and in 69 they intervened as first responders. Of the 69 interventions, 77% involved physical interaction and 49% involved more than one bystander.

#### 2.1.6 Bottom line results for interventions at railway stations and tracks

Ten studies evaluating or describing different types of interventions at railway stations and tracks have been identified. Only installing CCTV cameras showed some evidence of effectiveness. In one study (Too et al. 2015), a larger number of CCTV units at railway stations and car parks was associated with fewer suicides. It was a cross-sectional study that was generally well conducted.

Network Rail (2018) reported that a large number of suicide attempts were prevented by railway staff, emergency services, and members of the public during the period when staff were being trained in helping people in emotional distress and a public campaign encouraging bystanders to step in if they saw a person who appeared to be in crisis. However, there was no evidence of direct links between these interventions and prevented suicides, and no data on changes in suicide rates was available. Some suicides were prevented following the introduction of specialist Trespass & Welfare Officers, but there was some evidence that suicides might have been displaced from platforms to nearby bridges (Rail Safety and Standard Board 2020). Some data suggests that a crisis café adjacent to a railway station helped some people deal with suicidal ideation, but it is unclear if the café had any effect on suicide attempts and deaths at the station (Local Government Association 2022). These three studies only provided descriptive statistics, and no formal evaluation of any of the interventions described in them was available.

Three studies evaluating interventions aimed at increasing opportunity for help seeking were identified. All of them involved encouraging people in crisis to call helplines, either by installing phones with a link to a crisis line or signage with a phone number at stations. There was no evidence that this type of intervention was effective in reducing suicides. In the study by Chow et al. (2024), the number of suicides increased after the start of the intervention and in the study by Too et al. (2020) there was no difference in before and after the intervention. No relationship between the intervention and suicide rates could be established in the remaining study (Erlangsen et al. 2023) because physical barriers were installed at the station at the same time as the signage. These studies varied considerably in their quality. The Chow et al. (2024) study was considered to be well-conducted, but the remaining two (Erlangsen et al. 2023, Too et al. 2020) had many limitations, such as the interventions not being described in sufficient detail and having insufficient data to provide confidence in the findings. In the study by Erlangsen et al. (2023), no statistical analysis was conducted. In Too et al. (2020), it was unclear how the outcomes were determined and whether they were determined consistently.

The study that evaluated installation of blue lights at platforms showed that they were effective in preventing suicides but a number of methodological issues, discussed in detail in Section 2.1.4, limit its validity (Matsubayashi et al. 2014).

Finally, two studies described interventions initiated by bystanders. The data presented in the study by Ngo et al. (2022) suggested that bystander interventions might have prevented a considerable number of suicides at railway stations. This study was generally performed well in terms of the outcomes of interest to this review. The other study (Katsampa et al. 2022) examined bystanders’ experience of intervening. Some of the participants described the situation having had a lasting impact on them. This qualitative study did not reflect on the potential influence of the researchers on the research, and it was unclear how the researchers were situated culturally or theoretically, but otherwise it was conducted well.

### 2.2 Interventions at bridges

#### 2.2.1 Overview

Eight studies described interventions conducted at bridges. The bridges could be over water, roads, or railway tracks.

#### 2.2.2 Increasing opportunity for third-party intervention

Increasing opportunity for third-party intervention to prevent suicides at bridges was examined in five studies (Giraud 2021, Kolves et al. 2023, Lee et al. 2016, Shin et al. 2024a, Shin et al. 2024b), of which four were published as academic articles and one as a grey literature document (Giraud 2021). Three studies were about locations in South Korea, one in the UK, and one in Australia.

All three of the South Korean studies examined detection and alert systems. The study by Lee et al. (2016) described an intervention whereby security fences with infrared sensors and pole camera surveillance systems were placed on a bridge, and an intelligent safety control system that monitored people’s moving trajectory, time spent in different sections of the bridge, and their behavioural patterns by using digital CCTV with an automatic suicidal behaviour recognition and alert system. This intervention was implemented on two bridges with the highest number of drowning incidents. During the 12-month trial period, a total of 101 people were stopped from suicide attempts on the two bridges, of whom 92.1% were rescued on the bridge and 7.9% in the water. Pre-post data was only available for one of the bridges. In the year before the intervention, 15 suicide attempts were prevented, compared with 93 in the year during which the intervention was trialled.

The study by Shin et al. (2024a) described the effectiveness of installing a one-metre fence over an existing 1.5-metre railing with five tension wire sensors that alerted a rescue team if a wire is cut or pulled by more than 10 centimetres. The fence also had abacus-bead-shaped spinning rails on the top that prevented people from gripping it and climbing over. We also classified this set of interventions as physical means restriction with an additional or innovative element. The bridge where it was implemented already had fixed phone boxes with direct access to a crisis line, CCTV, and signage with supportive messages. After the intervention, there was a statistically significant reduction in the number of suicides on the bridge (IRR=0.37, 95% CI 0.26–0.54), with an average of 25.5 suicides per year pre-intervention and 9.5 per year post-intervention. The study does not specify how many suicides were prevented by the rescue team’s arrival versus those prevented by the spinning rails at the top of the fence. Besides, the vertical extension to the fence would have made it more difficult to climb. Therefore, it was not possible to attribute the decrease to any specific intervention component.

Another study by Shin et al. (2024b) described a different bridge in South Korea where a Video Incident Detection System (VIDS) was installed, comprised of 14-speed sensors that warned the operation control team if the speed of a car was below 30 km/h. Meter-high spinning bars were also added to the top of the existing meter-high guard rails on both sides of the bridge three years later, so data is available pre-intervention, after the installation of the VIDS, and after the installation of the VIDS and spinning bars. For the purpose of this review, the spinning bars were classified as physical means restriction with an additional or innovative element, and are therefore also listed under Section 2.2.5. Prior to the described intervention, the bridge already had a 1-metre-high rail and CCTV. A total of 146 incidents, including both suicide deaths and prevented suicides, occurred on the bridge during the 14-year and 1-month study period. Of these, 54 incidents took place during the 6.5-year pre-intervention period, 58 incidents during the 2 years and 11 months of the VIDS-only phase, and 34 incidents during the 4 years and 8 months of the VIDS and spinning bars phase. Pre-intervention, there were 20 deaths by suicide on the bridge, equivalent to 0.008 deaths per day, compared to 11 (0.010 per day) in the VIDS-only phase and fewer than five (0.002 per day) in the VIDS and spinning bars phase. Suicide deaths increased non-statistically significantly after the installation of the VIDS compared to the pre-intervention period (IRR=1.23, 95% CI 0.59–2.56) but decreased after the installation of the spinning bars compared to the VIDS-only period (IRR=0.23, 95% CI 0.07–0.71) as well as to the pre-intervention period (IRR=0.28, 95% CI 0.10–0.82).

The study also provides details about the number of interventions to stop a suicide attempt during the three periods (Shin et al. 2024b). There were 33 such interventions, equal to an average of 0.023 per day, in the pre-intervention period, 46 (0.054 per day) in the VIDS-only period, and 29 (0.021 per day) in the VIDS and spinning bars period. The number increased statistically significantly in the VIDS-only period compared to before the intervention (IRR=2.40, 95% CI 1.65–3.47), but then decreased statistically significantly in the VIDS and spinning bars phase vs. VIDS-only (IRR=0.37, 95% CI 0.25–0.57). The difference between the VIDS and spinning bars phase vs. pre-intervention was not significant (IRR=0.90, 95% CI 0.59–1.38). Before the intervention, 61.1% of incidents were intervened in, compared to 79.3% in the VIDS-only phase and 85.3% in the VIDS and spinning bars phase.

The effectiveness of installing crisis line phones and CCTV cameras was examined in an Australian study by Kolves et al. (2023). This study reviewed data for the period between 2001 and 2021. The phones and CCTV were installed in 2012, and in 2015 physical means restriction barriers were added. The data were analysed over three-year periods. As well as suicides on the bridge, the authors examined displacement of suicides to other locations. The number of suicides on the bridge did not change between the three years before the installation of the phones and CCTV (n=21) and after (n=21). However, it started to rapidly decline after the installation of the barriers, with each of the two following three-year periods having fewer than five suicides. Clustering the numbers of suicides in three-year periods does not show that the phones and CCTVs had effectiveness, but the authors report that a joinpoint regression analysis identified 2012 – the year of their installation – as the start of the decline in the number of suicides. Over the study period, two join points were identified, i.e., points in time when a statistically significant change in trend happened. The number of suicides decreased between 2001 and 2009, with the annual percentage change (APC) being −26.7% (95% CI −43.4−-5.1%, *p*=0.02). Following that, there was a rapid increase in suicides until 2012 and a period of decline between 2012 and 2021 (APC=-31.6%, 95% CI − 44.9−-15.1%, *p*=0.002). Regarding displacement to other locations, there was no substantial difference in the number of suicides at other bridges and cliffs in the city (0 join points; APC=0.6%, 95% CI −3.0−4.4%) and the difference in suicides in the suburbs bordering the bridge was not statistically significant (0 join points; APC=2.8%, 95% CI −0.1−5.9%). However, there were some fluctuations in suicides at man-made constructions in the inner city, including after the installation of the phones and CCTV on the bridge, although the upward trend was less pronounced after 2012 (3 join points; 2007−2012 APC=36.8%, 95% CI −0.2−87.5%; 2012−2021 APC=4.4%, 95% CI 4.0−13.5%). There is some confusion about the reporting of the latter confidence interval for this outcome because even though it does not cross 0, the authors of the study report that the trend was not statistically significant. We suspect that a minus sign might be missing.

A UK grey literature document by Giraud (2021), prepared as a presentation to the National Suicide Prevention Annual Conference “Suicide Prevention in the Square Mile”, reported on a number of interventions aimed at increasing opportunity for third-party intervention and well as increasing opportunity for help seeking to prevent suicides on bridges. The interventions included Samaritans signs on three bridges, training sessions to the public and frontline staff at various locations around the city, leaflets handed out to pedestrians at a bridge, mental health nurses accompanying police officers who respond to incidents, training of business staff along the river, promotion of water safety and suicide awareness materials to licensed premises along the river, and leaflets about suicide prevention distributed at transport hubs and on the bridges. No formal evaluation was identified, but Giraud (2021) reported that since the introduction of the intervention package, reattendance at the bridges was reduced to zero. No other outcomes were reported. We identified an evaluability assessment of this suicide prevention initiative conducted by the National Institute for Health and Care Research Public Health Intervention Responsive Studies Teams (2024) during the literature searches for this review, so it is possible that an evaluation will be conducted in the future.

#### 2.2.3 Increasing opportunity for help seeking

Three studies presented interventions aimed at increasing opportunity for help seeking at bridges, of which two (Giraud 2021, Kolves et al. 2023) were already described in Section 2.2.2 of this report. The remaining study (Stack 2015), from the USA, evaluated the effectiveness of installing phones with a direct link to a crisis centre on a traffic bridge across water without pedestrian walkways. Six such phones were installed. The study examined suicide numbers in the 13 years after the intervention compared to the 13 years before. Controlling for the suicide rate in the state, the number of suicides increased by an average of 2.73 (*SE*=1.57) suicides per year (*R^2^*=.418, *p*<.05). At the same time, the city population decreased by 1.4% during the decade that falls within the 13-year period since the implementation of the intervention. During the first 10 years of the crisis phones being operational, 27 people used them. Potentially confounding factors not controlled for in the study include a website dedicated to suicides on the bridge that originated around the same time as the crisis phone and a local newspaper’s policy of publishing articles on all suicides on the bridge. These two factors might have contributed to the image of the bridge as a location for suicides and interfered with effectiveness of the intervention.

#### 2.2.4 Memorials or suicide prevention messages other than crisis line signage

A UK study (O’Neill et al. 2021) examined the effects of placing memorials, messages, or notes (excluding official crisis line signage) on motorway bridges to deter people from suicide as well as media coverage of such “decorations”, as they are referred to in the study. Across the 26 bridges, in the period of one year, there were 160 suicides. Of these, 93 occurred pre-decoration, and 67 were post-decoration (56 with no media coverage and 11 with media coverage concerning the same bridge). This difference was not statistically significant (*p*=0.55). In terms of individual bridge-level data, 15 bridges had more incidents pre-decoration than post-decoration, but this difference was not statistically significant (Bonferroni corrected *p*>.05). Eleven bridges had more incidents post-decoration (*p*-value not reported), of which one had more incidents post-decoration with media reporting, but that was not statistically significant either (Bonferroni corrected *p*>.05).

#### 2.2.5 Physical means restriction with an additional or innovative element

We have already described the installation of the spinning rollers on fences, which was done in combination with another intervention (Shin et al. 2024a, Shin et al. 2024b), in Section 2.2.2 of this review. One more study described a physical means restriction intervention with an additional element on a bridge (Sinyor et al. 2024). This study described a 5-meter barrier with lights that reacted to the wind and followed pre-programmed routines at sundown, sunrise, and midnight, and whose colours depended on the season. The barrier was installed in 2003, but according to the website of a local news publication, the lights were only added in 2015 (Kupferman 2015). The study does not provide a breakdown of data before and after the lights were installed, so it is not possible to establish if they had an effect on the suicide. However, after the installation of the barrier and until the end of 2020, only two suicides occurred on the bridge, compared to 48 in the five years before, so it would not have been possible to analyse more fine-grained data. The study also analysed potential displacement to other large metropolitan cities in the same province and found no evidence of this (City 1 IRR=0.50, 95% CI 0.26–1.01, City 2 IRR=1.17, 95% CI 0.44–3.43). However, it is not necessarily reasonable to assume that preventing access to a location will cause people to travel to a different city. No data on displacement to nearby locations within the same city was available. Regarding method substitution determined by the number of suicides by methods other than jumping from bridges, Sinyor et al. (2024) found no change either (IRR=1.00, 95% CI 0.99–1.01).

#### 2.2.6 Bottom line results for interventions at bridges

Of the interventions described in this section, physical means restriction interventions with an additional or innovative element, sometimes in combination with a technological intervention aimed at increasing opportunity for third party intervention, appeared to have the most promise. In one study (Shin et al. 2024a), suicides on a bridge decreased significantly after a fence with wire sensors that alerted a rescue team and with spinning bars that prevented people from gripping it, however, because of a vertical extension to the fence installed at the same time, it was not possible to determine whether the decrease was due to the innovative features or to the physical means restriction. In another study (Shin et al. 2024b), installing a video detection system did not reduce suicide deaths on the bridge, but adding a vertical extension with spinning bars to the guard rails did. Again, it was not possible to determine if the reduction in suicides was due to the innovative feature or to the extension to the fence. The bridge in the study by Shin et al. (2024b) had no pedestrian access, so it is not clear whether the findings of this study are generalisable to those that do. It was unclear if the bridge in Shin et al. (2024a) had any unique features that might limit the generalisability of the findings. Also, the volume of data in each of the compared periods in Shin et al. (2024b) was not large, potentially limiting how much confidence can be placed in the results of the analysis. Another South Korean study (Lee et al. 2016) examined a combination of technological interventions that included an alert system and installing fences with infrared sensors and found a reduction in the number of suicides. However, it had some methodological limitations, such as a lack of clarity on how outcomes were measured, a small amount of data, and a lack of statistical analysis. Moreover, it only provided information on prevented suicide attempts, not on suicide rates on the bridge. Another physical means restriction intervention with an additional element, reported by Sinyor et al. (2024), in which a 5-meter barrier illuminated by lights as a piece of art, appeared to reduce suicides drastically, but the lights were only installed years after the barrier itself, so it is not possible to make any assumptions about whether they had an additional effect.

Two studies examined a combination of interventions aimed at increasing the likelihood of third party intervention and help seeking (Kolves et al. 2023, Giraud 2021). Installing crisis line phones and CCTV cameras on an Australian bridge did not change the number of suicides in the three years after compared to the three years before. However, there was some statistical evidence that the downward trend in suicides started at the time these interventions were introduced (Kolves et al. 2023). It was not possible to examine their longer-term effects because barriers were installed on the bridge three years after the phones and CCTV. There was a small amount of data for analysis for each period, limiting the validity of the statistical analysis. The grey literature document that also described a combination of these two types of interventions only reported that reattendance was reduced to zero, and no other data was available (Giraud 2021).

A study that examined the installation of crisis phones on a bridge as a means of increasing help-seeking found that suicides increased in the 13 years after the introduction of the intervention, but this bridge had consistently received attention from media and from a website dedicated to reporting and discussing suicides on the bridge, potentially counteracting any positive effects that the intervention might have had (Stack 2015). Limitations of this study included issues with generalisability to other locations due to the specific features of the bridge, particularly in terms of the levels of attention from the media and online, and the lack of consistency in how outcomes were recorded because the information on them came from multiple sources, including media articles. Finally, another study that investigated the effect of placing of memorials, messages, or notes excluding official crisis line signage on motorway bridges found no difference in the number of suicides before and after (O’Neill et al. 2021).

### 2.3 Interventions at cliffs or other natural heights

#### 2.3.1 Overview

Interventions at cliffs or other natural heights were described in three studies which covered the same location and set of interventions (Lockley et al. 2014, Ross et al. 2020, Torok et al. 2023).

#### 2.3.2 Increasing opportunity for third-party intervention and help seeking, and physical means restriction with an additional or innovative element

The studies by Lockley et al. (2014), Ross et al. (2020), and Torok et al. (2023) described a set of interventions carried out at the same park with access to a cliff in Australia. The park is described as a 4.7-hectare big coastal escarpment area, and suicides have been recorded there since the 1800s (Lockley et al. 2014). The interventions included two crisis telephones, two signs with the crisis line number and a suicide prevention message, and CCTV cameras, all installed by February 2010, as well as landscaping work that included a new main entrance, improved seating, lighting, and tourist information displays, and a 1.3-meter-high fence along the clifftops with sensors that activated an alarm for the security monitoring service and alert police, completed in July 2011 (Lockley et al. 2014, Ross et al. 2020).

The earliest of the identified studies that examined the effectiveness of this set of interventions was by Lockley et al. (2014). There was a non-significant downward trend in jumping incidents between 2006 and 2012, with an estimated annual percentage change (EAPC) of −2.61% (95% CI −21.1–20.2; *p*=.760). The change in confirmed suicides between 2001 and 2011 was not significant either (EAPC=6.71%, 95% CI −2.5–16.8; *p*=.137). However, there was a significant increase in police call-outs related to individuals located at or approaching the park between 2006 and 2012 (EAPC=16.04%, 95% CI 7.1–25.7; *p*=.005). The change in police call-outs when an individual was located over the fence during the same time period was not significant (EAPC=-0.89%, 95% CI −22.1–26.0; *p*=.927). There was no numeric data on the use of the crisis line, but it was suggested that in a small number of cases the telephones had played an important role, either because the suicidal persons themselves used them or because they allowed bystanders to call for help.

Another study, which had both a quantitative and a qualitative element, published data for the period between 2000 and 2016 (Ross et al. 2020). This study found a non-statistically significant increase in suicides in the park over this period (APC=5.41%, 95% CI 0.38–11.53, *p*=.07). An examination of the breakdown of data by gender found that while the change in trend was not statistically significant for men (APC=6.23%, 95% CI 0.41–13.30, *p*=.06), it was significant for women, with suicides rising in the pre-intervention period of 2000–2010 (APC=16.64%, 95% CI 8.18–25.76, p<.001) and falling during and following intervention implementation in 2010–2016 (APC=-21.27%, 95% CI −33.14–-7.30, *p*=.01). During interviews, police officers trained in responding to suicidal individuals at the park highlighted the need to improve communication between emergency responders, hospital staff, and mental health teams. The authors of the study reported a consensus that while the fence was not a strong deterrent and was easy to climb, the CCTV and alarms were effective in preventing suicides through detection and location of individuals in crisis. However, it was pointed out that re-attempting individuals were aware that the CCTV and alarms would notify the police, which meant they might act faster to avoid being intercepted. In terms of the personal impact on police officers working in the park, some described the stress and a sense of responsibility to save people’s lives, as well as anxiety about the scrutiny in case they were not successful in doing so. Some interviewees described their own and their colleagues’ distress caused by witnessing suicides, and how seeking help could have a negative effect on them due to the stigma associated with mental health difficulties.

The most recent study covering this location, by Torok et al. (2023), focused on displacement from the immediate area of the park to other locations in the local and broader areas as well as method substitution. Data between 2006 and 2019, with 2006–2011 being pre-intervention and 2012–2019 post-intervention, was analysed. The authors did not detect any statistically significant changes in suicides during the study period in the park itself (0 join points; APC=-1.95%, 95% CI −6.9–3.3, *p*=.140), in the local area (0 join points; APC=6.81%, 95% CI −4.6–19.5, *p*=.226), or in the broader area (0 join points; APC=1.85%, 95% CI −7.4–12.1, *p*=.683). Regarding changes in suicides by different methods, the difference in the number of jumping deaths in the areas under examination was not significant (0 join points; APC=0.90%, 95% CI −3.9–5.9, *p*=.695), but there was a slight statistically significant increase in all suicide deaths in the city area (0 join points; APC=1.39%, 95% CI 0.1–2.7, *p*=.037).

#### 2.3.3 Bottom line results for interventions at cliffs or other natural heights

All three studies considered in this section related to the same location and set of interventions, which included crisis phones, signs with the crisis line number and a suicide prevention message, CCTV cameras, landscaping work, and a short fence with sensors. There was no statistically significant long-term reduction in suicides following the introduction of these interventions in the park, in the local area, or in the broader area (Torok et al. 2023). A shorter-term study found that suicides among women decreased after the start of the interventions, but there was no significant change in trend for men or overall (Ross et al. 2020). The studies used different cut-off dates for what they considered pre- and post-intervention periods because installation of the different elements of the set of interventions was carried out over a period of approximately 1.5 years. The main limitation of these studies is the volume of data available for analysis, which meant that the power to detect changes was low.

### 2.4 Interventions at tall buildings

#### 2.4.1 Overview

Tall buildings were considered in one study (Waalen et al. 2020).

#### 2.4.2 Increasing opportunity for help seeking

Waalen et al. (2020) described measures taken to prevent suicides on campus and at parking structures of a university in the USA. Several interventions were implemented: helpline signs at the roof perimeter in parking structures, banners and bungee cords in light wells, landscape improvements, planters and concrete bins, patio furniture and umbrellas. These interventions were implemented over a period between November 2013 and 2015. Around the same time, various physical means restriction interventions, such as fence barriers, wire mesh screens, and awnings, were also installed, so it is not impossible to attribute any changes to non-physical means restriction interventions exclusively. In an area on campus, six suicides occurred in 2002–2013, two in 2013, one in 2014, and none in 2015–2016. In the parking structures, there were eight suicides in 2002–2012, two in 2013, and none in 2014–2016.

#### 2.4.3 Bottom line results for interventions at tall buildings

Only one study was available for this type of location and it had some serious methodological issues. The interventions were not described clearly, the volume of data was not large enough to make conclusions, and no statistical analysis was performed. Besides, physical means restriction interventions were implemented in the same period, so no outcome change could be attributed to the other kinds of interventions.

### 2.5 Interventions at multiple types of locations

#### 2.5.1 Overview

Two studies covered interventions that happened at multiple types of locations. The study by Joyner et al. (2024a) included bridges (24.7%); railway or underground stations (17.3%); railway tracks (6.2%); roads (8.6%); tall buildings (21.0%); cliffs or other natural heights (4.9%); and parks, woodlands, or other green spaces (6.2%). The study by Owens et al. (2019) included bridges, railway or underground stations, tall buildings, and cliffs or other natural heights. In these studies, no breakdown of data by type of location was available.

#### 2.5.2 Increasing opportunity for third-party intervention

Joyner et al. (2024a) surveyed representatives of organisations such as local authorities (32%), emergency services (13%), health services (10%), rail industry (9%), and management of green spaces (4%) and properties (7%) from all regions of the UK about their perceptions of effectiveness of smart surveillance technologies (SSTs) to prevent suicides in public spaces. On a scale from 0–100, the median response regarding perceived effectiveness of SSTs was 50.00 (IQR=8.75–70.75). There was a lot of variation in perceived effectiveness of different types of SSTs. Drones were rated the highest (Med=73.00 [50.00–89.00]), followed by virtual fencing (Med=60.00, IQR=32.50–83.75), AI camera/video analytics systems (Med=60.00, [34–70]), Bluetooth Low Energy Beacons (Med=60 [15–75]), CCTV activated by movement/proximity (Med=55, IQR=20–72.5), and Automated Number Plate Recognition (Med=8.00 [0–70]). The respondents were also asked to rate other interventions, including radars, infrared sensors, online interceptive tools deployed over public Wi-Fi networks, and technology to digitally observe electronic devices and the combined median for those was 42 (IQR=14.50–84.25). Standalone interventions, such as those producing audible deterrents or visual alerts, were rated higher (Med=62.5, IQR=37.5–88.5) than those that initiated human response, such as alerting the control room or calling emergency services (Med=55.00, IQR=31.50–77.5). Moreover, interventions whose primary intended use was to prevent suicide or suicide attempts were rated more highly (Med=74.00, IQR=51.25–80.00) than those that had other primary uses, such as to prevent accidental injury or death, trespass, or crime or anti-social behaviour (Med=42.00, IQR=12.50–70). This difference was statistically significant (U=153.50, p=.009). This study only examined perceived effectiveness and did not provide information on changes in actual suicide numbers following the introduction of the interventions it discussed.

#### 2.5.3 Interventions initiated by bystanders

Owens et al. (2019) interviewed 21 people who had intervened in suicide attempts as bystanders, and 12 people who had been stopped from suicide. The interveners included 13 members of the public, six railway workers, including two off-duty at the time, and two highways officers. In total, 19 interventions were reported by the survivors and 31 by the interveners. Both interventions that included handover to services and those that did not were described as difficult. In the event of handover, the interveners sometimes felt excluded and at a loss, as well as afraid of what was going to happen to the person they helped, especially if they were arrested or sectioned under the Mental Health Act. If there was no handover, the interveners said it was difficult to judge when it was safe to leave the person. Some described being disturbed by the experience of intervention.

#### 2.5.4 Bottom line results for interventions at multiple types of locations

Limited data was available for interventions covering multiple types of locations. Participants in the study by Joyner et al. (2024a) rated the effectiveness of different SSTs and, unsurprisingly, interventions that were primarily aimed at preventing suicides rather than those that were initially designed for other purposes were rated as more effective. It was a cross-sectional study that was generally conducted well. The qualitative study by Owens et al. (2019) reported people’s experiences of intervening in suicide attempts, and of experiencing a bystander intervention. This study did not reflect on the potential influence of the researchers on the research, and it was unclear how the researchers were situated culturally or theoretically, but other than that, it was well-conducted.

### 2.6 Overall summary of the findings

#### 2.6.1 Changes in suicide numbers

In this rapid review, we focused on interventions other than physical means restriction, which has already received substantial attention in previous research and whose effectiveness is rarely disputed (Public Health Scotland 2022a, Public Health England 2015). We identified 24 studies reported in 29 documents, of which three studies reported on the same location and set of interventions.

The types of interventions described in these studies included increasing opportunity for third-party intervention; increasing opportunity for help seeking; creating a calming atmosphere; memorials or suicide prevention messages other than crisis line signage; interventions initiated by bystanders; and physical means restriction interventions with an additional or innovative element.

Within these broad types, a wide range of interventions has been identified, and in some cases an intervention included elements that fell within more than one category. Interventions aimed at increasing opportunity for third-party intervention included technological interventions such as CCTV, sensors, and suicidal behaviour recognition and alert systems as well as non-technological interventions such as training staff and encouraging bystanders to help people who appear to be in distress. Interventions aimed at increasing opportunity for help seeking included encouraging people to use suicide helplines by installing free phones and signs with the phone number, or just signs. In one study, creating a calming atmosphere through installing blue lights was tested. Additional or innovative elements to physical means restriction interventions included installing spinning rollers or sensors on fences and adding artistic lights.

Table 2 below summarises the evidence of effectiveness and important caveats of each of the included studies.

**Table 2.**
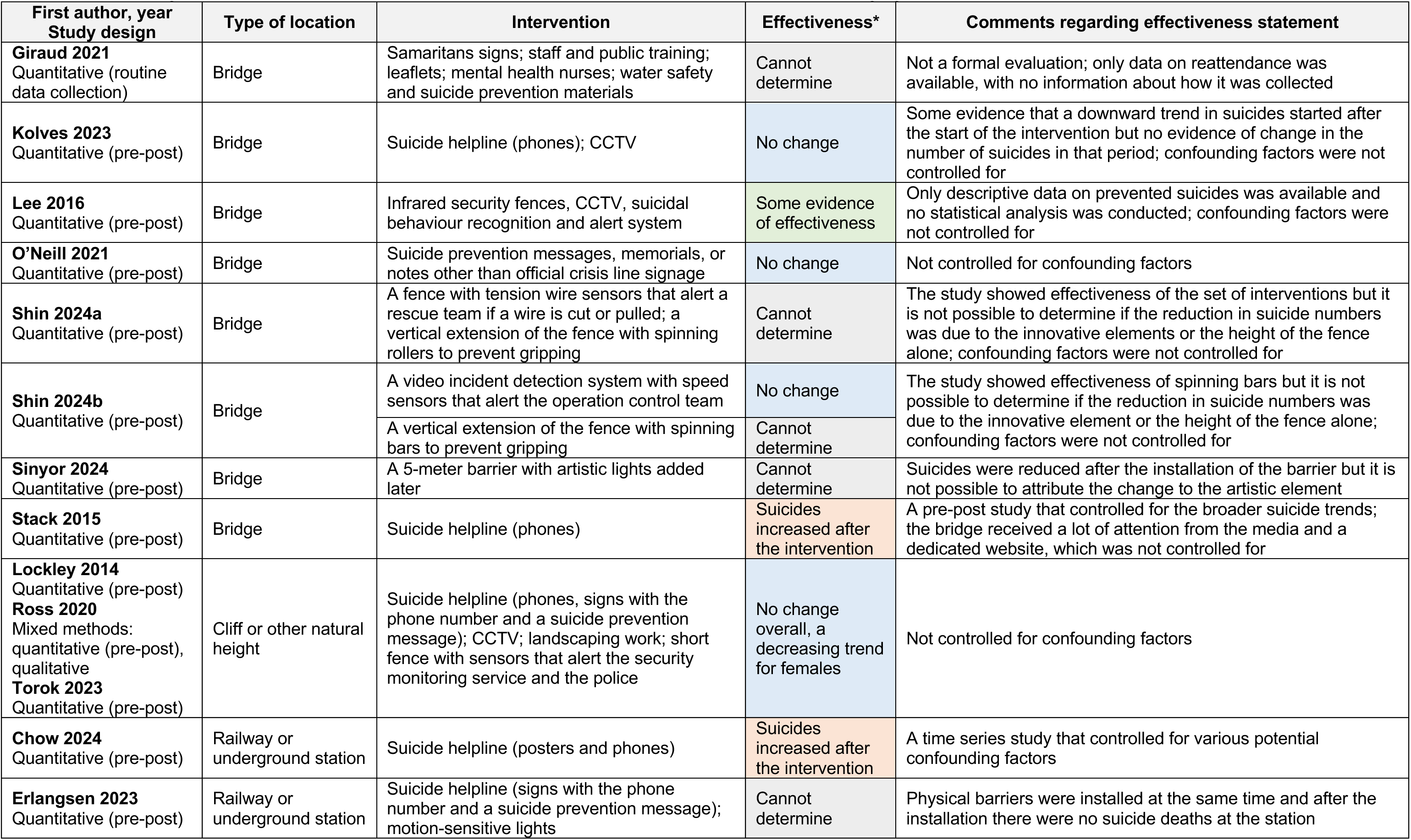
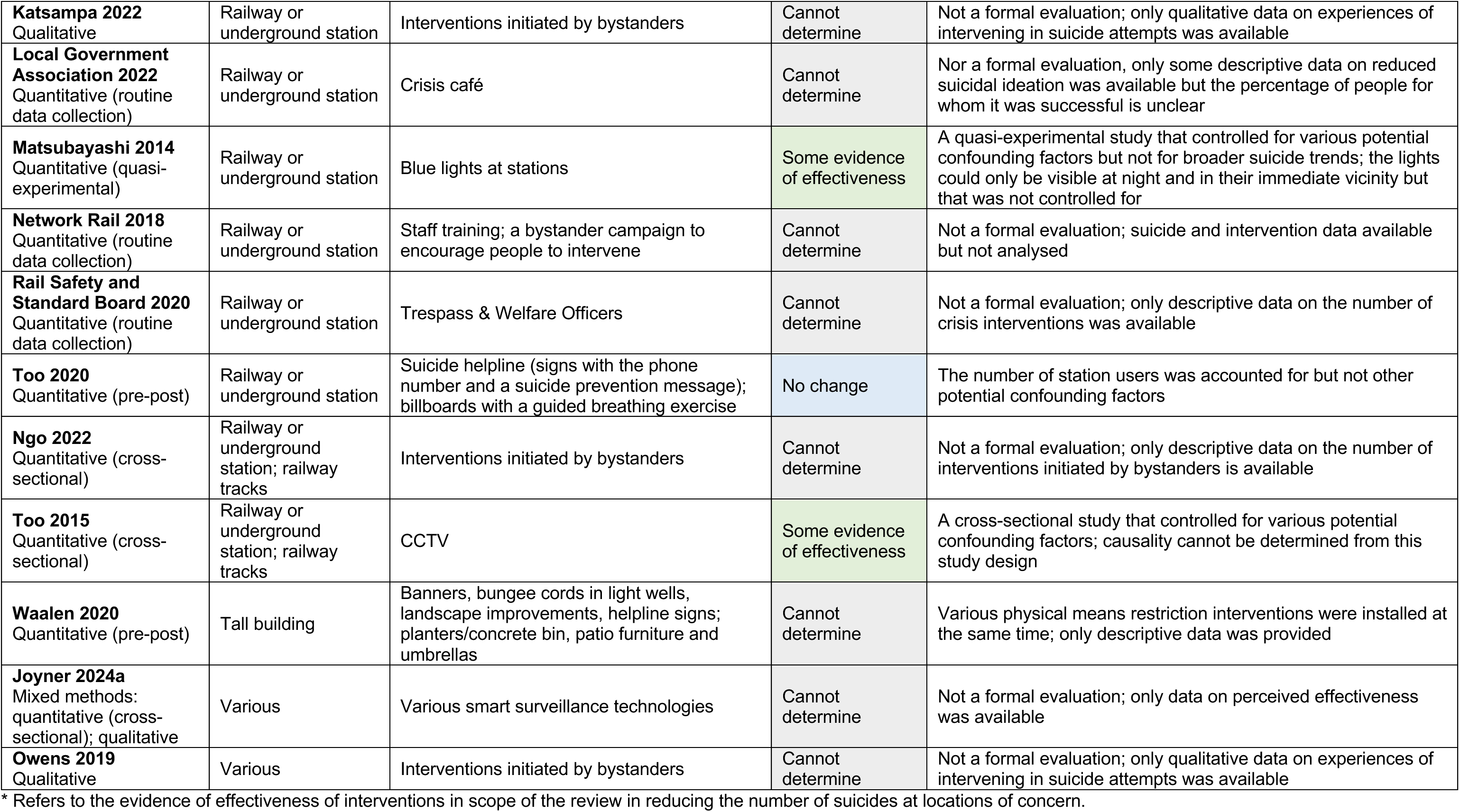
Summary of the effectiveness of intervention in the identified studies, sorted by type of location.

##### Potentially effective interventions

Technological interventions, i.e. surveillance technologies including CCTV, have shown the most promise, even though the evidence of their effectiveness was scarce and limited. In one study (Too et al. 2015), having more CCTV units was associated with fewer suicides at railway stations. It is not possible to establish whether the number of suicides was reduced because of the CCTV due to the cross-sectional design of this study. Another study (Lee et al. 2016) tested a set of interventions including CCTV, infrared security fences, and a suicidal behaviour recognition and alert system and provided some promising descriptive data that showed an increase in the number of prevented suicides, but a full evaluation that includes data on suicide rates, statistical analysis, and controls for confounding factors is needed to make any firmer conclusions. At the same time, there were studies that examined surveillance technology interventions that showed no change in outcomes after the start of the interventions. These will be discussed in the next sub-section.

The only other study that showed some evidence of effectiveness was one that examined the effects of installing blue lights at railway stations (Matsubayashi et al. 2014). It reported that suicides significantly decreased after the intervention but had a number of methodological limitations, such as not accounting for the fact that the lights were only visible at night and in their immediate vicinity. The authors also acknowledged that the underlying mechanism of action for this intervention was unclear. With than in mind, further research, including a replication study, is needed to make any firmer conclusion.

##### Interventions where no change was recorded

No change in suicides was observed as a result of five interventions or intervention sets. Promoting crisis helplines (Too et al. 2020), including in combination with CCTV (Kolves et al. 2023), and with CCTV and a whole range of other interventions including landscaping work and a short fence with sensors that alerted the security monitoring service and the police (Torok et al. 2023) showed no evidence of effectiveness. Another study (Shin et al. 2024b) showed no change after the introduction of a video detection system with speed sensors that alerted the operation control team if the speed of a car was too slow. The possible mechanism of action here would have been through identifying drivers that act unusually on a bridge and may be planning to stop with the intention to end their life, however, there are many other reasons why a car may be driving slowly. Finally, in another study (O’Neill et al. 2021), placing suicide prevention messages, memorials, or notes other than official crisis line signage on bridges had no effect on the number of suicides either.

There might have been important confounding factors that were not accounted for in these studies, so it is not possible to rule out that there were external factors that caused the number of suicides to increase and countered the effects of the interventions.

##### Interventions that may result in unintended harms

Two studies, both examining the effects of promoting crisis lines, reported that suicides increased after the start of the interventions. In the study by Chow et al. (2024), the number of suicides at an underground station increased after the installation of phones connected to a crisis line and posters. Although various potential confounding factors were controlled for in the study, there might have been others that were not accounted for that resulted in the increase in suicides over time. Another explanation is that installing the phones and posters contributed to the negative image of the location as a place where it is possible to die by suicide and attracted more people intending to end their life to it. The other study, by Stack (2015), examined the change in suicide numbers following the installation of crisis phones on a bridge and also reported that suicides increased after the intervention. The important confounding factor in this study that was not controlled for was the attention that suicides on the bridge received from the local press and a dedicated website, which might have contributed to maintaining the image of the bridge as a suicide location. Installing the phones could also have potentially exacerbated that.

##### Interventions whose effect could not be determined

Finally, there were studies in which it was not possible to determine the effectiveness of the interventions of interest to this review, due to their designs and/or quality, and in some cases because physical means restriction interventions were implemented at the same time. A few of them were not formal evaluations (Giraud 2021, Joyner et al. 2024a, Katsampa et al. 2022, Local Government Association 2022, Network Rail 2018, Ngo et al. 2022, Owens et al. 2019, Rail Safety and Standard Board 2020), so they did not provide data necessary to make a judgement regarding effectiveness. In five studies physical means restrictions were either installed at the same time as interventions in scope of this review (Erlangsen et al. 2023, Waalen et al. 2020) or were their intrinsic element (Shin et al. 2024a, Shin et al. 2024b, Sinyor et al. 2024), so no judgement regarding the effect of the non-physical means restriction intervention, or the innovative elements of the physical means restriction interventions alone, can be made based on the data provided in these studies even if they showed effectiveness of the overall set of interventions.

#### 2.6.2 Displacement to other locations

Another outcome of interest in this review is displacement to other locations. There are concerns that if suicides are prevented at one location, it may force individuals to choose a different location. Five of the included studies examined potential displacement (Kolves et al. 2023, Matsubayashi et al. 2014, Rail Safety and Standard Board 2020, Sinyor et al. 2024, Torok et al. 2023). The studies by Kolves et al. (2023) and Torok et al. (2023) reported no evidence of displacement, but they found no change in suicides at the locations of interventions either, so no conclusions can be made from this.

Sinyor et al. (2024), who found that the intervention was effective at reducing suicides at the location, examined changes in suicides in two other large metropolitan areas in the same province and found no change. However, people turning to a different location because their chosen one becomes inaccessible may not necessarily choose, or be able, to travel to a different city and no data on displacement to nearby locations was available.

Matsubayashi et al. (2014) found no systematic evidence of an increase in suicides at neighbouring railway stations after the introduction of the intervention, but they only had access to data from one railway company and no information about changes in suicides at stations in the same area managed by other companies was available.

The Rail Safety and Standard Board (2020) reported that they had observed displacement from station platforms to adjacent bridges, but no statistical analysis was conducted and no numeric data was provided, so it is unclear how that conclusion was made.

All things considered, there is not enough evidence in this review to make conclusions about the displacement effect.

#### 2.6.3 Method substitution

Only two studies examined whether interventions at locations of concern resulted in method substitution. One was by Torok et al. (2023) who reported an increase in the overall number of suicide deaths in the city but found no change in suicide numbers in the area of interest. The other study was by Sinyor et al. (2024), who found that there was no change in the number of suicides by other methods.

### 2.7 Assessment of the overall body of evidence

No formal assessment of the certainty in the overall body of evidence was performed as part of this rapid review, but here we summarise some of the main factors. Section 5.8 provides more information about how these were identified. Where possible, we assessed the confidence in the findings from the overall body of the quantitative evidence in terms of methodological quality, consistency across study findings, precision, directness of the evidence, and the possibility of publication bias. Due to the high heterogeneity in the interventions and locations, it was not possible to include all of the evidence in this assessment.

#### 2.7.1 The effect of interventions aimed at increasing opportunity for help seeking on the number of suicides

Firstly, we focus on promoting suicide helplines as an intervention aimed at increasing opportunity for help seeking. Three studies of interventions at railway or underground stations (Chow et al. 2024, Erlangsen et al. 2023, Too et al. 2020), two at bridges (Kolves et al. 2023, Stack 2015), one at tall buildings (Waalen et al. 2020), and three reporting on the same location and intervention at a cliff (Lockley et al. 2014, Ross et al. 2020, Torok et al. 2023) reported on this kind of interventions. Only in two of them (Chow et al. 2024, Stack 2015) suicide helplines were not combined with another kind of intervention. In the other studies, they were combined with motion lights and physical barriers (Erlangsen et al. 2023), CCTV (Kolves et al. 2023), CCTV, landscaping work, and a short fence with sensors that alert the security monitoring service and the police (Lockley et al. 2014, Ross et al. 2020, Torok et al. 2023), billboards with a guided breathing exercise (Too et al. 2020), or a range of interventions including banners, bungee cords in light wells, landscape and interior design improvements, and physical barriers (Waalen et al. 2020). The presence of the other interventions is a limitation of this assessment since it is impossible to untangle the effect of the crisis phone lines from that of the rest of the interventions.

The first dimension we focus on is the methodological quality of these studies, and the risk of bias resulting from it. All of these studies used a pre-post design. They varied considerably in quality, from not having serious methodological concerns (Chow et al. 2024), to having some issues, particularly related to confidence in the findings being reduced due to the small amount of data available for analysis (Kolves et al. 2023, and the three studies reporting on the same location and set of interventions: Lockley et al. 2014, Ross et al. 2020, Torok et al. 2023), to having a number of number of methodological issues limiting the validity of the findings (Erlangsen et al. 2023, Stack 2015, Too et al. 2020, Waalen et al. 2020). In addition, only three of these studies (Chow et al. 2024, Stack 2015, Too et al. 2020) accounted for at least some confounding factors. Therefore, there is a high risk of bias related to the outcomes of this kind of intervention. Section 6.3 provides more detailed information on the quality assessment of each study and Section 5.6 explains how it was performed.

The next area is consistency across study findings. While Chow et al. (2024) and Stack (2015) reported that suicides increased after the introduction of the intervention, Kolves et al. (2023), Too et al. (2020) and, reporting on the same location and set of interventions, Lockley et al. (2014), Ross et al. (2020), and Torok et al. (2023) observed no change. In the case of the studies by Erlangsen et al. (2023) and Waalen et al. (2020), the effect could not be determined. No studies reporting on promoting suicide helplines found that they resulted in a reduction in the number of suicides. Overall, there was little consistency in the findings, with some reporting an increase in suicides and others reporting no change.

Regarding precision, all of the studies had a small amount of data available for the analysis apart from Chow et al. (2024), in which whether it was large enough to provide confidence in the findings or not could not be determined, and Stack (2015), which was judged to have sufficient data. The confidence intervals were narrow in the Chow et al. (2024) study, but there was less precision in the Kolves et al. (2023), Too et al. (2020), Lockley et al. (2014), Ross et al. (2020), and Torok et al. (2023) studies. Confidence intervals were not reported in the remaining three studies (Erlangsen et al. 2023, Stack 2015, Waalen et al. 2020), although Stack (2015) provided standard errors. Overall, there was low precision in these findings.

The evidence was direct in all of the studies since they all examined changes in suicides at locations of concern, so no proxy outcomes were used. The final dimension is publication bias. In this type of review, it is not possible to formally assess publication bias, however, we took steps to counter it by extensively searching grey literature.

All in all, the assessment of the overall body of evidence indicates that there is a low level of confidence in the evidence for this outcome.

#### 2.7.2 The effect of surveillance technologies on the number of suicides

In this section, we assess the overall body of evidence related to surveillance technologies, as a sub-category of interventions aimed at increasing opportunity for third-party intervention. Most of these were CCTV, but some studies also included technologies such as sensors, video detection and alert systems, and others. These kinds of interventions were examined in nine studies (Joyner et al. 2024a, Kolves et al. 2023, Lee et al. 2016, Lockley et al. 2014, Ross et al. 2020, Torok et al. 2023, Shin et al. 2024a, Shin et al. 2024b, Too et al. 2015), which included three studies of the same location and set of interventions (Lockley et al. 2014, Ross et al. 2020, Torok et al. 2023). Of the nine studies, four were at bridges (Kolves et al. 2023, Lee et al. 2016, Shin et al. 2024a, Shin et al. 2024b), three reporting on the same location and interventions at a cliff (Lockley et al. 2014, Ross et al. 2020, Torok et al. 2023), one at railway stations and railway tracks (Too et al. 2015), and one included different kinds of locations (Joyner et al. 2024a). Some of these studies included some other kinds of interventions as well, including a suicide helpline (Kolves et al. 2023), a suicide helpline and landscaping work (Lockley et al. 2014, Ross et al. 2020, Torok et al. 2023, reporting on the same location and set of interventions), and vertical extension of the fence with spinning rollers to prevent gripping (Shin et al. 2024a). As in the previous section, this is a limitation in the assessment of the overall body of evidence related to surveillance technologies as an intervention aimed at increasing opportunity for third-party intervention. The study by Kolves et al. (2023) also evaluated a suicide helpline and was included in the previous section of this assessment as well.

Regarding methodological limitations, and the risk of bias resulting from them, again there was a lot of variability between the studies. The cross-sectional study by Too et al. (2015) was considered to be well-conducted but its design precludes us from making assumptions about the causal relationship between the intervention and the number of suicides. Most of the other studies were fairly well-conducted, but included some methodological limitations, such as having too little data available for analysis to give confidence in the findings (Kolves et al. 2023, Lee et al. 2016, Lockley et al. 2014, Ross et al. 2020, Torok et al. 2023, Shin et al. 2024b). One of the studies included a larger number of methodological limitations, including a lack of clarity regarding how the outcomes were determined and a lack of statistical analysis (Lee et al. 2016). Of the studies that examined these types of interventions, only one (Too et al. 2015) controlled for confounding factors. Overall, there is a high risk of bias related to the outcomes of surveillance technology interventions.

In terms of consistency in the findings, only two studies found some evidence of effectiveness of these interventions (Lee et al. 2016, Too et al. 2015), but it was limited either by methodological issues (Lee et al. 2016) or the study design (Too et al. 2015). The rest of the studies either reported no change (Kolves et al. 2023, Shin et al. 2024b, as well as the three studies on the same location and interventions: Lockley et al. 2014, Ross et al. 2020, Torok et al. 2023), or the effect could not be determined because no formal evaluation was conducted (Joyner et al. 2024a) or it could not be untangled from physical means restriction interventions within the study (Shin et al. 2024a). With some studies indicating some evidence of effectiveness and others showing no effect, there was little consistency regarding this kind of interventions.

In terms of precision, most of these studies (Kolves et al. 2023, Lee et al. 2016, Shin et al. 2024b, as well as Lockley et al. 2014, Ross et al. 2020, Torok et al. 2023) had little data available for analysis to give confidence in the findings. Regarding confidence intervals, they were quite narrow in the study by Too et al. (2015), but less precise in the rest of the studies where they were available (Kolves et al. 2023, Shin et al. 2024a, Shin et al. 2024b, Lockley et al. 2014, Ross et al. 2020, Torok et al. 2023). In the study by Joyner et al. (2024a) only response ranges were provided and they tended to be wide. No indication of precision was available in the remaining study (Lee et al. 2016).

All of these studies examined the effect of surveillance technologies on suicide numbers directly, apart from the one by Lee et al. (2016), who reported prevented suicide attempts rather than the number of suicides, and Joyner et al. (2024a), who used perceptions of effectiveness as the outcome. As previously stated, although in this type of review it was not possible to formally assess publication bias, we took steps to reduce it by performing grey literature searches.

Based on the assessment of the overall body of the evidence, there is a low level of confidence in the findings related to surveillance technologies.

## 3. DISCUSSION

The Scottish national guidance on action to address suicides at locations of concern (Public Health Scotland 2022a) uses the integrated motivational volitional (IMV) model of suicidal behaviour as a theoretical framework for understanding the transition from suicidal ideation to action. This model includes three stages: 1) pre-motivational, that includes background factors and triggering events; 2) motivational, during which intentions are formed; 3) and volitional, also referred to as behavioural enaction (O’Connor & Kirtley 2018). During the last stage, access to means of suicide is considered an important factor in determining whether suicidal ideation/intent will transition to suicidal behaviour.

Therefore, as previous research has shown (Okolie et al. 2020b, Pirkis et al. 2015), preventing physical access is important, but removing means of suicide does not only entail installing fences and nets. As the Public Health Scotland (2022a) national guidance explains, interrupting the suicidal process is the aim of actions to reduce suicides at locations of concern. This may include interventions by other people, such as emergency services, railway staff, or bystanders, or any other sort of intervention that delays or disrupts suicidal behaviour long enough to allow the individual in distress to receive help (Public Health England 2015). Heightened risk of suicide is often short-term, so interrupting a suicidal act and providing timely support can be life-saving (World Health Organization 2014).

To provide policymakers and organisations and individuals managing locations of concern with an evidence base to allow them to choose from a range of interventions at specific locations, the present review focused on any interventions other than physical means restriction aimed at reducing suicides at public locations. However, we found that research aimed at evaluating such intervention was limited and had many methodological limitations. As discussed previously in this report in detail, some technology-based interventions appeared to show promise, but the effectiveness of the other types of identified interventions could not be determined based on the available research. The main takeaway of this review is that more robust research is needed to create a high-quality evidence base.

### 3.1 Strengths and limitations of the available evidence

Twenty-four studies were included in this review, covering a wide range of interventions and types of public locations of concern. Most of these were uncontrolled pre-post studies, with three cross-sectional studies that do not evidence a causal relationship between the interventions and outcomes, and only one controlled quasi-experimental study which, despite its design, also had methodological limitations. Some qualitative evidence regarding the experience of intervening to prevent a suicide attempt was also available.

The overall quality of the identified evidence is quite low and precludes us from making firm conclusions about the effectiveness of any of the included interventions. In addition to methodological issues present in almost all of the included studies, the quality of reporting was low, with the interventions often poorly described. Most of the included studies were based on little data and did not account for confounding factors, which limits the validity of their findings. Some of the included literature provided some descriptive data but did not include a formal evaluation of the interventions. This was especially true of the identified grey literature. A few of the studies included physical means restrictions introduced at the same time as the interventions of interest in this review, so it was not possible to determine if any of the observed effect could be attributed to the interventions in scope of the review.

Despite these limitations, some of the included studies provided some initial evidence regarding potentially effective interventions that future research may examine in more detail.

### 3.2 Strengths and limitations of this rapid review

The question and the eligibility criteria for this review were developed in consultation with stakeholders from the NHS Wales Executive, a Health and Care Research Wales Evidence Centre Public Partnership Group member, and a representative from Public Health Scotland. We conducted extensive literature searches including systematic searches of nine bibliographic databases of academic literature and supplementary searches of almost fifty websites of relevant government, third sector, and research organisations and a grey literature database. We also reviewed literature citing, and cited by, the already identified studies as well as that included in existing relevant systematic reviews. The literature was independently screened for inclusion by two reviewers. This allowed us to maximise the amount of identified relevant evidence. All of the academic literature was critically appraised and its methodological limitations considered in the interpretation of the findings.

The limitations of this review are as follows. First, due to it being a rapid review conducted within a limited timeframe, only grey literature from the UK was included, although academic literature from any country was accepted. Even though this review is primarily targeted at UK policymakers and managers of locations of concern, there may be international grey literature relevant to decision making. The grey literature was not formally critically appraised, but since it did not include formal evaluations, it did not have a substantial influence on our interpretation of the findings.

Regarding literature searches, we suspect that there may be more studies in scope of this review that evaluated physical means restriction interventions with an additional or innovative element that we were unable to identify because the interventions were not described in sufficient detail in the publications. For example, we were only able to identify the study by Sinyor et al. (2024) as being in scope because the name of the barrier described in it suggested that the intervention included an artistic element, which we verified through a report in a newspaper local to the site.

Another limitation is in the critical appraisal of the included studies. The critical appraisal checklists were designed for studies of human populations, not locations, so some of the questions, particularly in the checklist for pre-post studies, were not applicable. We countered this limitation by using the checklists only as a guide for the appraisal and using our own judgement, in discussion within the review team, when determining the quality of the studies and considering it in reporting their results.

### 3.3 Implications for policy and practice

The main finding of this review is that more robust evaluations are needed before any of the reviewed interventions can be recommended for implementation. There is some initial evidence that surveillance-based interventions aimed at increasing opportunity for third-party intervention may be effective, however, more research is needed. We also identified through grey literature that there are many initiatives happening across the UK, however, they are either not being evaluated, or the evaluations are not being made public. To create a better evidence base, robust evaluations should be supported and encouraged, and their findings shared so that places can learn from each other’s experience.

### 3.4 Implications for future research

There is an urgent need for more high-quality research evaluating interventions aimed at reducing suicides at locations of concern other than physical means restriction. From our searches of grey literature, we know that such interventions are being implemented in various places across the UK (e.g., the “Small Talk Saves Lives” or “Make a Connection” campaigns across the rail networks, or a set of different interventions in the City of London), however, the evidence of their effectiveness is lacking.

Concerningly, we identified two studies that showed that suicides increased at the locations after the introduction of interventions promoting the use of suicide helplines, which may potentially be explained by the visibility of the phones and signs contributing to the negative image of the locations as places where many suicides happen, although there may be other factors obscuring the relationship between the interventions and the number of suicides.

This shows the need for robust evaluations to create a strong evidence base that allows policymakers and managers of locations of concern to ensure that evidence-based interventions are used to prevent unintended harm and save resources.

Another important avenue for future research is to understand which interventions work for who and in what circumstances. For example, future studies may examine whether different demographic groups experience interventions differently and have different outcomes.

Our literature searches identified a number of studies reporting the development of artificial intelligence models that aim to detect suicidal behaviour (Li et al. 2022, Onie et al. 2023, Yogesan et al. 2023). We anticipate that once such models are finalised and implemented, there may be future evaluation studies assessing their effectiveness in preventing suicides at public locations.

##### 3.5 Economic considerations^*^

The loss of life due to suicide in Wales could cost the Welsh economy at least the Welsh £537 million1 each year. As well as being a tragic event for families and communities, suicides can cost the economy at least the Welsh £1.6 million^1^ per every life lost (Samaritans 2024a).

The largest contributions to this economic loss are intangible costs (costed using HM Treasury Green book guidance of the Welsh £60,000 per statistical year of life). Other considerable impacts include future potential employment and productivity, emergency service callout, healthcare, and potential productivity losses.

Suicides in public spaces pose unique economic impacts. The Office of Rail and Road calculated the rail and road industry faces a cost of the Welsh £306,0001 per suicide event (Prosser 2022).

Future research evaluating interventions aimed at reducing suicides at public locations should consider the economic impacts of suicides in such locations from a wider societal perspective. Future studies may wish to utilise a cost-of-illness methodology, such as those identified in the review by Jain et al. (2024).

## 5. RAPID REVIEW METHODS

The protocol for this rapid review was published on the OSF website before the review process commenced (Kiseleva et al. 2024). It is available through the following URL: https://doi.org/10.17605/OSF.IO/3XGRA.

### 5.1 Eligibility criteria

The inclusion and exclusion criteria for this review, developed in consultation with stakeholders from the Welsh Government, a Health and Care Research Wales Evidence Centre Public Partnership Group member, and a representative from Public Health Scotland, are provided in Table 3. These criteria guided the literature search and selection process.

**Table 3.** Eligibility criteria.

|  | **Inclusion criteria** | **Exclusion criteria** |
| --- | --- | --- |
| **Problem / place** | Public locations where suicide attempts and deaths happen, including but not limited to:<br>• Tall buildings, bridges, cliffs, and other manufactured or natural structures that allow access to a height<br>• Rural/secluded locations (woodlands, parks etc)<br>• Railway lines and stations<br>• Roads<br>• Locations with access to water<br>• Heritage or historical sites | Non-public locations, such as homes or other residential settings |
| **Intervention** | • Interventions aimed at increasing the likelihood of third-party intervention<br>• Interventions aimed at increasing help seeking/help giving<br>• Non-barrier design interventions aimed at creating a calming atmosphere<br>• Non-barrier design interventions aimed at reducing fatality<br>• Means restriction interventions with an innovative or artistic element, such as planting bushes<br>• Suicide prevention messages, memorials, artworks<br>• Community engagement, including but not limited to designing and delivering interventions<br>• Media campaigns<br>• Placement of animals around the location<br>• Any other intervention other than physical means restriction | Means restriction through creating physical barriers such as fencing or netting or otherwise preventing physical access to a location |
| **Comparison** | • Any other intervention<br>• No intervention<br>• No comparison |  |
| **Outcomes** | Primary outcomes:<br>• Suicide rates at locations of concern<br>• Suicide ideation at locations of concern<br>• Suicide attempts at locations of concern<br>Secondary outcomes:<br>• Displacement to other similar nearby locations<br>• Suicide method substitution<br>• Impact on communities (individuals, family, witnesses, first responders, service delivery, infrastructure, financial) |  |
| **Study design** | Any quantitative, qualitative, or mixed methods study design |  |
| **Countries** | Academic literature: No limits<br>Grey literature: UK |  |
| **Language** | English |  |
| **Publication date** | From 2014 |  |
| **Publication type** | • Published academic studies<br>• Grey literature reports | • Conference abstracts<br>• Editorials<br>• Comments<br>• Book chapters<br>• Theses and dissertations |

### 5.2 Literature search

#### 5.2.1 Evidence sources

The following bibliographic databases were systematically searched: MEDLINE via Ovid, PsycInfo via Ovid, Cochrane Library (CDSR and CENTRAL), Scopus, Sociology Collection (Applied Social Sciences Index & Abstracts, Sociological Abstracts, Sociology Database) via ProQuest, Social Science Database via ProQuest.

The policy and grey literature database Overton and websites of relevant government, third sector, and research organisations were searched for grey literature. Forward and backward citation searching was performed on the relevant primary studies identified through database searches.

During preliminary work, we identified or were supplied with a number of existing rapid (Public Health Scotland 2022b) and systematic (Barker et al. 2017, Chamberlain & Woodnutt 2024, Havârneanu et al. 2015, Okolie et al. 2020a, Okolie et al. 2020b, Pirkis et al. 2015) reviews of interventions targeted at reducing suicides at locations of concern. These reviews, and any other reviews that we identified during literature searches, were unpicked and relevant primary studies included in this review.

#### 5.2.2 Search strategy

A search strategy was developed in MEDLINE via Ovid by one review member and checked by two other members. The stakeholders provided input into the search terms. The search strategy was then translated for the other bibliographic databases before searches were performed in them.

Details of the database searches are presented in Appendix 1. The list of searched websites is provided in Appendix 2.

Literature published since 2014 was searched for. The bibliographic database searches were conducted at the end of October 2024, the Overton database was searched in November 2024, citations were searched in December 2024, reviews were unpicked in January 2025, and websites were searches between November 2024 and January 2025.

#### 5.2.3 Reference management

Identified references were exported from the bibliographic databases and deduplicated using the reference management software EndNote, after which they were uploaded into the online screening tool Rayyan, where deduplication was completed.

### 5.3 Study selection process

#### 5.3.1 Literature identified through bibliographic databases, review unpicking, and citation searching

All records identified through database searches, review unpicking, and citation searching were screened independently by two reviewers in Rayyan based on the information provided in the titles and abstracts. Where a record appeared to meet the eligibility criteria, or if a decision could not be made based on the information in the titles and abstracts alone, it was retained and proceeded to the full-text screening stage. Full-text screening was performed by two reviewers independently. Conflicts were resolved through discussion, involving a third reviewer where necessary.

#### 5.3.2 Literature identified through grey literature database and website searches

The Overton database and website searches were performed by a single reviewer. On Overton, the first ten pages (500 results) were reviewed and potentially relevant documents retrieved. For searching most websites, both the integrated search function where available and Google Advanced Search were used. If >100 results were returned, a decision was made whether to refine the search to reduce the number of results or only screen the first few pages. Retrieval of relevant literature was maximised by using two search methods on each website where possible. The identified documents were briefly scanned at full text by a single reviewer and those that appeared to meet the eligibility criteria, or where a decision could not be made without a more careful examination, were proceeded to be screened independently by two reviewers, with any conflicts resolved through discussion and involving a third reviewer where necessary.

### 5.4 Data extraction

Data extraction was performed by one reviewer and checked by another. The following data were extracted where available: country, study aim, study design, dates of data collection, data collection methods, details of the location(s) of concern, details of the intervention and, where applicable, comparison, and relevant findings. The data extraction table was first piloted on a selection of studies of different designs.

### 5.5 Study design classification

Study design classification was performed for the purpose of selecting the most appropriate quality appraisal checklist by one reviewer and checked by another, resolving any disagreements through discussion or by involving a third reviewer where necessary. No formal identification algorithm was used.

### 5.6 Quality appraisal

The following critical appraisal checklists were used to assess the methodological quality of the studied, depending on the study design: the Quality Assessment Tool for Before-After (Pre-Post) studies with no control group (National Heart Lung and Blood Institute 2013), the JBI Checklist For Analytical Cross Sectional Studies (Moola et al. 2020), the JBI Checklist For Quasi-Experimental Studies (Barker et al. 2023), or the JBI Critical Appraisal Checklist For Qualitative Research (Lockwood et al. 2015). The quality appraisal was conducted by one reviewer and checked by another, with any disagreements resolved through discussion or by involving a third reviewer where necessary. Because these critical appraisal checklists were designed for studies of human populations and not locations, some of the questions were not applicable. We interpreted the population questions of the checklists as referring to locations (e.g., whether the location was representative of other locations where the same intervention could be used).

### 5.7 Synthesis

Data was synthesised narratively according to types of locations and interventions. Consideration was given to the methodological limitations of the included studies when reporting the results.

### 5.8 Assessment of body of evidence

No formal assessment of the overall body of evidence was performed, however, the dimensions included in the Grading of Recommendations, Assessment, Development and Evaluation (GRADE) approach (Schünemann et al. 2023) were considered. Therefore, when narratively describing the overall body of the quantitative evidence, where possible, we reflected on the risk of bias, imprecision, inconsistency, and indirectness of the evidence as well as possible publication bias.

## 6. EVIDENCE

### 6.1 Search results and study selection

**Figure 1.**
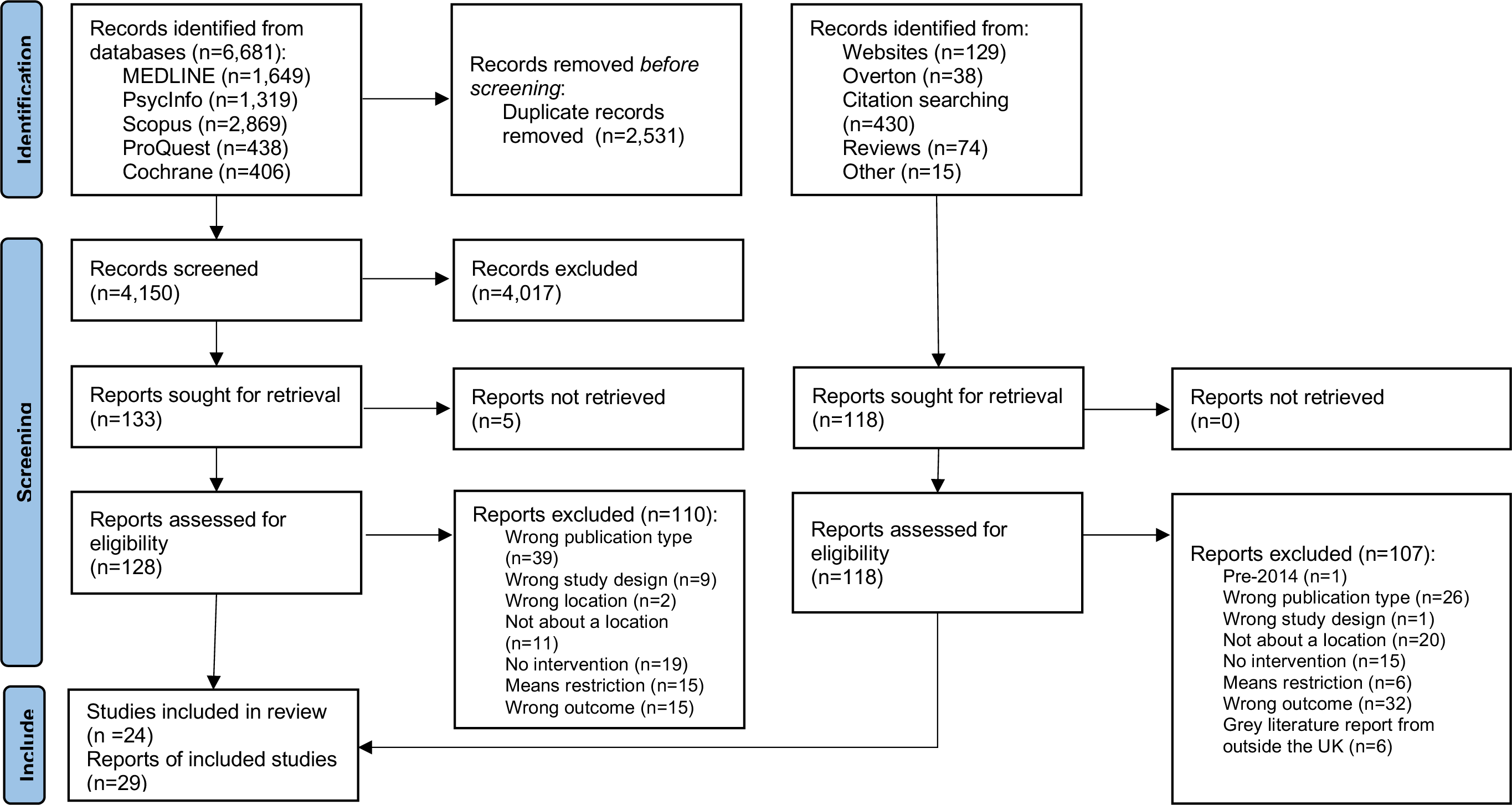
PRISMA flow diagram.

### 6.2 Data extraction

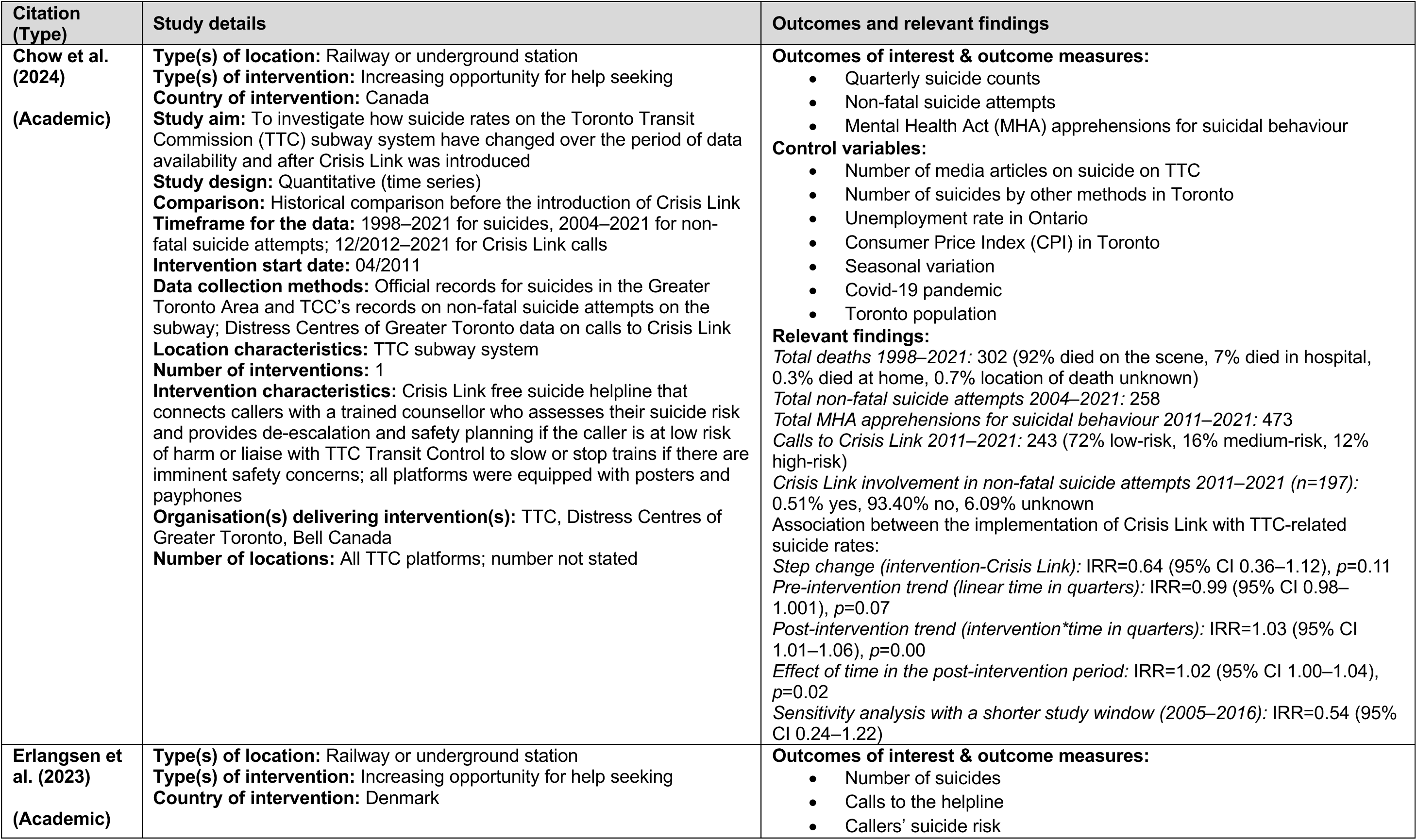
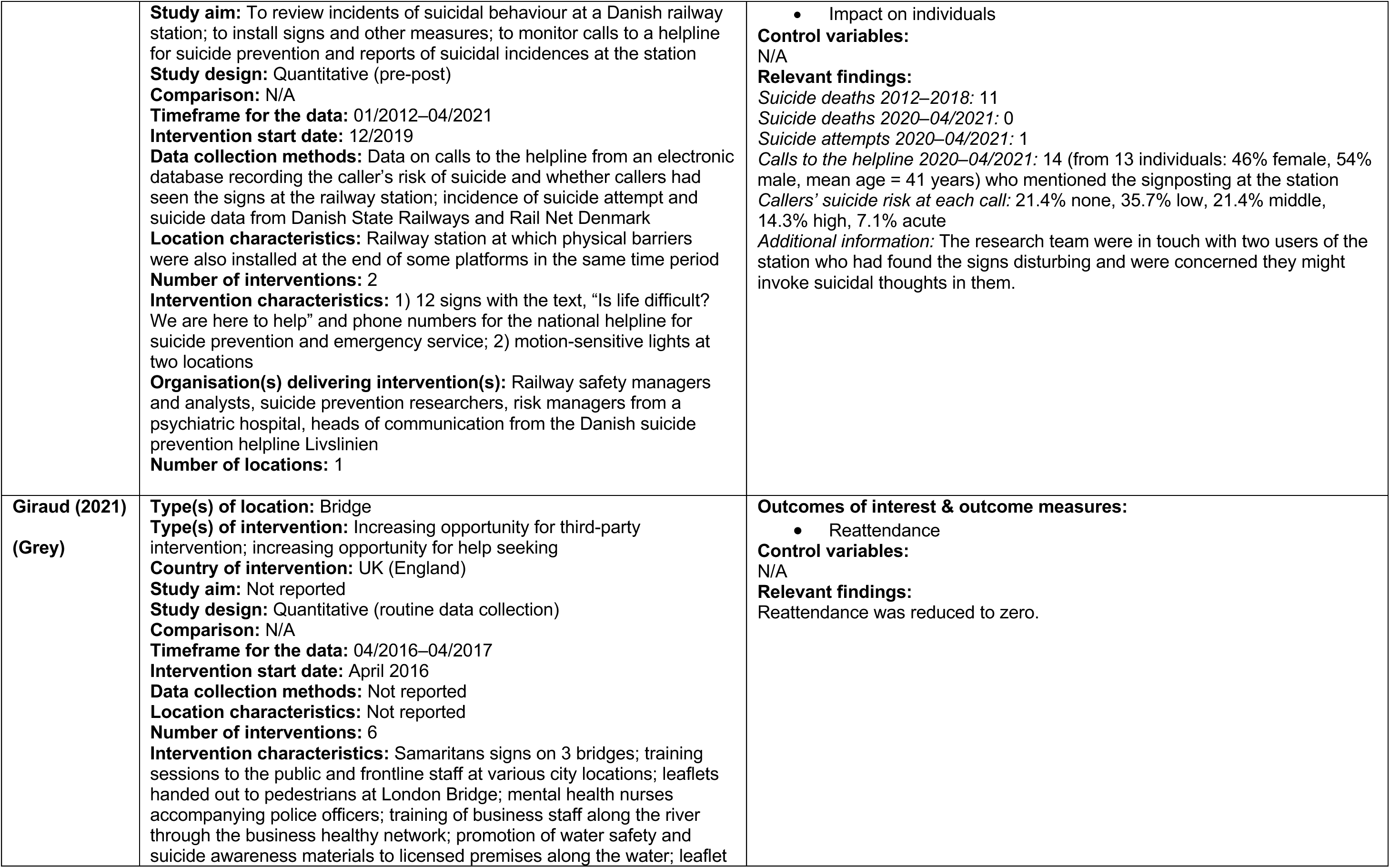
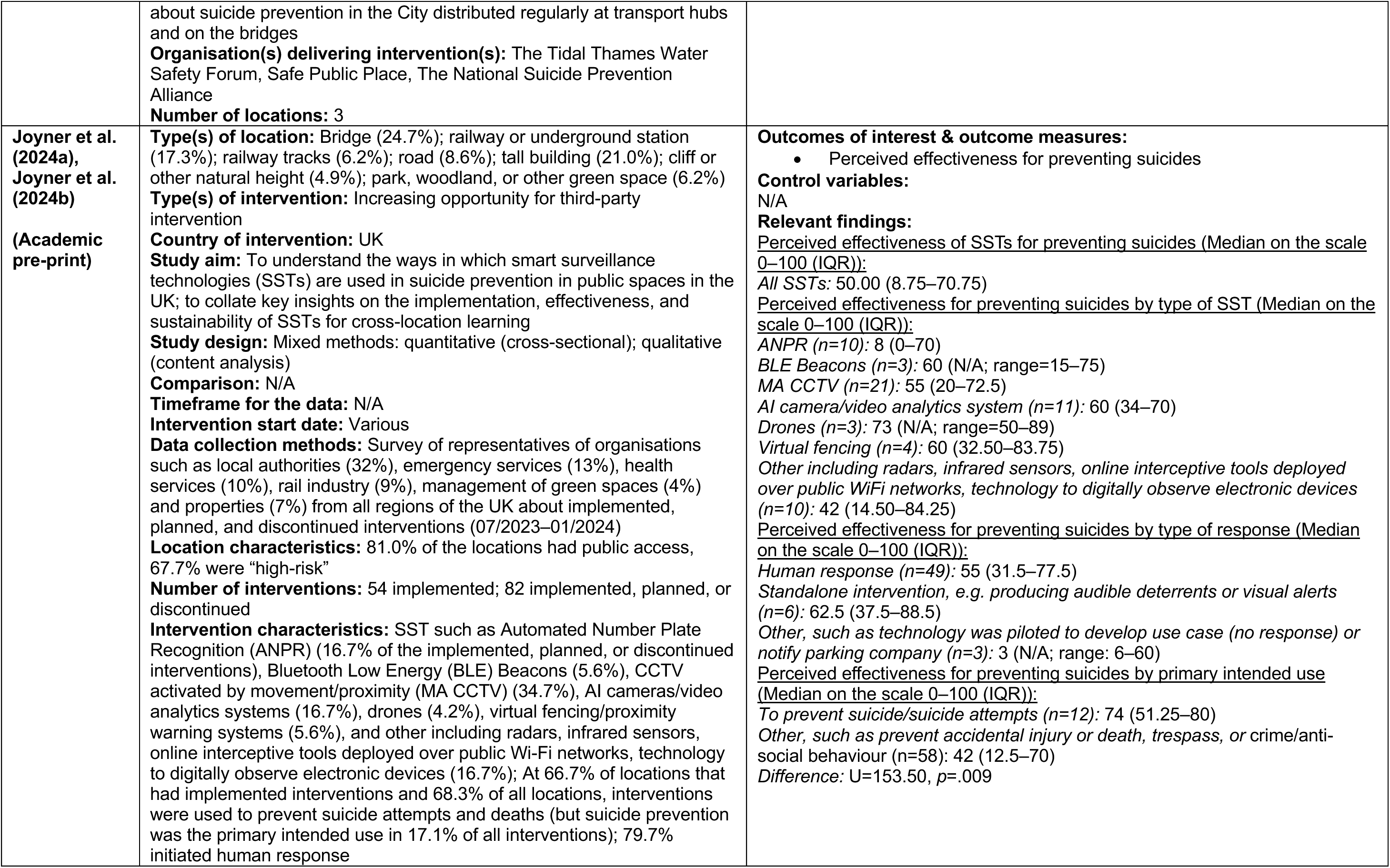
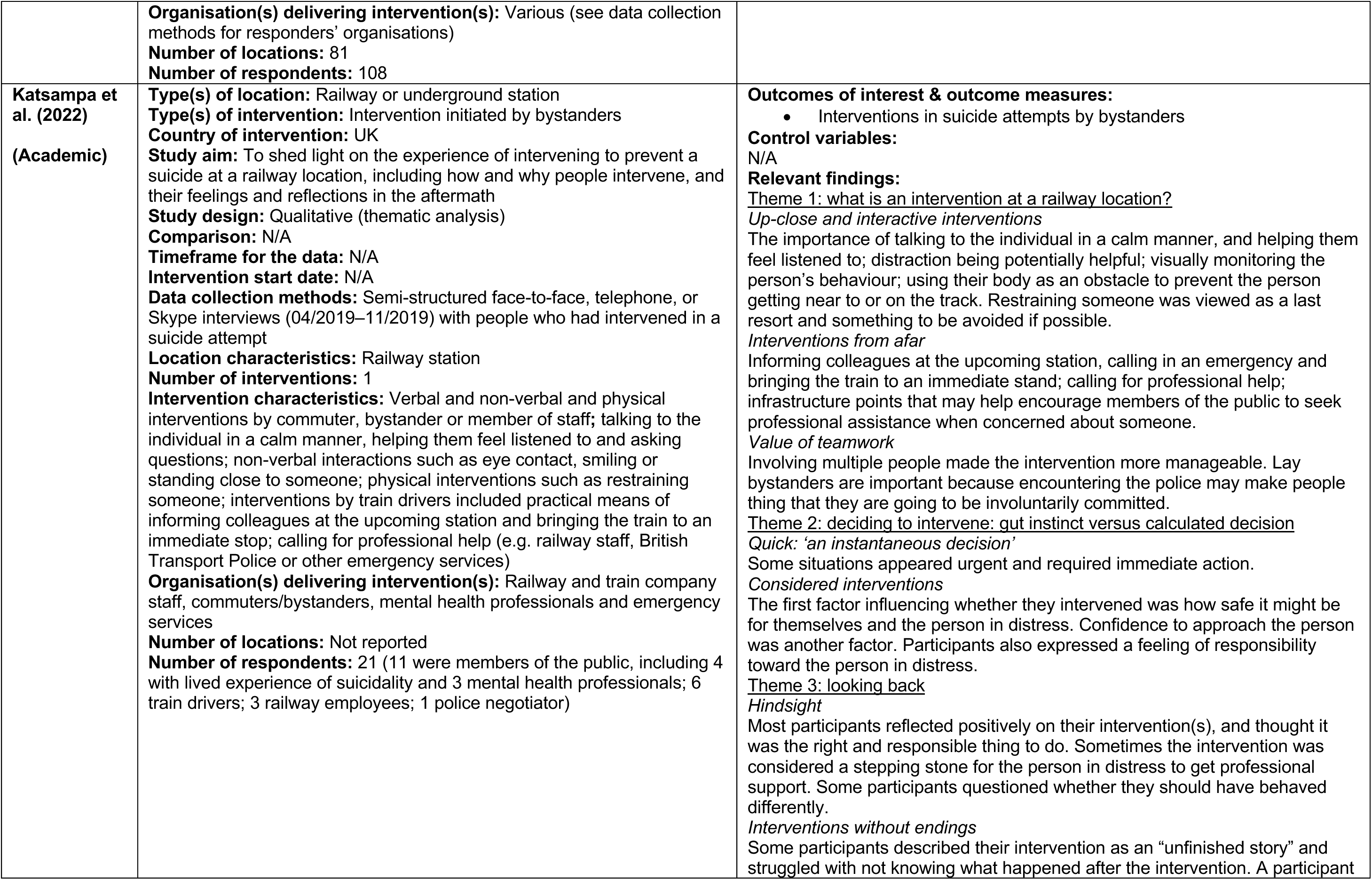
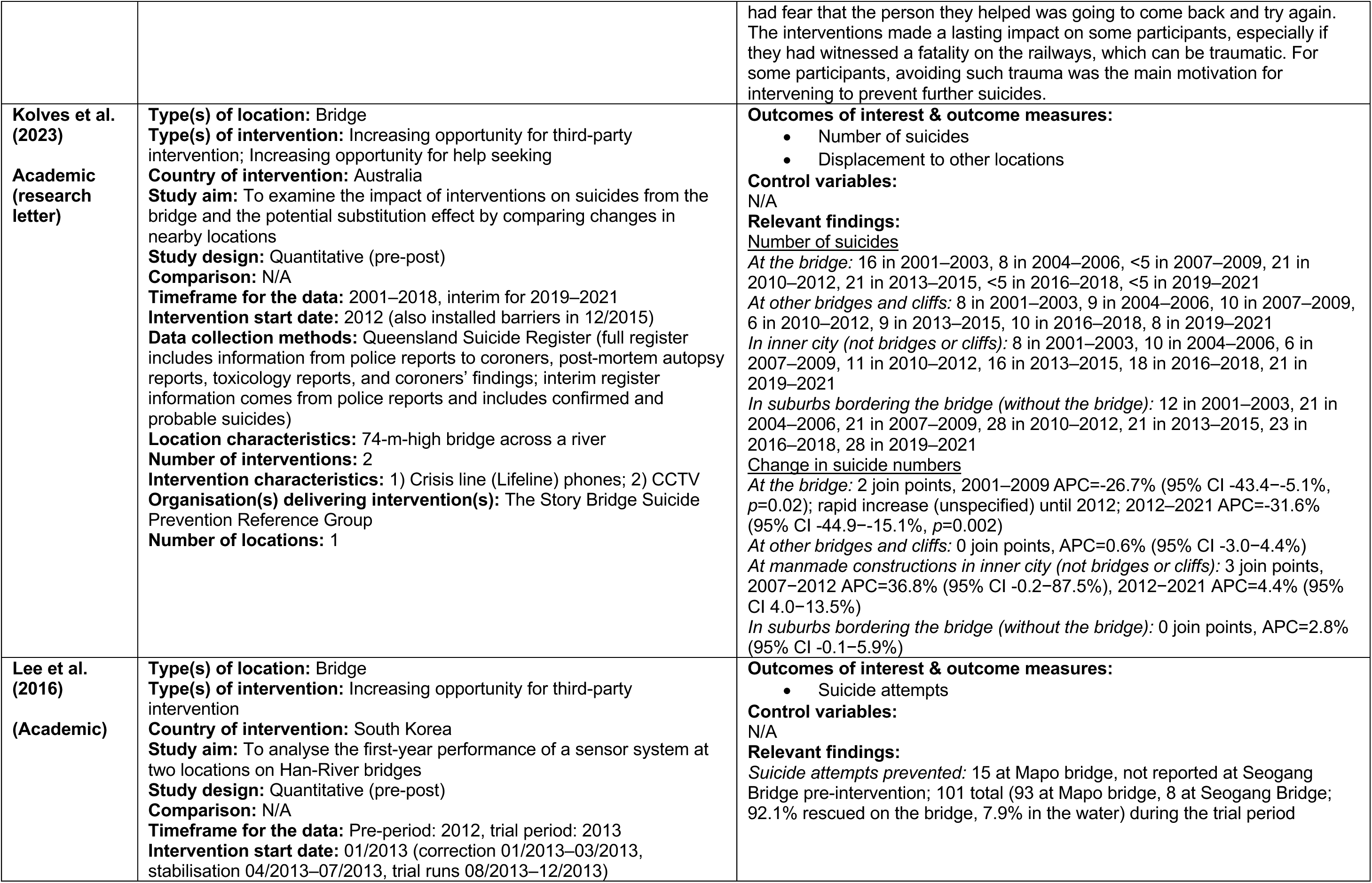
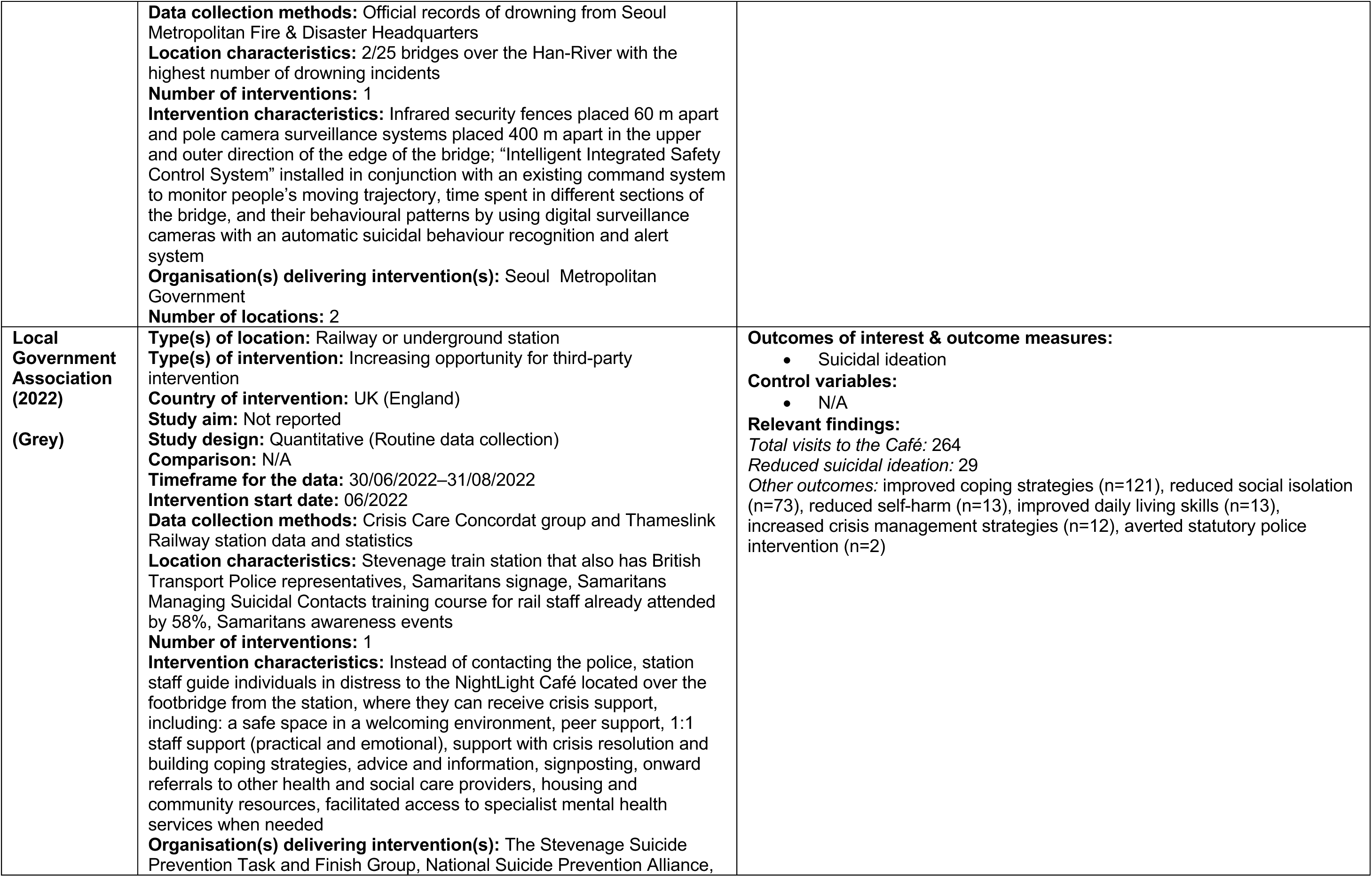
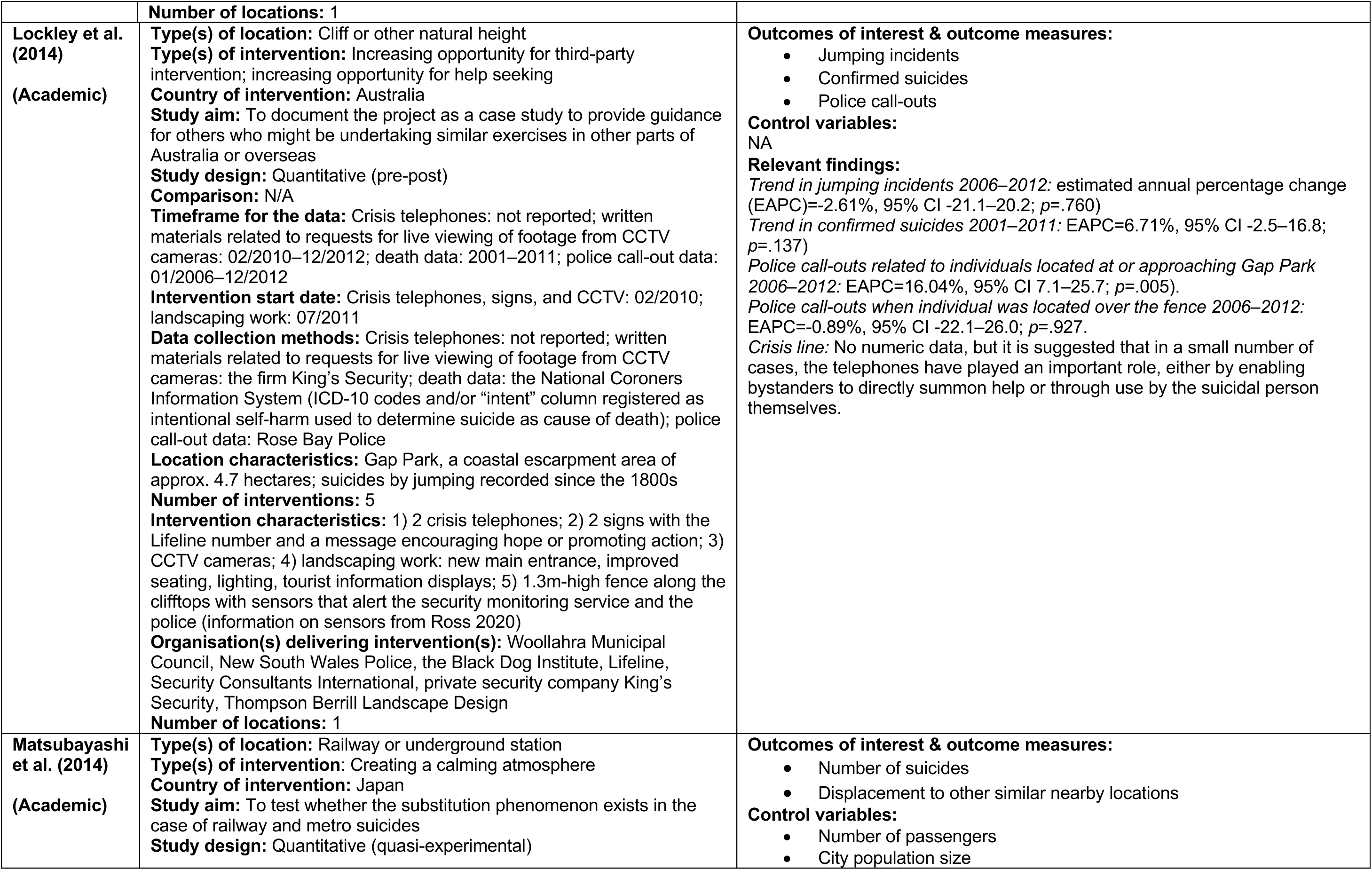
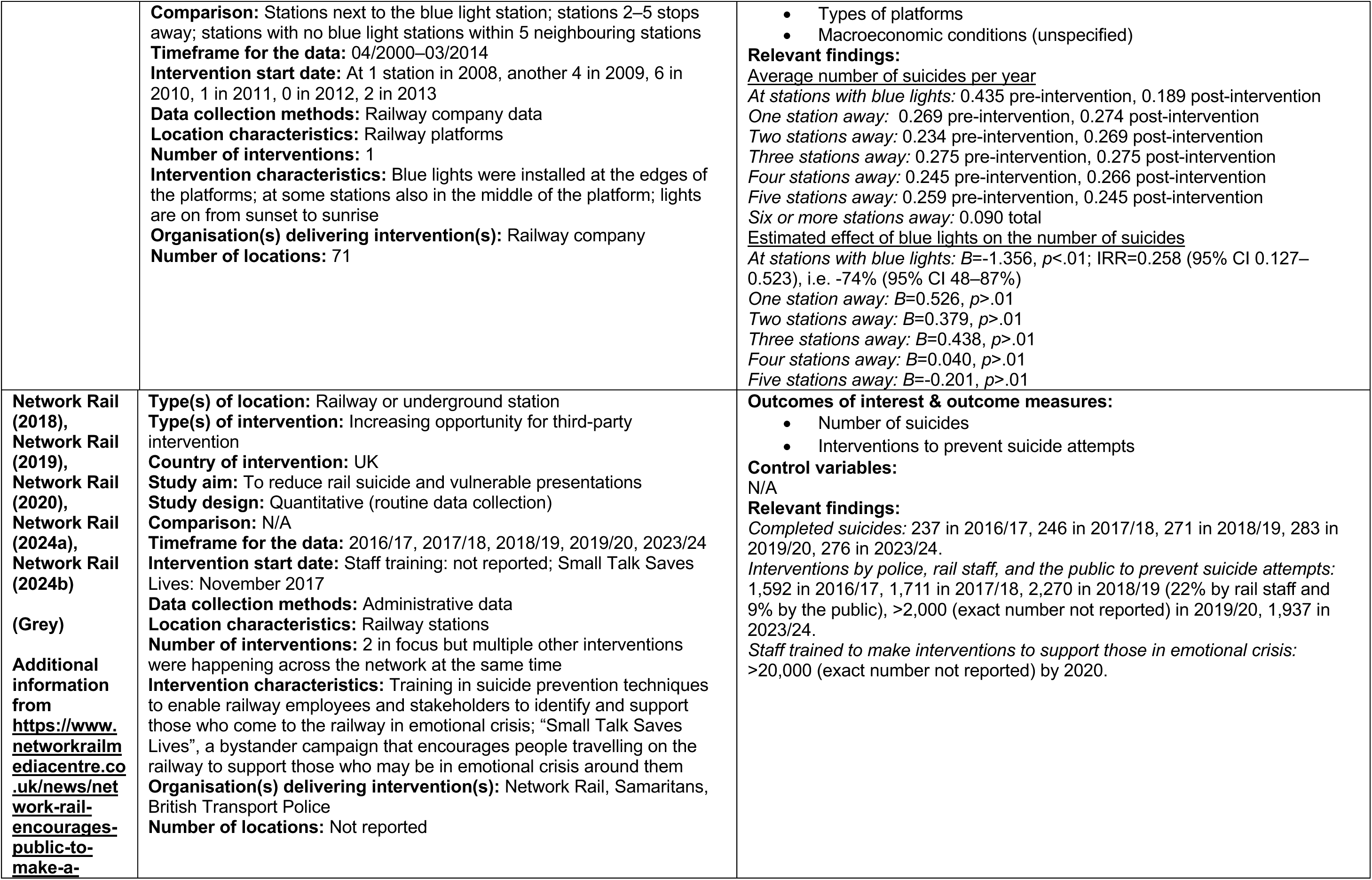
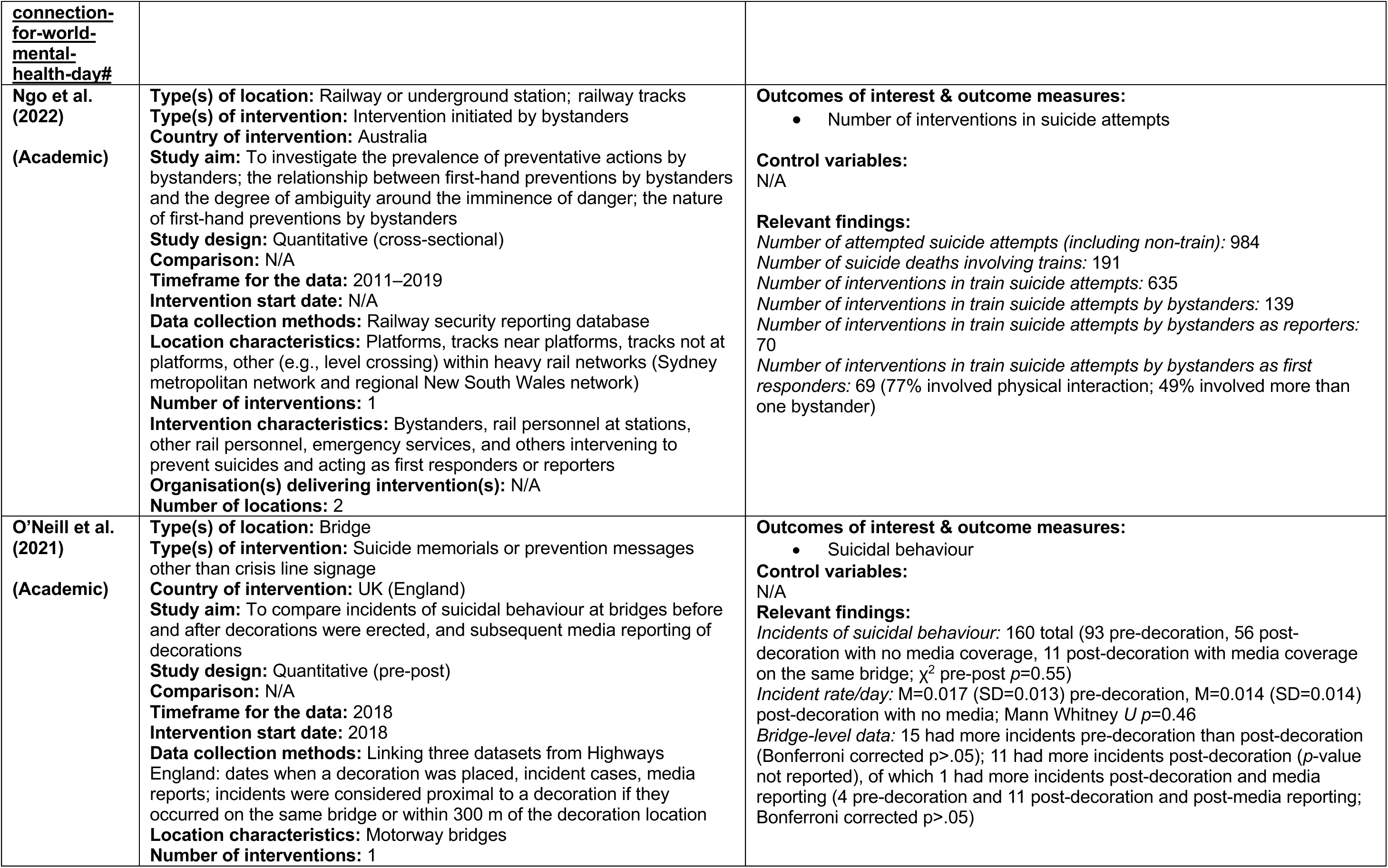
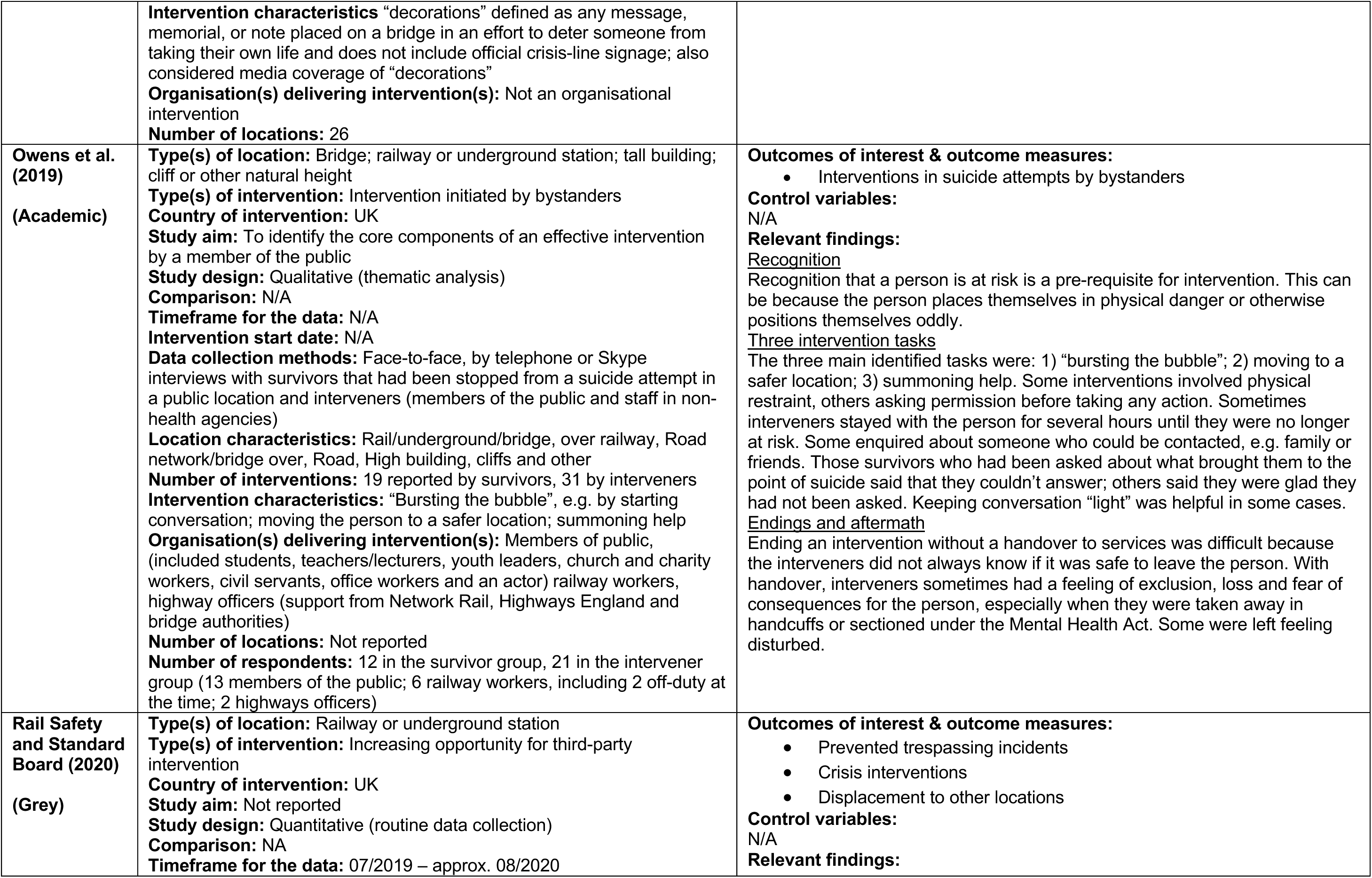
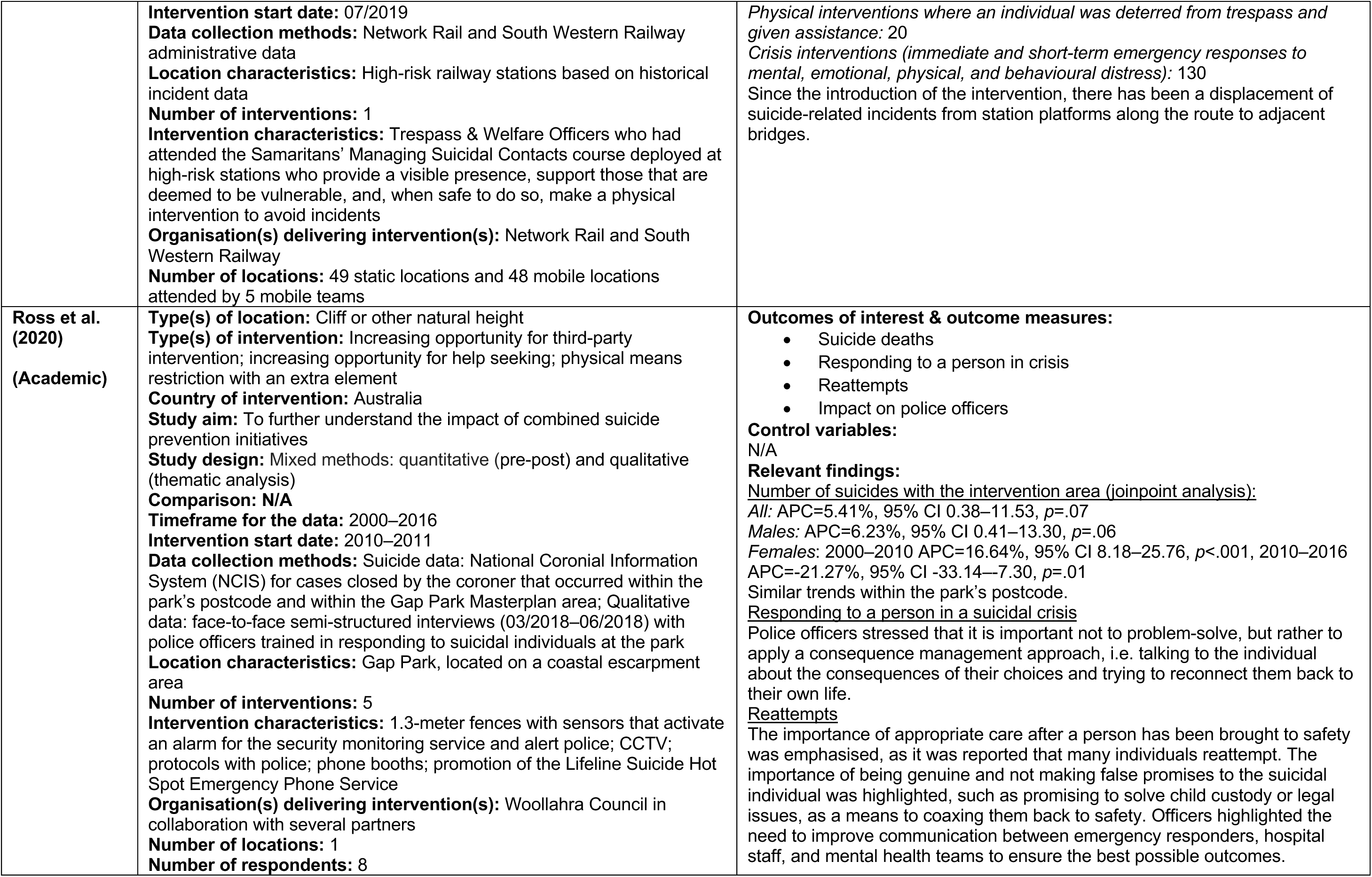
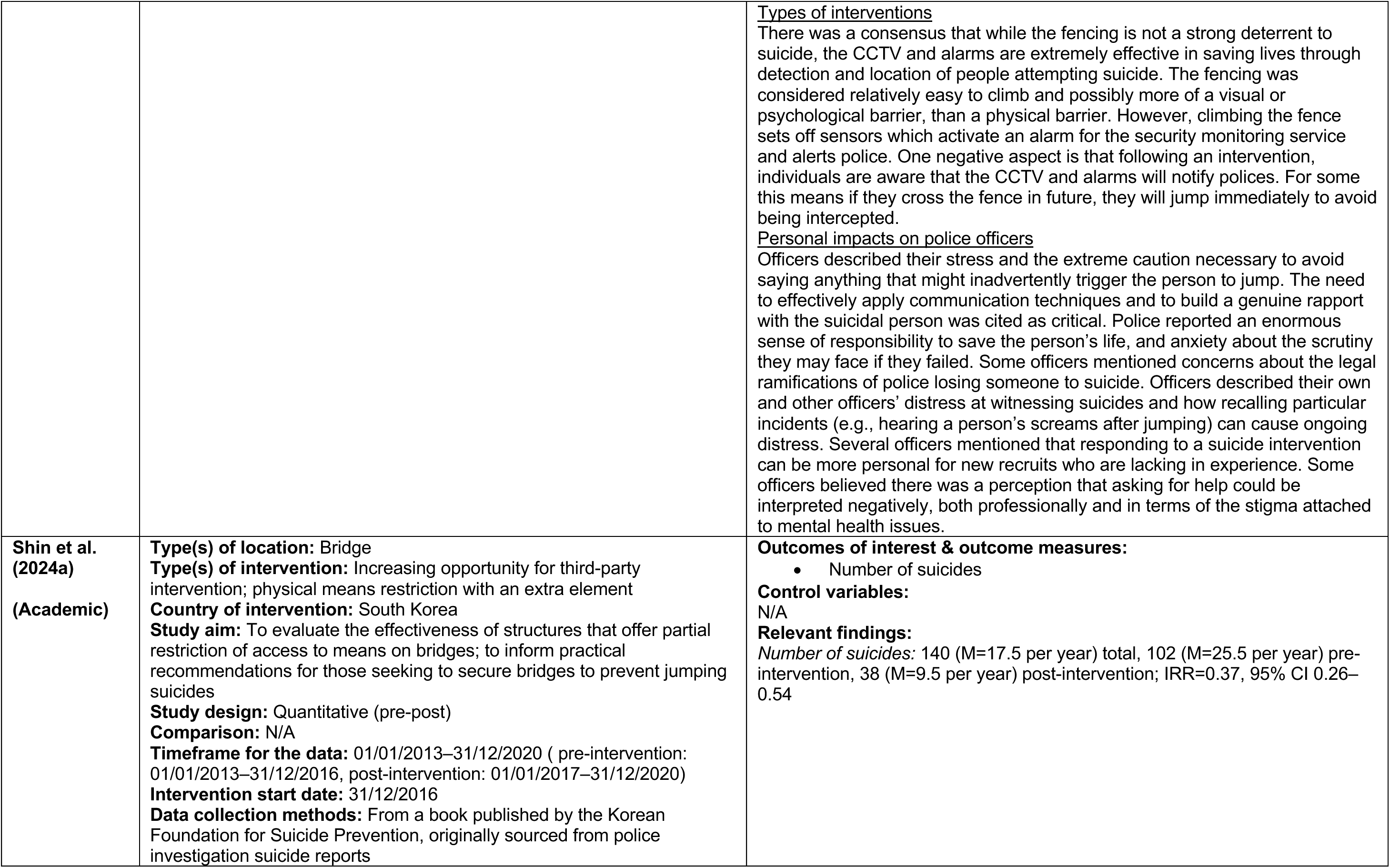
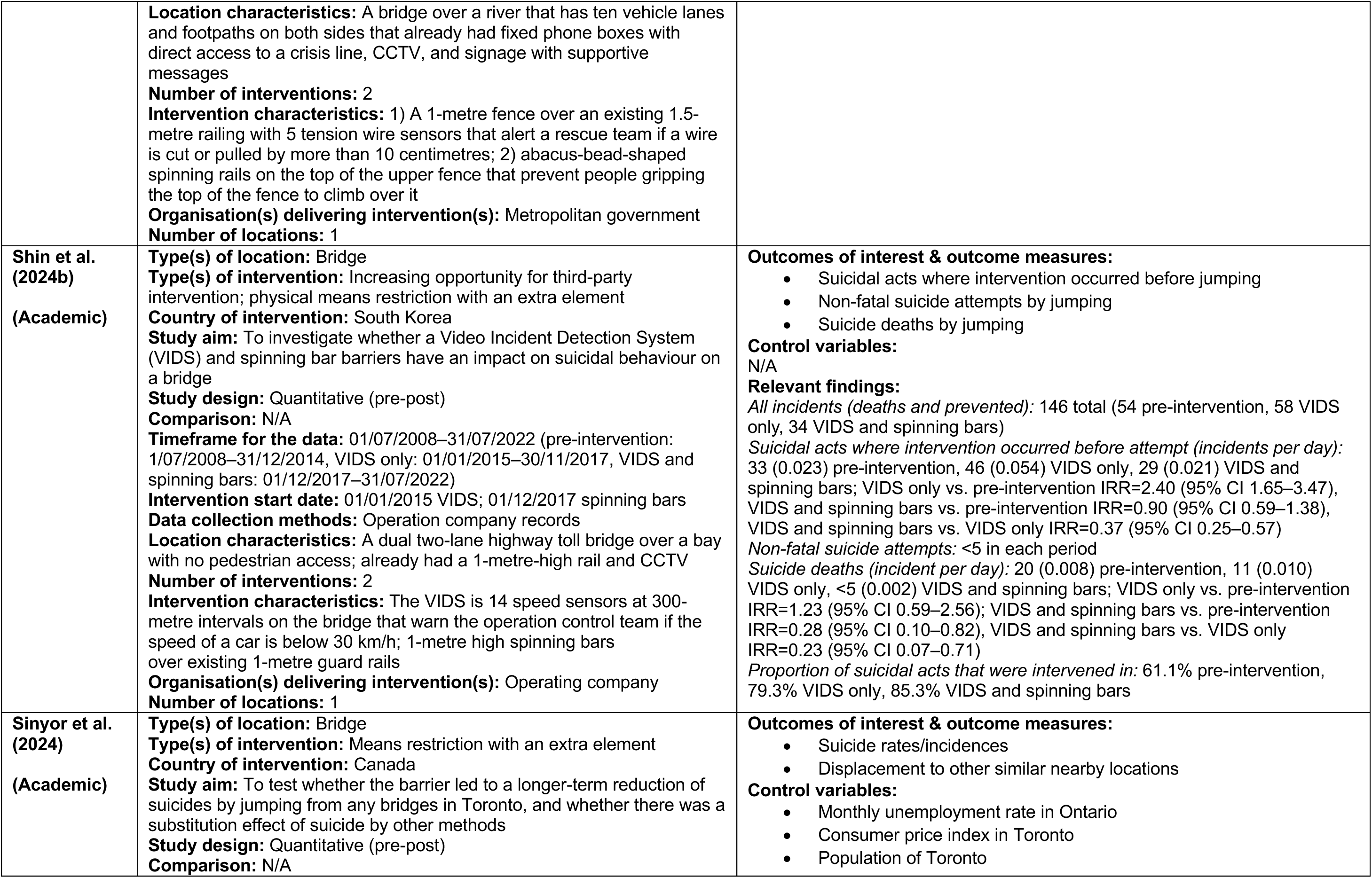
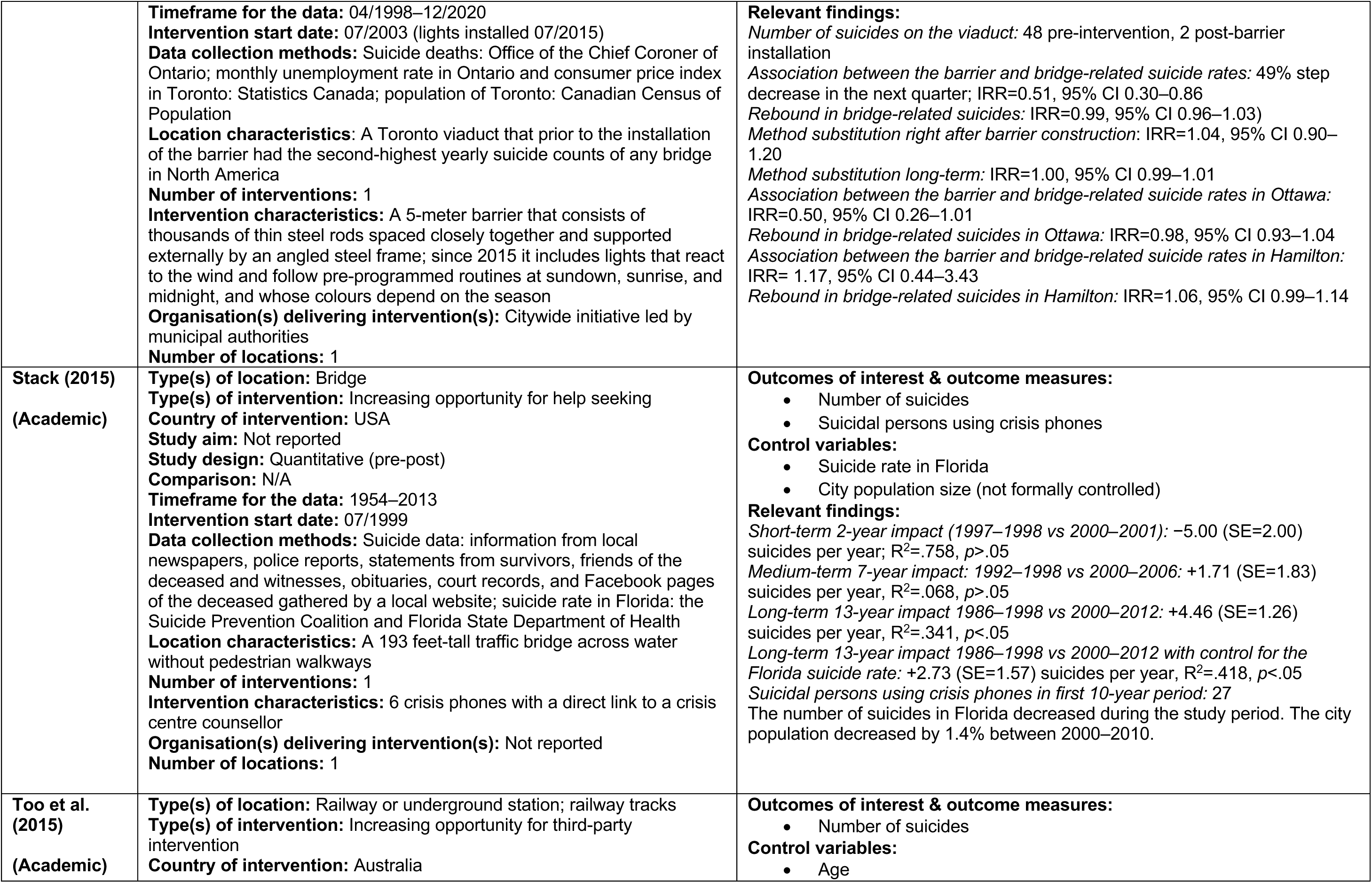
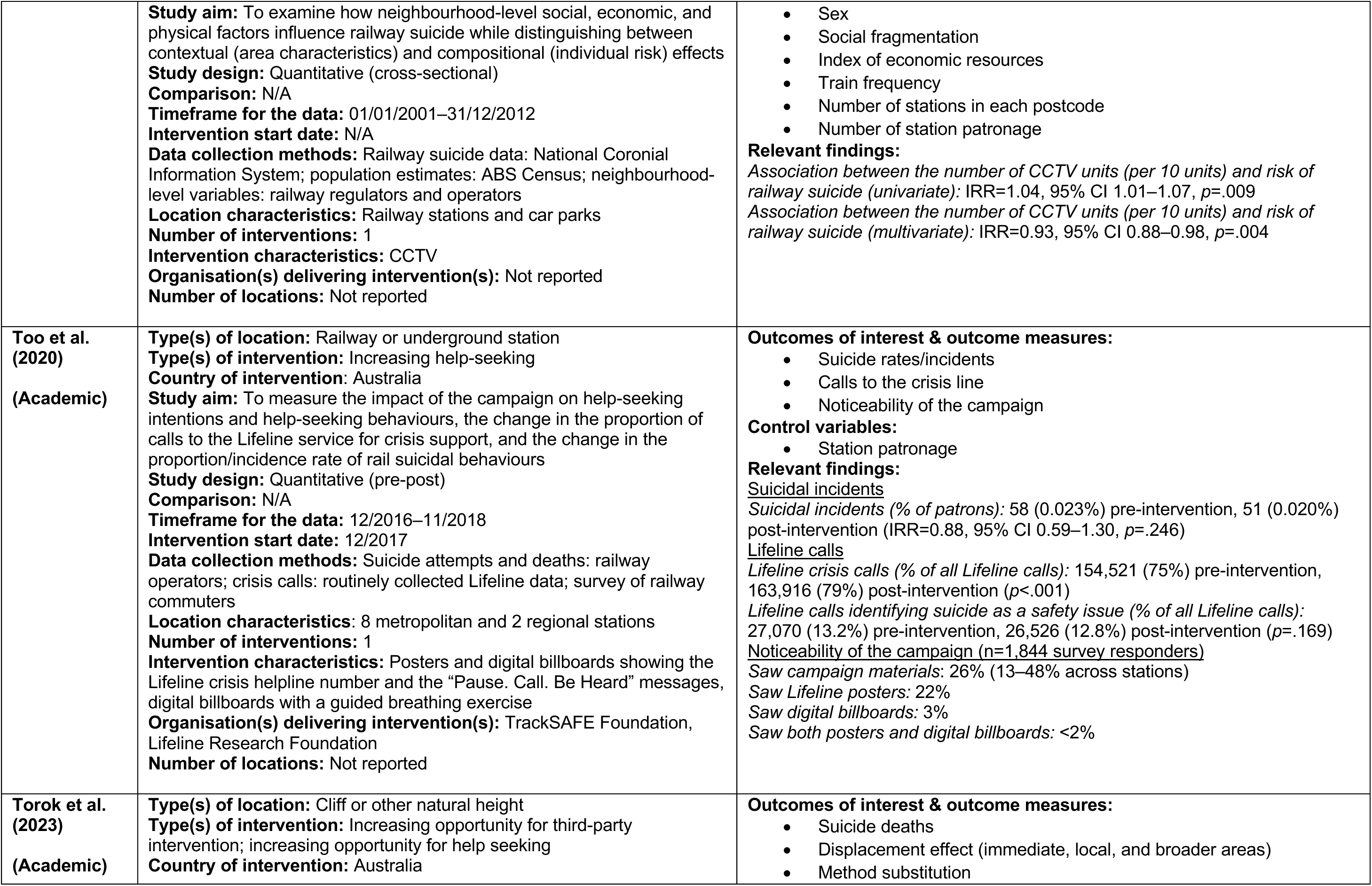
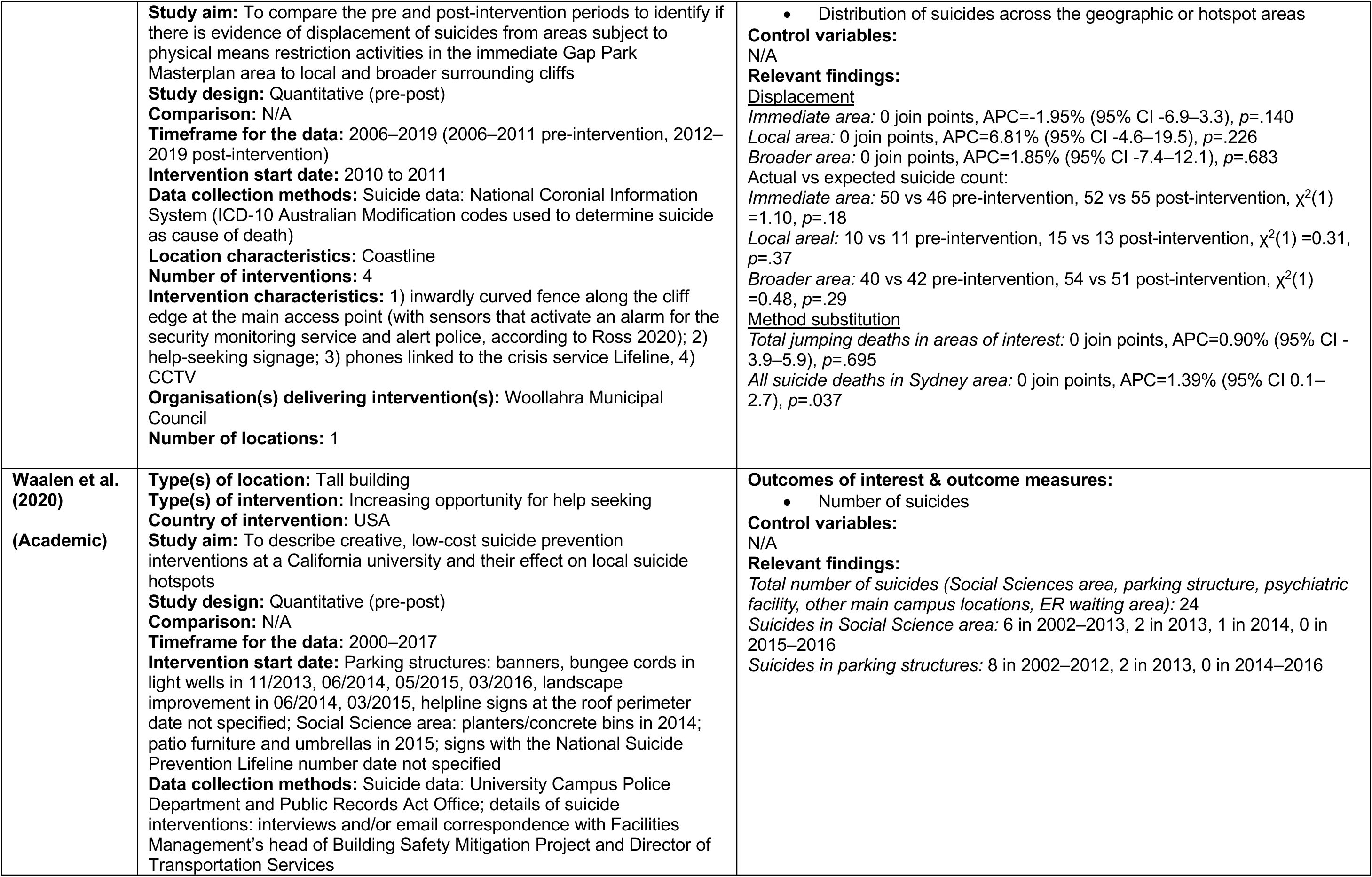
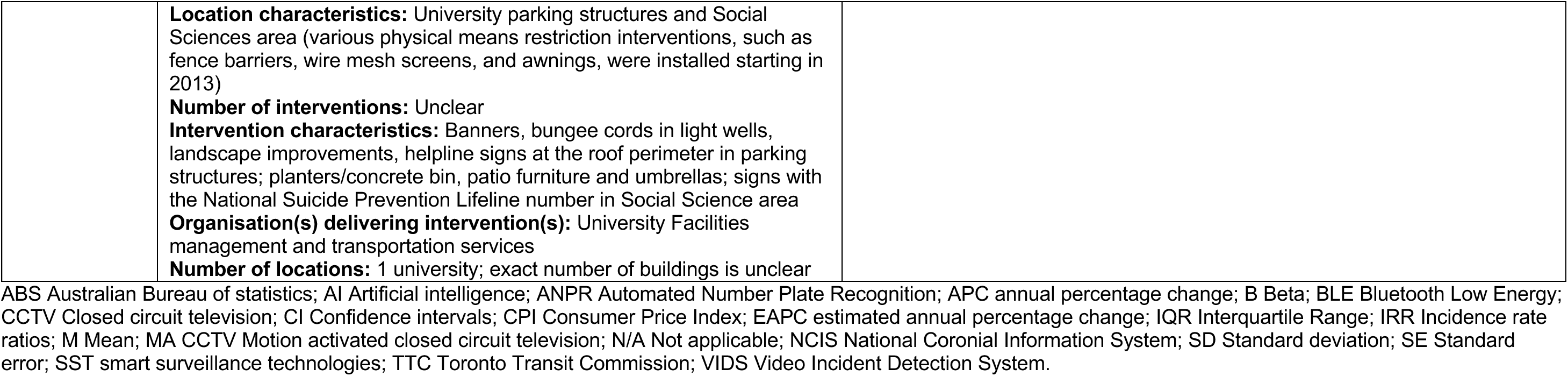

### 6.3 Quality appraisal

#### 6.3.1 Summary of the critical appraisal of the pre-post studies

| **NHLBI Quality Assessment Tool for Before-After (Pre-Post) Studies With No Control Group** | **Chow**<br>**2024** | **Erlangsen**<br>**2023** | **Kolves**<br>**2023** | **Lee**<br>**2016** | **Lockley**<br>**2014** | **O'Neill**<br>**2021** | **Ross**<br>**2020** | **Shin**<br>**2024a** | **Shin**<br>**2024b** | **Sinyor**<br>**2024** | **Stack**<br>**2015** | **Too**<br>**2020** | **Torok**<br>**2023** | **Waalén**<br>**2020** |
| --- | --- | --- | --- | --- | --- | --- | --- | --- | --- | --- | --- | --- | --- | --- |
| 1. Was the study question or objective clearly stated? | Yes | Yes | Yes | Yes | Yes | Yes | Yes | Yes | Yes | Yes | No | Yes | Yes | Yes |
| 2. Were eligibility/selection criteria for the study population prespecified and clearly described? | Yes | N/A | N/A | N/A | N/A | N/A | N/A | N/A | N/A | N/A | Yes | No | N/A | N/A |
| 3. Were the participants in the study representative of those who would be eligible for the test/service/intervention in the general or clinical population of interest? | Yes | CD | CD | Yes | Yes | No | Yes | CD | No | Yes | No | Yes | Yes | Yes |
| 4. Were all eligible participants that met the prespecified entry criteria enrolled? | Yes | N/A | N/A | N/A | N/A | N/A | N/A | N/A | N/A | N/A | N/A | N/A | N/A | N/A |
| 5. Was the sample size sufficiently large to provide confidence in the findings? | CD | No | No | No | No | No | No | Yes | No | Yes | Yes | No | No | No |
| 6. Was the test/service/intervention clearly described and delivered consistently across the study population? | Yes | No | No | Yes | Yes | No | Yes | Yes | Yes | Yes | No | No | No | No |
| 7. Were the outcome measures prespecified, clearly defined, valid, reliable, and assessed consistently across all study participants? | Yes | Yes | Yes | No | Yes | No | Yes | Yes | Yes | Yes | No | No | Yes | Yes |
| 8. Were the people assessing the outcomes blinded to the participants' exposures/interventions? | N/A | N/A | N/A | N/A | N/A | N/A | N/A | N/A | N/A | N/A | N/A | N/A | N/A | N/A |
| 9. Was the loss to follow-up after baseline 20% or less? Were those lost to follow-up accounted for in the analysis? | N/A | N/A | N/A | N/A | N/A | N/A | N/A | N/A | N/A | N/A | N/A | N/A | N/A | N/A |
| 10. Did the statistical methods examine changes in outcome measures from before to after the intervention? Were statistical tests done that provided p values for the pre-to-post changes? | Yes | No | Yes | No | Yes | Yes | Yes | Yes | Yes | Yes | Yes | Yes | Yes | No |
| 11. Were outcome measures of interest taken multiple times before the intervention and multiple times after the intervention (i.e., did they use an interrupted time-series design)? | Yes | No | Yes | No | Yes | No | Yes | No | No | Yes | Yes | No | Yes | No |
| 12. If the intervention was conducted at a group level (e.g., a whole hospital, a community, etc.) did the statistical analysis take into account the use of individual-level data to determine effects at the group level? | N/A | N/A | N/A | N/A | N/A | N/A | N/A | N/A | N/A | N/A | N/A | N/A | N/A | N/A |
| **Overall rating** | **Good** | **Poor** | **Fair** | **Poor** | **Fair** | **Poor** | **Fair** | **Fair** | **Fair** | **Good** | **Poor** | **Poor** | **Fair** | **Poor** |

CD = Cannot be determined; N/A = Not applicable.

#### 6.3.2 Summary of the critical appraisal of the cross-sectional studies

| **JBICritical Appraisal Checklist For Analytical Cross Sectional Studies** | **Joyner 2024a** | **Ngo 2022** | **Too 2015** |
| --- | --- | --- | --- |
| 1. Were the criteria for inclusion in the sample clearly defined? | Yes | Yes | Yes |
| 2. Were the study subjects and the setting described in detail? | Yes | Yes | Yes |
| 3. Was the exposure measured in a valid and reliable way? | N/A | N/A | N/A |
| 4. Were objective, standard criteria used for measurement of the condition? | N/A | N/A | N/A |
| 5. Were confounding factors identified? | Yes | No | Yes |
| 6. Were strategies to deal with confounding factors stated? | No | No | Yes |
| 7. Were the outcomes measured in a valid and reliable way? | Yes | Yes | Yes |
| 8. Was appropriate statistical analysis used? | Yes | Yes | Yes |
| **Overall rating** | **Fair** | **Fair** | **Good** |

#### 6.3.3 Summary of the critical appraisal of the quasi-experimental study

| **JBIChecklist for Quasi-Experimental Studies** | **Matsubayashi 2014** |
| --- | --- |
| 1. Is it clear in the study what is the “cause” and what is the “effect” (i.e. there is no confusion about which variable comes first)? | Yes |
| 2. Was there a control group? | Yes |
| 3. Were participants included in any comparisons similar? | Unclear |
| 4. Were the participants included in any comparisons receiving similar treatment/care, other than the exposure or intervention of interest? | Yes |
| 5. Were there multiple measurements of the outcome, both pre and post the intervention/exposure? | Yes |
| 6. Were the outcomes of participants included in any comparisons measured in the same way? | Yes |
| 7. Were outcomes measured in a reliable way? | No |
| 8. Was follow-up complete and if not, were differences between groups in terms of their follow-up adequately described and analysed? | N/A |
| 9. Was appropriate statistical analysis used? | No |
| **Overall rating** | **Fair** |

#### 6.3.4 Summary of the critical appraisal of the qualitative studies

| **JBICritical Appraisal Checklist for Qualitative Research** | **Joyner 2024a** | **Katsampa 2022** | **Owens 2019** | **Ross 2020** |
| --- | --- | --- | --- | --- |
| 1. Is there congruity between the stated philosophical perspective and the research methodology? | Unclear | Unclear | Unclear | Unclear |
| 2. Is there congruity between the research methodology and the research question or objectives? | Yes | Yes | Yes | Yes |
| 3. Is there congruity between the research methodology and the methods used to collect data? | Yes | Yes | Yes | Yes |
| 4. Is there congruity between the research methodology and the representation and analysis of data? | Yes | Yes | Yes | Yes |
| 5. Is there congruity between the research methodology and the interpretation of results? | Yes | Yes | Yes | Yes |
| 6. Is there a statement locating the researcher culturally or theoretically? | No | No | No | No |
| 7. Is the influence of the researcher on the research, and vice-versa, addressed? | Unclear | No | Unclear | No |
| 8. Are participants, and their voices, adequately represented? | Yes | Yes | Yes | Yes |
| 9. Is the research ethical according to current criteria or, for recent studies, and is there evidence of ethical approval by an appropriate body? | Yes | Yes | Yes | Yes |
| 10. Do the conclusions drawn in the research report flow from the analysis, or interpretation, of the data? | Yes | Yes | Yes | Yes |
| **Overall rating** | **Fair** | **Fair** | **Fair** | **Fair** |

## 7. ADDITIONAL INFORMATION

### 7.1 Information available on request

There is no additional information to be provided on request.

### 7.2 Conflicts of interest

The authors declare they have no conflicts of interest to report.

## Data Availability

All data produced in the present study are available upon reasonable request to the authors

## Acknowledgements

The authors would like to thank the following people for their time, expertise, and contributions during stakeholder meetings, in guiding the focus of the review and presentation of the findings:

- Claire Cotter, National Programme Lead, NHS Wales Executive
- Deborah Job, Regional Lead (North Wales), NHS Wales Executive
- Holly Howe-Davies, Senior Policy Officer for Suicide Prevention and Self-harm, Welsh Government
- Shirley Windsor, Public Mental Health Lead, Public Health Scotland
- Susie Heywood, Suicide Prevention Implementation Support Lead, Public Health Scotland
- Nathan Davies, Health and Care Research Wales Evidence Centre Public Partnership Group member

## ABBREVIATIONS

Abbreviation: Full description
AI: Artificial intelligence
APC: Annual percentage change
CCTV: Closed-circuit television
EAPC: Estimated annual percentage change
GRADE: Grading of Recommendations, Assessment, Development and Evaluation
IMV: Integrated motivational volitional
Med: Median
N/A: Not applicable
RR: Rapid review
VIDS: Video Incident Detection System

## GLOSSARY

Academic literature: is literature published in peer-reviewed academic journals.
Grey literature: is literature published outside of academic journals, for example, reports by government organisations, charities, research institutes etc.
Pre-print: is a version of an academic article that has been published online that has not yet been published in a journal. Pre-prints are usually not peer-reviewed.
Publication bias: is the trend for studies that report positive/statistically significant findings or findings that are perceived to be important to be more likely to be published or published quickly. It can be minimised by searching grey literature.
Qualitative research: is research that uses non-numeric data such as people’s views, for example, findings from interviews or focus groups.
Quantitative research: is research that uses numbers or statistical data.
Confounding factor: is a factor that interferes in the relationship between the intervention and the outcome.

## 8 APPENDIX

## 8.1 APPENDIX 1: Database search strategies

**Ovid MEDLINE(R) ALL <1946 to October 29, 2024>**

| # | Query | Hits |
| --- | --- | --- |
| 1 | suicide/ or suicidal ideation/ or suicide, attempted/ or suicide, completed/ | 68535 |
| 2 | suicid*.ti,ab,kw. | 104831 |
| 3 | ((tak* or took or end*) adj2 (own adj1 (life or lives))).ti,ab,kw. | 272 |
| 4 | (kill* adj1 (hersel* or himsel* or themsel* or onesel*)).ti,ab,kw. | 521 |
| 5 | or/1-4 | 118392 |
| 6 | (building* or multistor* or multi-stor* or skyscraper* or high-rise* or heritage site* or castl*).ti,ab,kw. | 181621 |
| 7 | (flyover* or overpass* or motorway* or highway* or carriageway* or freeway* or road*).ti,ab,kw. | 81899 |
| 8 | (train or trains or traintrack* or rail or railtrack* or railway* or railroad* or metro or underground* or subway* or the tube).ti,ab,kw. | 263725 |
| 9 | (beach* or cliff* or river* or lake* or sea or canal* or reservoir* or viaduct* or coast*).ti,ab,kw. | 497955 |
| 10 | (car park* or carpark* or parking*).ti,ab,kw. | 2269 |
| 11 | bridge*.ti,ab,kw. | 132511 |
| 12 | ((suicide adj3 location*) or hotspot* or hot spot* or high incidence location* or location* of concern).ti,ab,kw. | 51682 |
| 13 | (monument* or tourist site* or woodland* or park or parks or aqueduct* or balcony or balconies or shopping centre* or shopping center* or mall or malls or arcade* or roof* or water* or shore* or quarry or quarries or waste ground or playground* or pier*).ti,ab,kw. | 1184657 |
| 14 | (magnet or iconic or public space* or public place* or frequently used location*).ti,ab,kw. | 24167 |
| 15 | exp Architecture/ | 27736 |
| 16 | Environment Design/ | 7480 |
| 17 | or/6-16 | 2246395 |
| 18 | Suicide Prevention/ | 11915 |
| 19 | Crisis Intervention/ | 6381 |
| 20 | (prevent* or reduc* or address* or manag* or interrup* or restrict* or interven* or lower* or surveil* or barrier* or help* or support* or respon* or rescue* or plan* or infrastructur* or monitor* or agenc* or chang*).ti,ab,kw. | 17689629 |
| 21 | (fenc* or parapet* or net* or pit* or sign* or poster* or helpline* or surveillance* or CCTV* or patrol* or media or reporting* or television* or radio* or mural* or artwork* or message* or technolog* or social media or campaign* or training or access* or planting* or spike* or roller bar* or lighting* or Artificial Intelligence or changing perception* or community awareness).ti,ab,kw. | 13385027 |
| 22 | 18 or 19 or 20 or 21 | 22110594 |
| 23 | 5 and 17 and 22 | 2899 |
| 24 | limit 23 to english language | 2661 |
| 25 | limit 24 to yr="2000 -Current" | 2360 |
| 26 | limit 25 to yr="2014 -Current" | 1650 |

**APA PsycInfo <1806 to October 2024 Week 4>**

| # | Query | Hits |
| --- | --- | --- |
| 1 | exp Attempted Suicide/ or exp Suicide/ or exp Suicidal Ideation/ or exp Suicidal Behavior/ | 49943 |
| 2 | suicid*.tw. | 84111 |
| 3 | ((tak* or took or end*) adj2 (own adj1 (life or lives))).tw. | 357 |
| 4 | (kill* adj1 (hersel* or himsel* or themsel* or onesel*)).tw. | 704 |
| 5 | or/1-4 | 84724 |
| 6 | (building* or multistor* or multi-stor* or skyscraper* or high-rise* or heritage site* or castl*).tw. | 89455 |
| 7 | (flyover* or overpass* or motorway* or highway* or carriageway* or freeway* or road*).tw. | 24382 |
| 8 | (train or trains or traintrack* or rail or railtrack* or railway* or railroad* or metro or underground* or subway* or the tube).tw. | 28617 |
| 9 | (beach* or cliff* or river* or lake* or sea or canal* or reservoir* or viaduct* or coast*).tw. | 26716 |
| 10 | (car park* or carpark* or parking*).tw. | 872 |
| 11 | bridge*.tw. | 24813 |
| 12 | ((suicide adj3 location*) or hotspot* or hot spot* or high incidence location* or location* of concern).tw. | 2045 |
| 13 | (monument* or tourist site* or woodland* or park or parks or aqueduct* or balcony or balconies or shopping centre* or shopping center* or mall or malls or arcade* or roof* or water* or shore* or quarry or quarries or waste ground or playground* or pier*).tw. | 67859 |
| 14 | (magnet or iconic or public space* or public place* or frequently used location*).tw. | 7269 |
| 15 | exp Architecture/ | 2988 |
| 16 | 6 or 7 or 8 or 9 or 10 or 11 or 12 or 13 or 14 or 15 | 260497 |
| 17 | exp Suicide Prevention/ | 7024 |
| 18 | exp Crisis Intervention/ | 11778 |
| 19 | (prevent* or reduc* or address* or manag* or interrup* or restrict* or interven* or lower* or surveil* or barrier* or help* or support* or respon* or rescue* or plan* or infrastrucur* or monitor* or agenc* or chang*).tw. | 3519833 |
| 20 | (fenc* or parapet* or net* or pit* or sign* or poster* or helpline* or surveillance* or CCTV* or patrol* or media or reporting* or television* or radio* or mural* or artwork* or message* or technolog* or social media or campaign* or training or access* or planting* or spike* or roller bar* or lighting* or Artificial Intelligence or changing perception* or community awareness).tw. | 2264035 |
| 21 | 17 or 18 or 19 or 20 | 4177652 |
| 22 | 5 and 16 and 21 | 2528 |
| 23 | limit 22 to english language | 2373 |
| 24 | limit 23 to yr="2014 -Current" | 1319 |

**SCOPUS 30/10/2024**

| # | Query | Hits |
| --- | --- | --- |
|  | ( EXCLUDE ( DOCTYPE , "ch" ) OR EXCLUDE ( DOCTYPE , "bk" ) OR EXCLUDE ( DOCTYPE , "no" ) OR EXCLUDE ( DOCTYPE , "ed" ) OR EXCLUDE ( DOCTYPE , "cr" ) OR EXCLUDE ( DOCTYPE , "le" ) ) | 2,869 |
| 6 | PUBYEAR > 2013 AND PUBYEAR < 2026 | 3,258 |
| 5 | LIMIT-TO ( LANGUAGE , "English" ) | 5,164 |
| 4 | 1 AND 2 AND 3 | 5,628 |
| 3 | TITLE-ABS-KEY ( prevent* OR reduc* OR address* OR manag* OR interrup* OR restrict* OR interven* OR lower* OR surveil* OR barrier* OR help* OR support* OR respon* OR rescue* OR plan* OR infrastructur* OR monitor* OR agenc* OR chang* OR fenc* OR parapet* OR net* OR pit* OR sign* OR poster* OR helpline* OR surveillance* OR CCTV* OR patrol* OR media OR reporting* OR television* OR radio* OR mural* OR artwork* OR message* OR technolog* OR "social media" OR campaign* OR training OR access* OR planting* OR spike* OR "roller bar*" OR lighting* OR "Artificial Intelligence" OR "changing perception*" OR "community awareness" ) | 60,619,984 |
| 2 | TITLE-ABS-KEY ( roof* OR water* OR shore* OR quarry OR quarries OR "waste ground" OR playground* OR pier* OR magnet OR iconic OR "public space*" OR "public place*" OR "frequently used location*" OR building* OR multistor* OR multi-stor* OR skyscraper* OR high-rise* OR "heritage site*" OR castl* OR flyover* OR overpass* OR motorway* OR highway* OR carriageway* OR freeway* OR road* OR train OR trains OR traintrack* OR rail OR railtrack* OR railway* OR railroad* OR metro OR underground* OR subway* OR "the tube" OR beach* OR cliff* OR river* OR lake* OR sea OR canal* OR reservoir* OR viaduct* OR coast* OR "car park*" OR carpark* OR parking* OR bridge* OR ( suicide W/3 location* ) OR hotspot* OR "hot spot*" OR "high incidence location*" OR "location* of concern" OR monument* OR "tourist site*" OR woodland* OR park OR parks OR aqueduct* OR balcony OR balconies OR "shopping centre*" OR "shopping center*" OR mall OR malls OR arcade* ) | 10,885,193 |
| 1 | TITLE-ABS-KEY ( ( ( ( kill* ) W/1 ( hersel* OR himsel* OR themsel* OR onesel* ) ) ) ) OR TITLE-ABS-KEY ( ( tak* OR took OR end* ) W/2 ( own W/1 ( life OR lives ) ) ) OR ( TITLE-ABS-KEY ( suicid* ) ) | 194,208 |

**Proquest: Social Science Database, Sociology Collection 30/10/2024**

| # | Query | Hits |
| --- | --- | --- |
| S1 | title(suicid*) OR abstract(suicid*) | 40,757 |
| S2 | title(kill* NEAR/1 (hersel* OR himsel* OR themsel* OR onesel*)) OR abstract(kill* NEAR/1 (hersel* OR himsel* OR themsel* OR onesel*)) | 444 |
| S3 | title((tak* OR took OR end*) NEAR/1 ("own life" OR "own lives")) OR abstract((tak* OR took OR end*) NEAR/1 ("own life" OR "own lives")) | 240 |
| S4 | [S1] OR [S2] OR [S3] | 40,959 |
| S5 | title(roof* OR water* OR shore* OR quarry OR quarries OR "waste ground" OR playground* OR pier* OR magnet OR iconic OR "public space*" OR "public place*" OR "frequently used location*" OR building* OR multistor* OR multi-stor* OR skyscraper* OR high-rise* OR "heritage site*" OR castl* OR flyover* OR overpass* OR motorway* OR highway* OR carriageway* OR freeway* OR road* OR train OR trains OR traintrack* OR rail OR railtrack* OR railway* OR railroad* OR metro OR underground* OR subway* OR "the tube" OR beach* OR cliff* OR river* OR lake* OR sea OR canal* OR reservoir* OR viaduct* OR coast* OR "car park*" OR carpark* OR parking* OR bridge* OR ( suicide NEAR/3 location* ) OR hotspot* OR "hot spot*" OR "high incidence location*" OR "location* of concern" OR monument* OR "tourist site*" OR woodland* OR park OR parks OR aqueduct* OR balcony OR balconies OR "shopping centre*" OR "shopping center*" OR mall OR malls OR arcade* ) OR abstract(roof* OR water* OR shore* OR quarry OR quarries OR "waste ground" OR playground* OR pier* OR magnet OR iconic OR "public space*" OR "public place*" OR "frequently used location*" OR building* OR multistor* OR multi-stor* OR skyscraper* OR high-rise* OR "heritage site*" OR castl* OR flyover* OR overpass* OR motorway* OR highway* OR carriageway* OR freeway* OR road* OR train OR trains OR traintrack* OR rail OR railtrack* OR railway* OR railroad* OR metro OR underground* OR subway* OR "the tube" OR beach* OR cliff* OR river* OR lake* OR sea OR canal* OR reservoir* OR viaduct* OR coast* OR "car park*" OR carpark* OR parking* OR bridge* OR ( suicide NEAR/3 location* ) OR hotspot* OR "hot spot*" OR "high incidence location*" OR "location* of concern" OR monument* OR "tourist site*" OR woodland* OR park OR parks OR aqueduct* OR balcony OR balconies OR "shopping centre*" OR "shopping center*" OR mall OR malls OR arcade* ) | 324,227 |
| S6 | title(prevent* OR reduc* OR address* OR manag* OR interrup* OR restrict* OR interven* OR lower* OR surveil* OR barrier* OR help* OR support* OR respon* OR rescue* OR plan* OR infrastructur* OR monitor* OR agenc* OR chang* OR fenc* OR parapet* OR net* OR pit* OR sign* OR poster* OR helpline* OR surveillance* OR CCTV* OR patrol* OR media OR reporting* OR television* OR radio* OR mural* OR artwork* OR message* OR technolog* OR "social media" OR campaign* OR training OR access* OR planting* OR spike* OR "roller bar*" OR lighting* OR "Artificial Intelligence" OR "changing perception*" OR "community awareness") OR abstract(prevent* OR reduc* OR address* OR manag* OR interrup* OR restrict* OR interven* OR lower* OR surveil* OR barrier* OR help* OR support* OR respon* OR rescue* OR plan* OR infrastructur* OR monitor* OR agenc* OR chang* OR fenc* OR parapet* OR net* OR pit* OR sign* OR poster* OR helpline* OR surveillance* OR CCTV* OR patrol* OR media OR reporting* OR television* OR radio* OR mural* OR artwork* OR message* OR technolog* OR "social media" OR campaign* OR training OR "human intervention*" OR access* OR planting* OR spike* OR "roller bar*" OR lighting* OR "Artificial Intelligence" OR "changing perception*" OR "community awareness") | 2,957,665 |
| S7 | [S4] AND [S5] AND [S6] | 974 |
| S8 | [S4] AND [S5] AND [S6] Limited to English language | 943 |
| S9 | [S4] AND [S5] AND [S6] Limited to publication date (20140101-20241030) | 438 |

**Cochrane 30/10/2024**

| # | Query | Hits |
| --- | --- | --- |
| #1 | MeSH descriptor: [Suicide] explode all trees | 2212 |
| #2 | (suicid*):ti,ab,kw | 9090 |
| #3 | (kill* NEAR/1 (hersel* or himsel* or themsel* or onesel*)):ti,ab,kw | 13 |
| #4 | ((tak* or took or end*) NEAR/2 (own NEAR/1 (life or lives))):ti,ab,kw | 11 |
| #5 | #1 OR #2 OR #3 OR #4 | 9092 |
| #6 | (roof* OR water* OR shore* OR quarry OR quarries OR "waste ground" OR playground* OR pier* OR magnet OR iconic OR building* OR multistor* OR multi stor* OR skyscraper* OR high rise* OR castl* OR flyover* OR overpass* OR motorway* OR highway* OR carriageway* OR freeway* OR road* OR train OR trains OR traintrack* OR rail OR railtrack* OR railway* OR railroad* OR metro OR underground* OR subway* OR "the tube" OR beach* OR cliff* OR river* OR lake* OR sea OR canal* OR reservoir* OR viaduct* OR coast* OR carpark* OR parking* OR bridge* OR hotspot OR monument* OR woodland* OR park OR parks OR aqueduct* OR balcony OR balconies OR mall OR malls OR arcade*):ti,ab,kw | 91128 |
| #7 | (Public NEXT space*):ti,ab,kw | 101 |
| #8 | (frequently NEXT used NEXT location*):ti,ab,kw | 0 |
| #9 | (heritage NEXT site*):ti,ab,kw | 0 |
| #10 | (car NEXT park*):ti,ab,kw | 6 |
| #11 | (hot NEXT spot*):ti,ab,kw | 258 |
| #12 | (high NEXT incidence NEXT location*):ti,ab,kw | 0 |
| #13 | (location* NEXT of NEXT concern):ti,ab,kw | 0 |
| #14 | (Tourist NEXT site*):ti,ab,kw | 1 |
| #15 | (shopping NEXT centre*):ti,ab,kw | 7 |
| #16 | (shopping NEXT center*):ti,ab,kw | 11 |
| #17 | (suicide NEAR/2 location OR suicide NEAR/2 locations):ti,ab,kw | 0 |
| #18 | OR 6-16 | 91424 |
| #19 | (roller NEXT bar*):ti,ab,kw | 3 |
| #20 | (changing NEXT perception*):ti,ab,kw | 38 |
| #21 | (prevent* OR reduc* OR address* OR manag* OR interrupt* OR restrict* OR interven* OR lower* OR surveil* OR barrier* OR help* OR support* OR respon* OR rescue* OR plan* OR infrastructur* OR monitor* OR agenc* OR chang* OR fenc* OR parapet* OR net* OR pit* OR sign* OR poster* OR helpline* OR surveillance* OR CCTV* OR patrol* OR media OR reporting* OR television* OR radio* OR mural* OR artwork* OR message* OR technolog* OR "social media" OR campaign* OR training OR access* OR planting* OR spike* OR lighting* OR "artificial intelligence" OR "community awareness"):ti,ab,kw | 1799367 |
| #22 | OR #19-#21 | 1799367 |
| #23 | #5 AND #18 AND #22 | 423 |
| #24 | #5 AND #18 AND #22 with Cochrane Library publication date Between Jan 2014 and Nov 2024 | 407 |

N.B. One is an editorial that is not relevant and wasn't exported

**Overton 27/11/2024**

(suicid* AND (“public place” OR “public places” OR “public space” OR “public spaces” OR “public location” OR “public locations” OR “high incidence location” OR “high incidence locations”)) OR (“suicide location” OR “suicide locations” OR “location of concern” OR “locations of concern” OR “location suicide”∼3 OR “location suicides”∼3 OR “locations suicide”∼3 OR “locations suicides”∼3)

Published since 2014. Location: UK. Sorted by relevance.

## 8.2 APPENDIX 2: Searched websites

- All On Board https://allonboard.org.uk/
- British Transport Police https://www.btp.police.uk/
- Cadw https://cadw.gov.wales/
- Campaign Against Living Miserably (CALM) https://www.thecalmzone.net/
- Chasing the stigma https://www.chasingthestigma.co.uk/
- Harmless (The centre of Excellence for self-harm and suicide prevention) https://harmless.org.uk/
- Health and Safety Executive (Ireland) https://www.hse.ie
- Highways England https://highwaysengland.co.uk/
- Historic England https://historicengland.org.uk/
- Historic Environment Scotland https://www.historicenvironment.scot/
- Historic Royal Palaces http://www.hrp.org.uk/
- International Institute for Environment and Development https://www.iied.org/
- Joseph Rowntree Foundation https://www.jrf.org.uk/
- Local Government Association https://www.local.gov.uk/
- Mental Health Foundation https://www.mentalhealth.org.uk/
- MIND https://harmless.org.uk/
- National Centre for Social Research https://natcen.ac.uk/
- National Institute of Economic and Social Research https://niesr.ac.uk/
- National Suicide Prevention Alliance https://nspa.org.uk/
- Network Rail https://www.networkrail.co.uk/
- New Local https://www.newlocal.org.uk/
- NICE http://www.nice.org.uk/
- Nuffield Foundation https://www.nuffieldfoundation.org/
- Office of Health Economics https://www.ohe.org/
- Papyrus (prevention of young suicide) https://www.papyrus-uk.org/
- Public Health Agency (Northern Ireland) https://www.publichealth.hscni.net/
- Public Health Intervention Responsive Studies Teams (PHIRST) https://phirst.nihr.ac.uk/
- Public Health Scotland https://publichealthscotland.scot/
- Public Health Wales https://phw.nhs.wales/
- Rail Safety and Standards Board https://www.rssb.co.uk/
- Rail Suicide Prevention https://railsuicideprevention.co.uk/
- RESTRAIL https://www.restrail.eu/
- RAND https://www.rand.org/
- Royal College of Psychiatrists https://www.rcpsych.ac.uk/
- Royal Life Saving Society https://www.rlss.org.uk/
- Samaritans https://www.samaritans.org/
- Suicide prevention UK https://www.spuk.org.uk
- Tavistock Institute of Human Relations https://www.tavinstitute.org/
- The British Psychological Society (BPS) https://www.bps.org.uk/
- TRL https://www.trl.co.uk/
- UK government https://www.gov.uk/
- University of Glasgow Suicide Behaviour Research Laboratory https://suicideresearch.info/
- Wales Centre for Public Policy https://www.wcpp.org.uk/
- Water Safety Scotland https://www.watersafetyscotland.org.uk/
- Welsh Government https://www.gov.wales/
- What Works Wellbeing https://whatworkswellbeing.org/

## 8.3 APPENDIX 3: Identified systematic reviews

- Barker E, Kolves K, De Leo D. (2017). Rail-suicide prevention: Systematic literature review of evidence-based activities. Asia Pac Psychiatry. 9(3). doi: 10.1111/appy.12246
- Chamberlain B, Woodnutt S. (2024). Preventing suicide by jumping in public locations: a systematic review of interventions. Mental Health Practice. 27(3): 24-30. doi: 10.7748/mhp.2024.e1681
- Grabušić S, Barić D. (2023). A Systematic Review of Railway Trespassing: Problems and Prevention Measures. Sustainability (Switzerland). 15(18). doi: 10.3390/su151813878
- Havârneanu GM, Burkhardt JM, Paran F. (2015). A systematic review of the literature on safety measures to prevent railway suicides and trespassing accidents. Accid Anal Prev. 81: 30-50. doi: 10.1016/j.aap.2015.04.012
- Hoffberg AS, Stearns-Yoder KA, Brenner LA. (2020). The Effectiveness of Crisis Line Services: A Systematic Review. Frontiers in Public Health. 7. doi: 10.3389/fpubh.2019.00399
- Mishara BL, Bardon C. (2016). Systematic review of research on railway and urban transit system suicides. Journal of affective disorders. 193: 215-26. doi: https://10.1016/j.jad.2015.12.042
- Niederkrotenthaler T, Braun M, Pirkis J, et al. (2020). Association between suicide reporting in the media and suicide: Systematic review and meta-analysis. The BMJ. 368. doi: 10.1136/bmj.m575
- Okolie C, Hawton K, Lloyd K, et al. (2020a). Means restriction for the prevention of suicide on roads. Cochrane Database Syst Rev. 9(9): CD013738. doi: 10.1002/14651858.CD013738
- Okolie C, Wood S, Hawton K, et al. (2020b). Means restriction for the prevention of suicide by jumping. Cochrane Database Syst Rev. 2(2): CD013543. doi: 10.1002/14651858.CD013543
- Pirkis J, Too LS, Spittal MJ, et al. (2015). Interventions to reduce suicides at suicide hotspots: a systematic review and meta-analysis. Lancet Psychiatry. 2(11): 994-1001. doi: 10.1016/S2215-0366(15)00266-7
- Public Health Scotland. (2022). Rapid literature review in reducing suicides at locations of concern.
- Radun I, Kannan P, Partonen T, et al. (2024). A systematic review of road traffic suicides: Do we know enough to propose effective preventive measures? Transportation Research Part F: Traffic Psychology and Behaviour. 106: 14-26. doi: 10.1016/j.trf.2024.07.028
- Too LS, Milner A, Bugeja L, et al. (2014). The socio-environmental determinants of railway suicide: a systematic review. BMC public health. 14: 20. doi: https://10.1186/1471-2458-14-20
- Zalsman G, Hawton K, Wasserman D, et al. (2016). Suicide prevention strategies revisited: 10-year systematic review. The lancet. Psychiatry. 3(7): 646-59. doi: https://10.1016/S2215-0366(16)30030-X

## 8.4 APPENDIX 4: Studies excluded at full-text screening

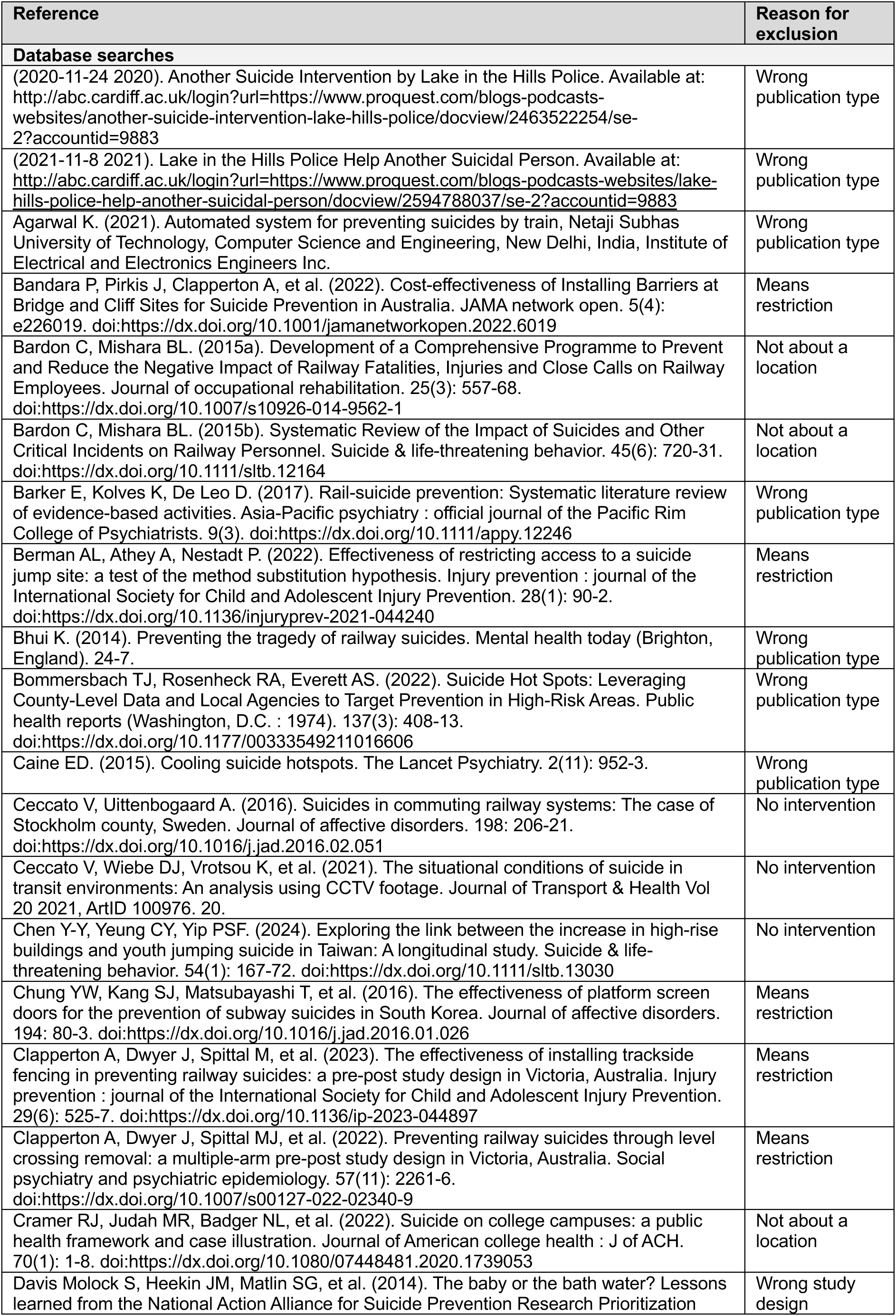
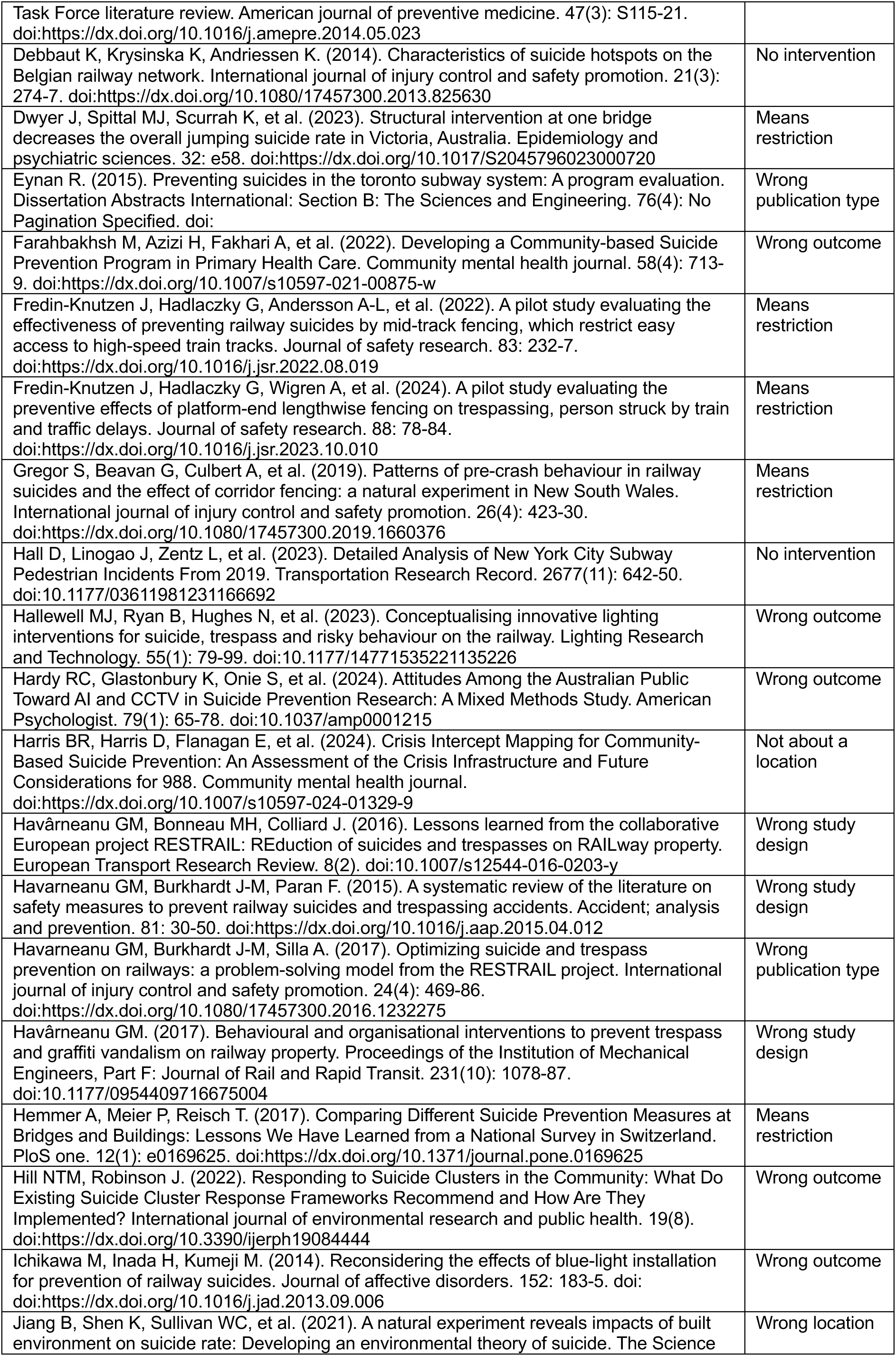
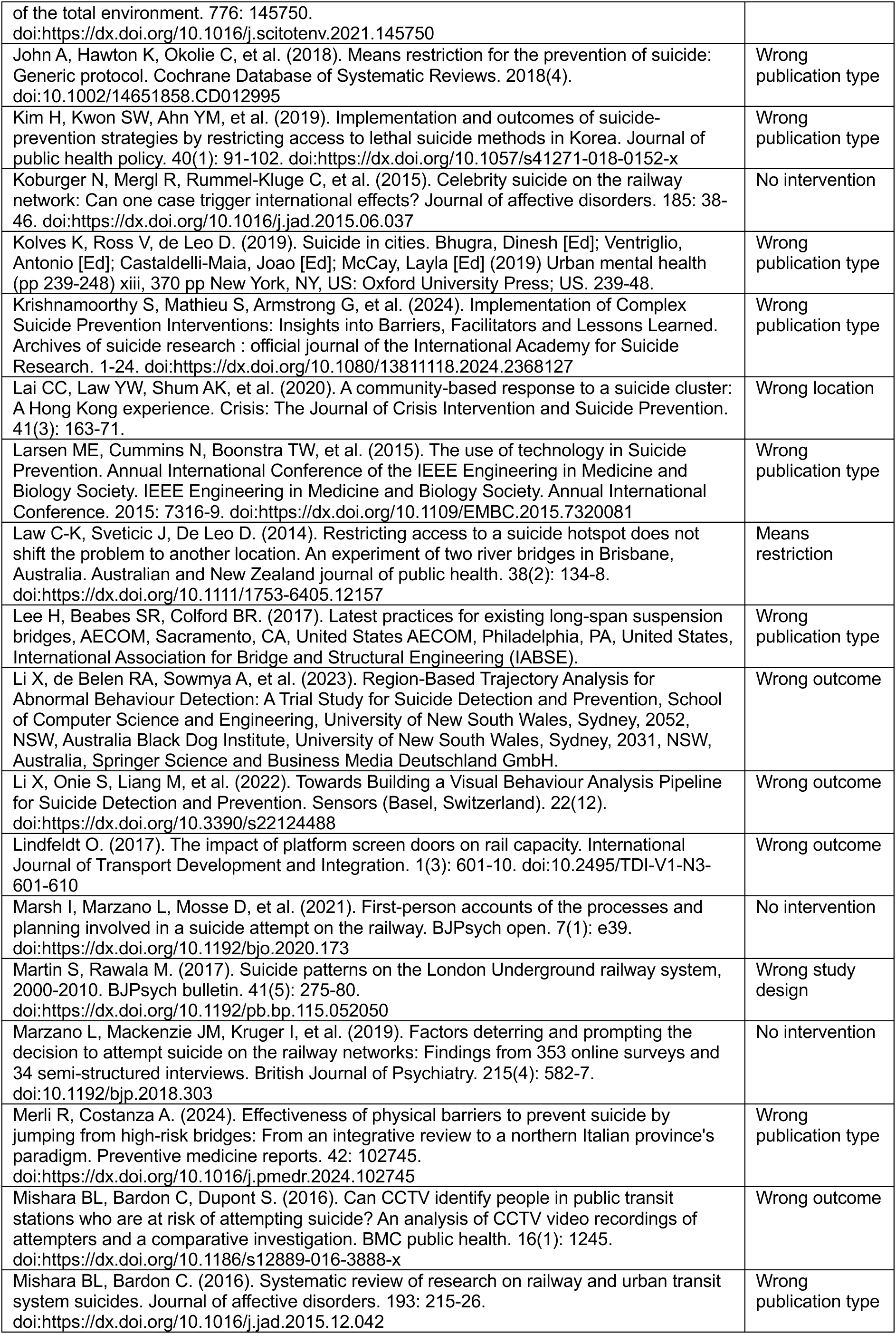
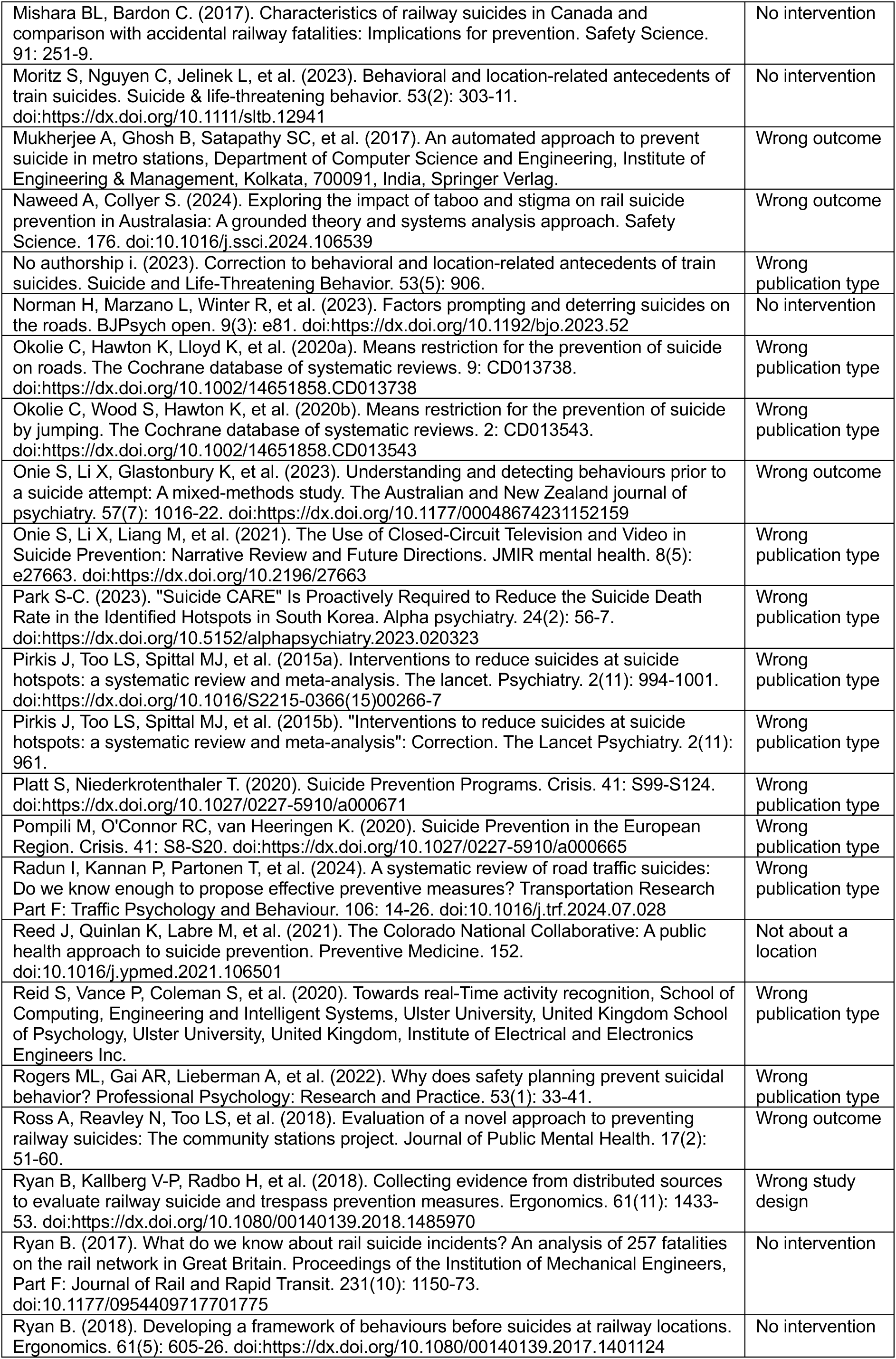
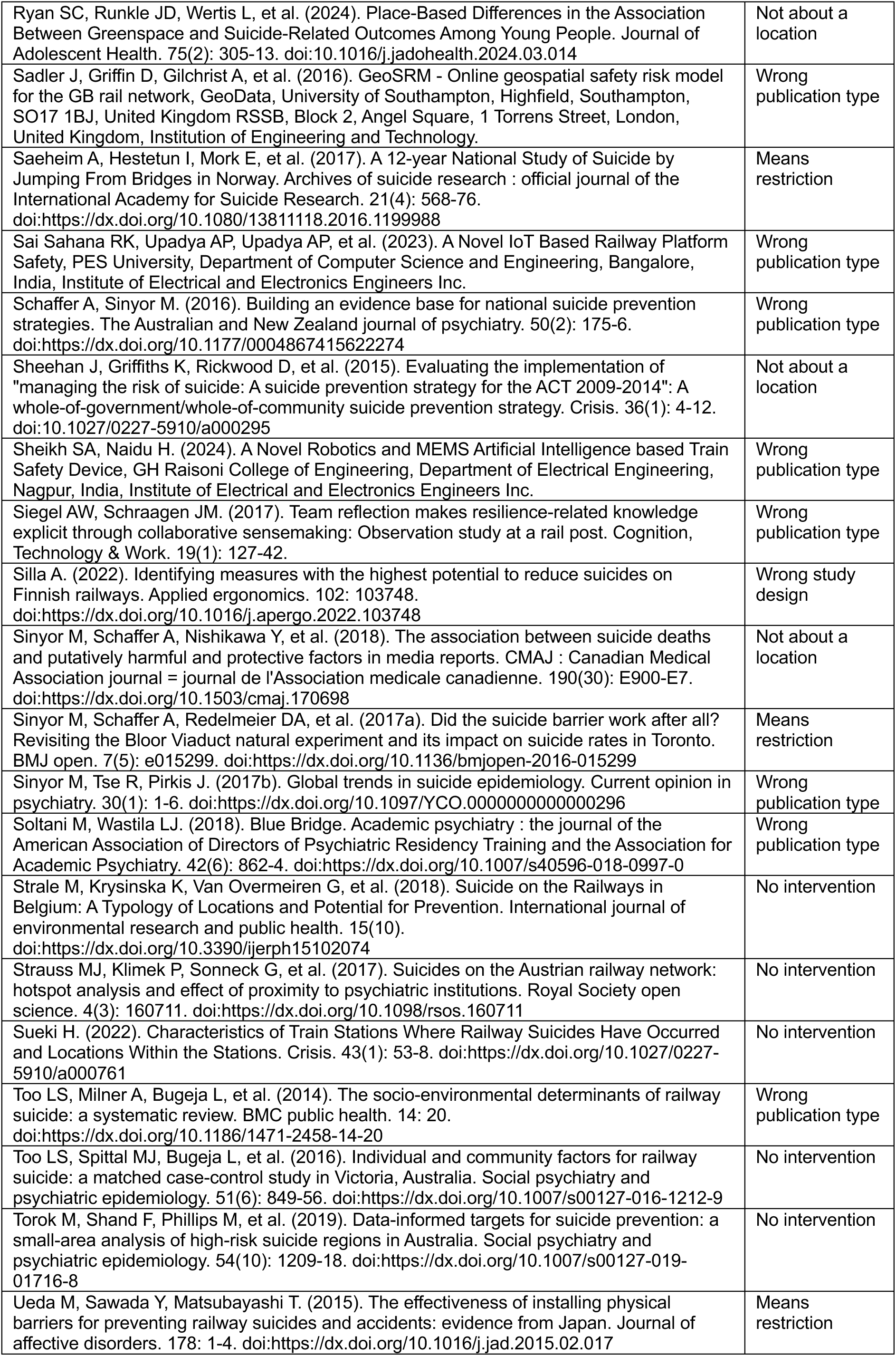
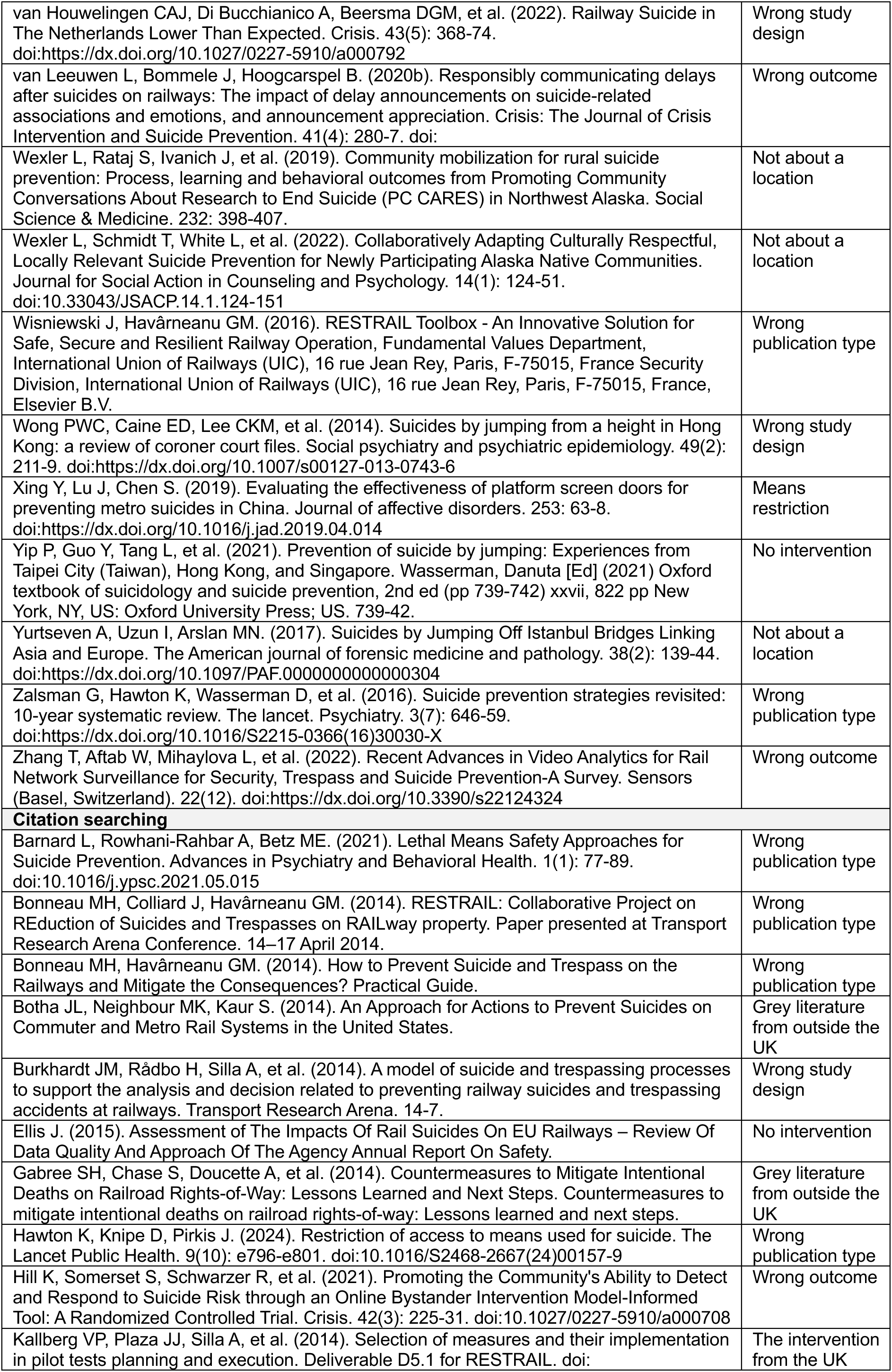
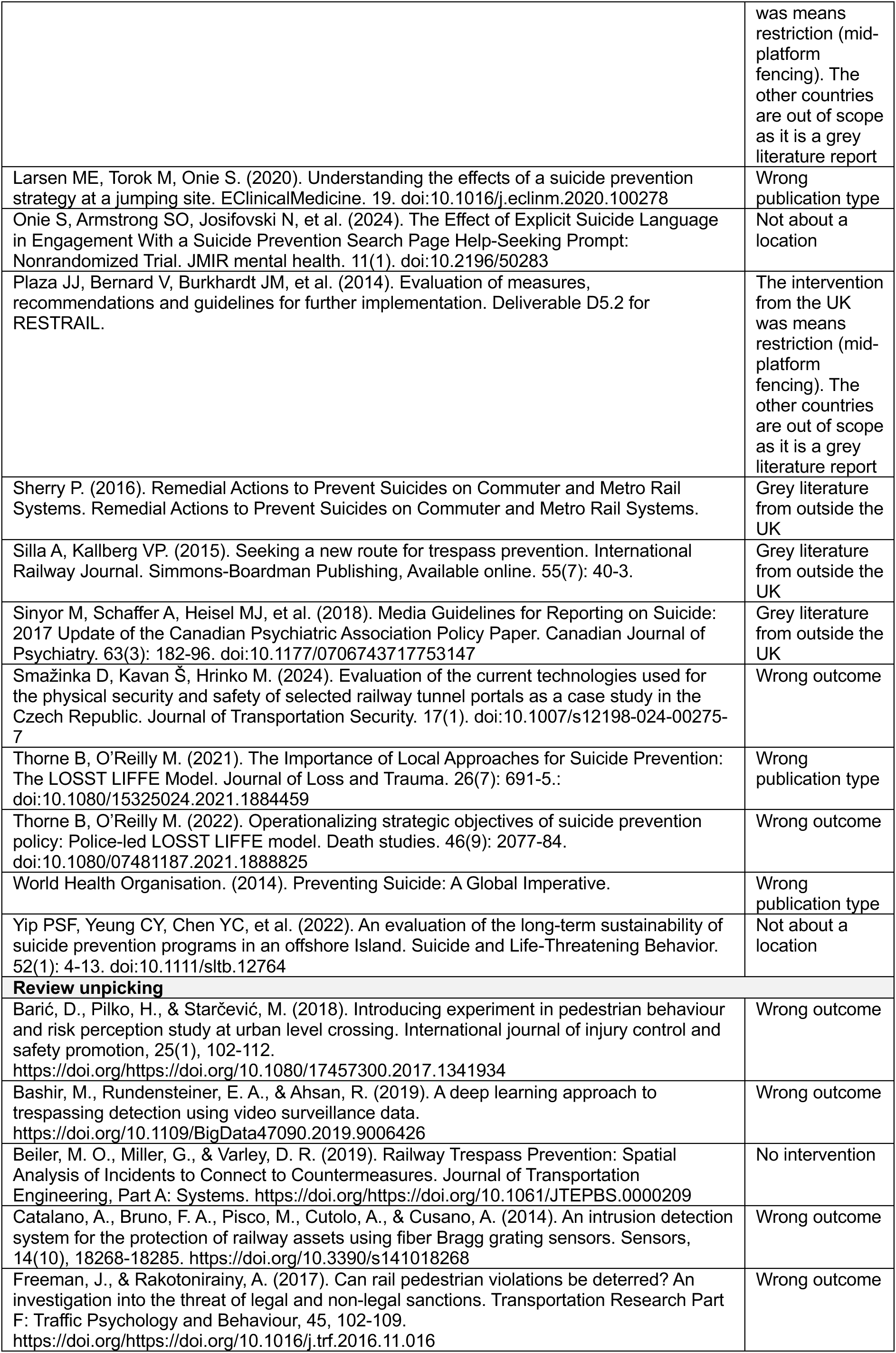
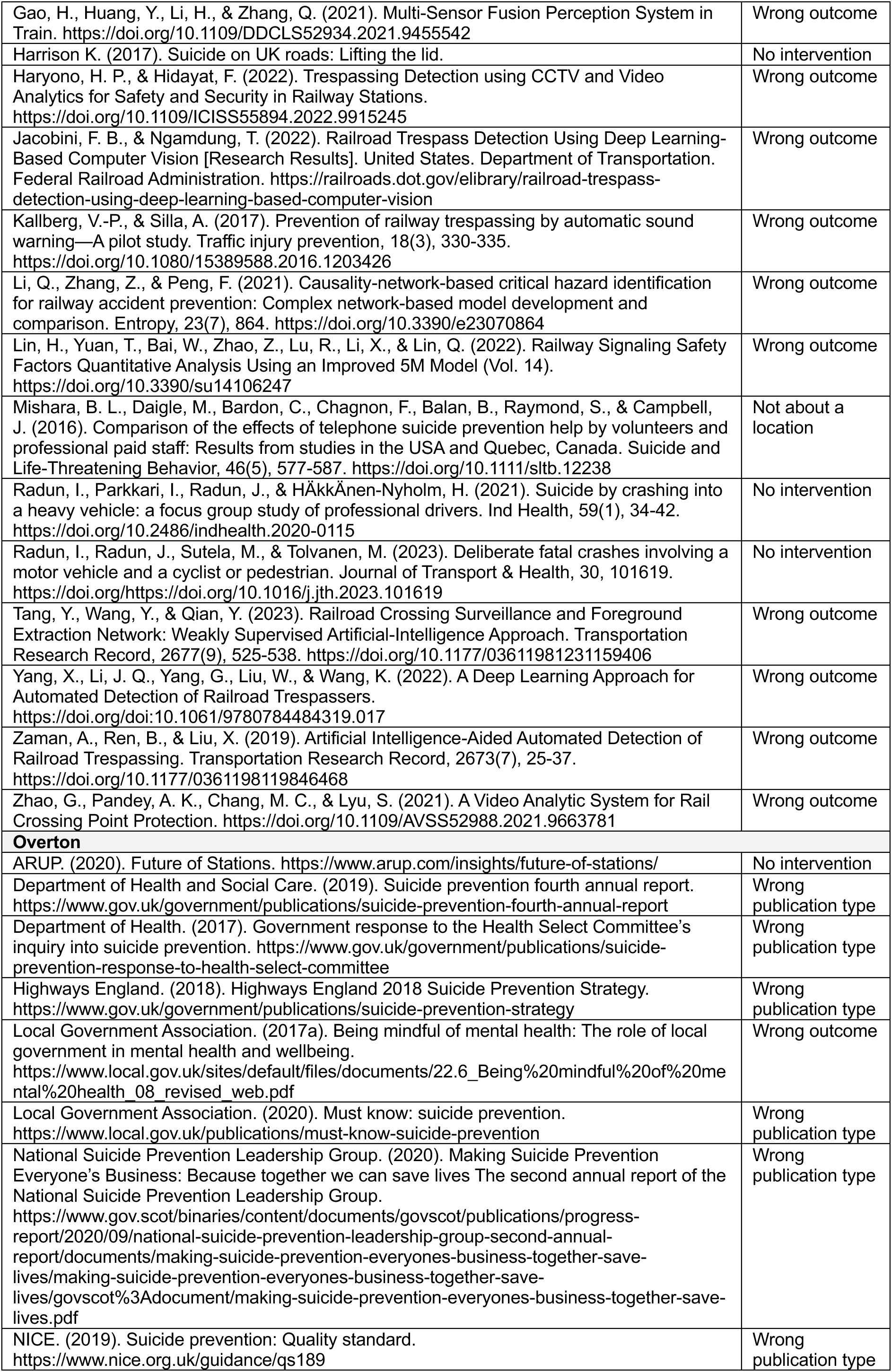
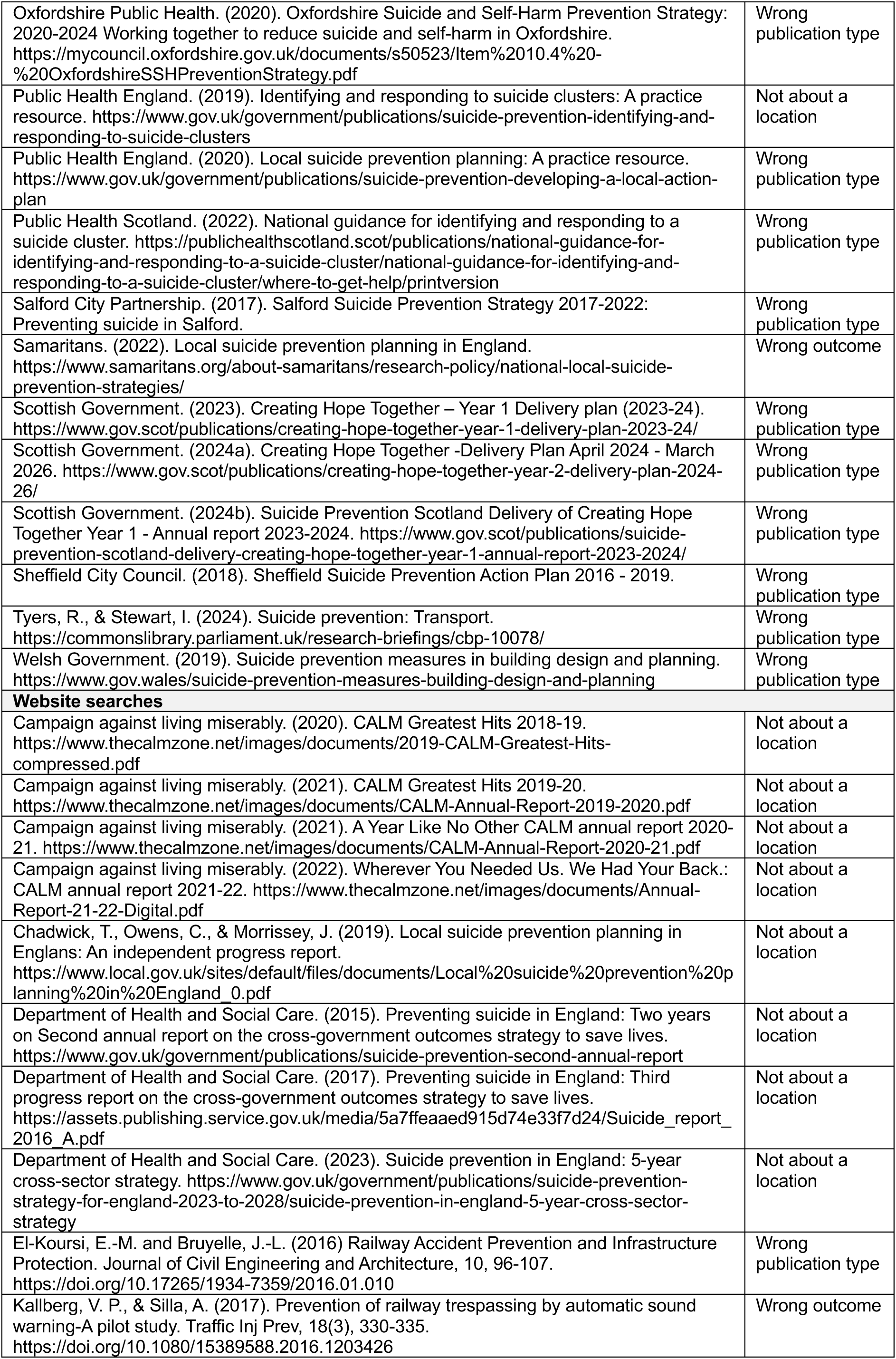
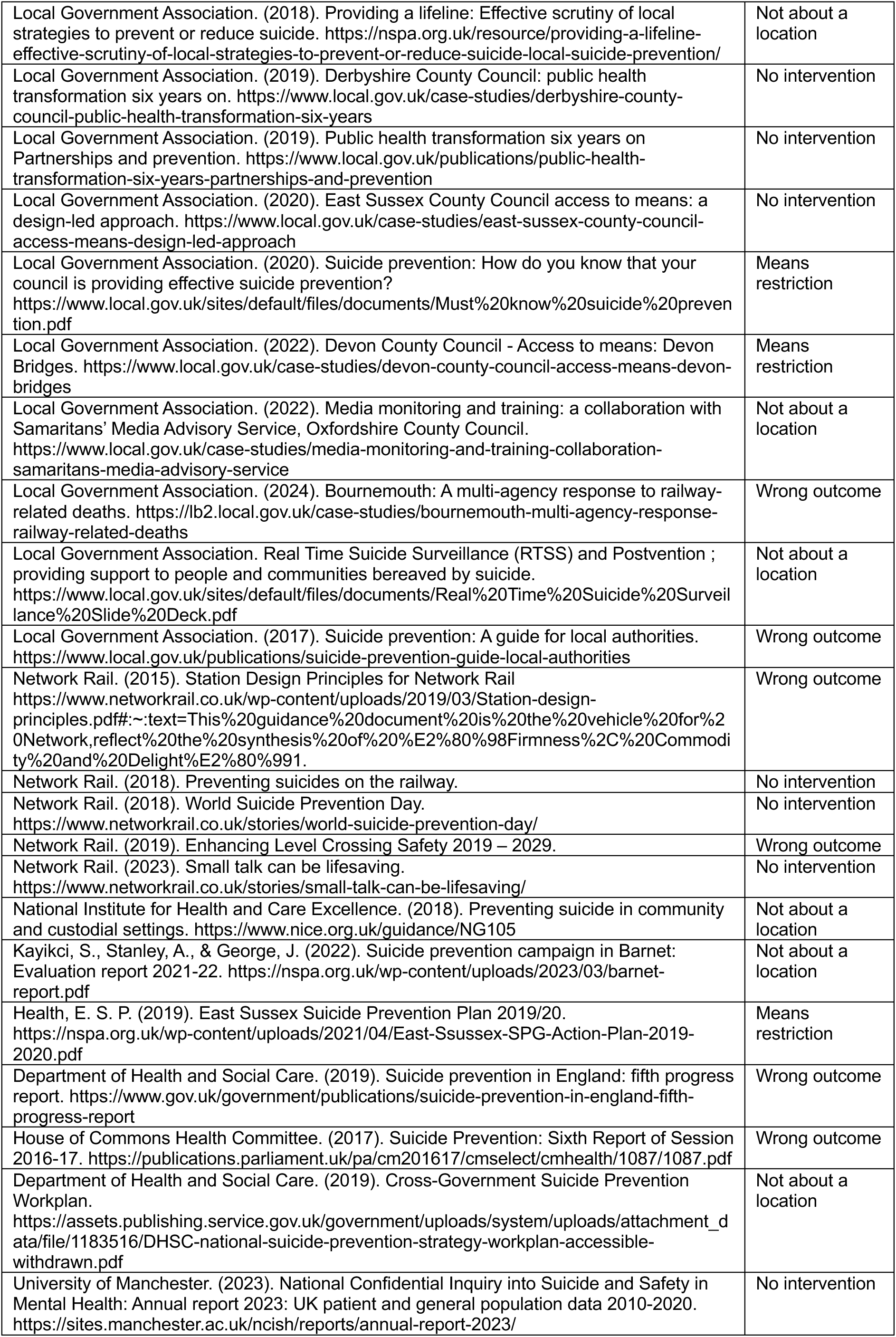
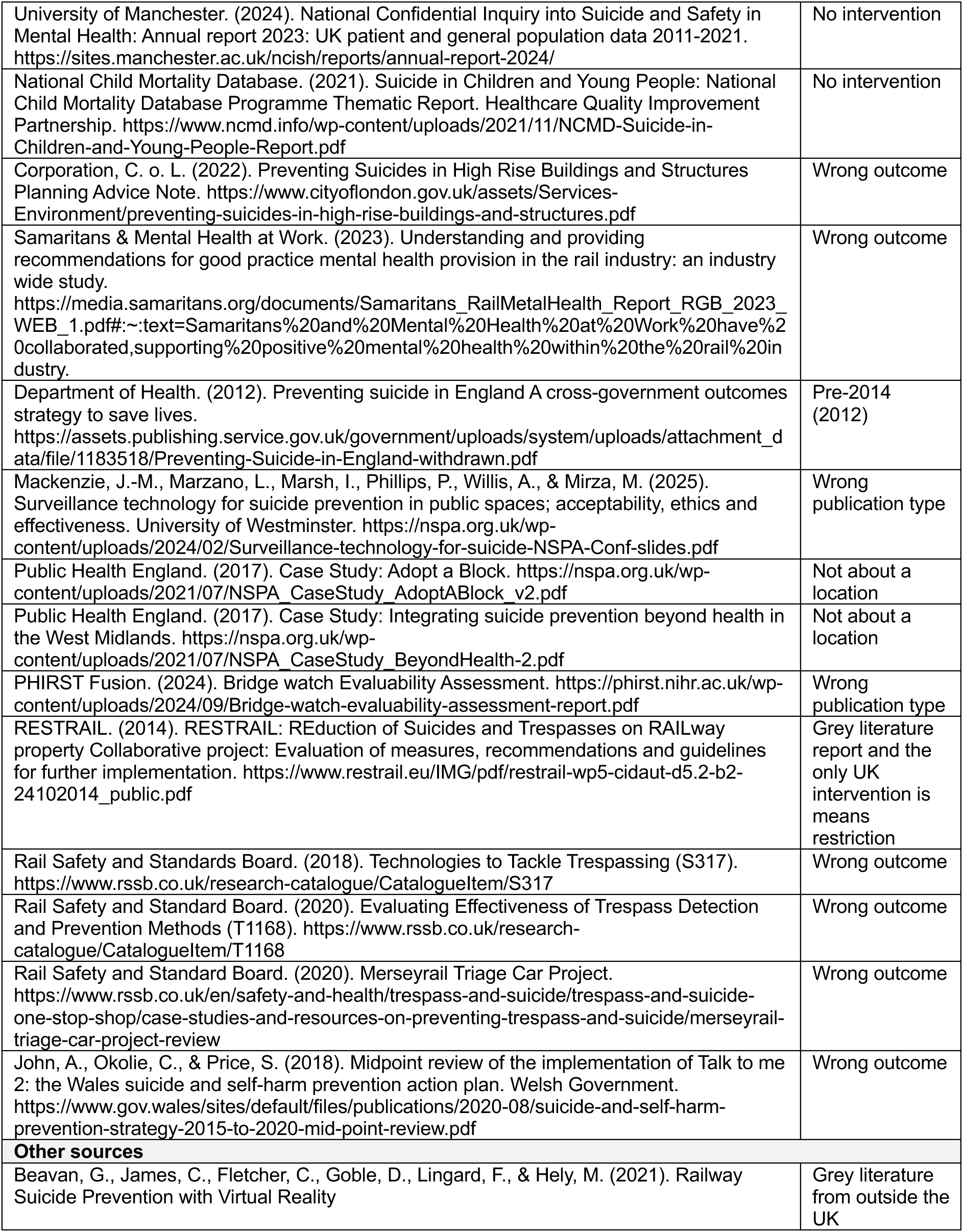

## Footnotes

* This section has been completed by the Centre for Health Economics and Medicines Evaluation (CHEME), Bangor University

1 Figures updated to January 2025 prices from 2022 prices using the Bank of England Inflation calculator (Bank of England. (2025). Inflation calculator. Available at: https://www.bankofengland.co.uk/monetary-policy/inflation/inflationcalculator?number.Sections%5B0%5D.Fields%5B0%5D.Value=482604852&current_year=121.7&comparison_year=135.403 [Accessed 20 March 2025].)

## Notes

### Competing Interest Statement

The authors have declared no competing interest.

## REFERENCES

Bank of England. (2025). Inflation calculator. Available at: https://www.bankofengland.co.uk/monetary-policy/inflation/inflationcalculator?number.Sections%5B0%5D.Fields%5B0%5D.Value=482604852&current_year=121.7&comparison_year=135.403 [Accessed 20 March 2025].

Barker E, Kolves K, De Leo D. (2017). Rail-suicide prevention: Systematic literature review of evidence-based activities. Asia Pac Psychiatry. 9(3). doi: 10.1111/appy.12246

Barker TH, Stone JC, Sears K, et al. (2023). The revised JBI critical appraisal tool for the assessment of risk of bias for randomized controlled trials. JBI Evid Synth. 21(3): 494–506. doi: 10.11124/jbies-22-00430

Chamberlain B, Woodnutt S. (2024). Preventing suicide by jumping in public locations: a systematic review of interventions. Mental Health Practice. 27(3): 24–30. doi: 10.7748/mhp.2024.e1681

Chow S, Men VY, Zaheer R, et al. (2024). Suicide on the Toronto Transit Commission subway system in Canada (1998-2021): a time-series analysis. Lancet regional health. Americas. 34: 100754. doi: 10.1016/j.lana.2024.100754

Erlangsen A, la Cour N, Larsen CO, et al. (2023). Efforts to Prevent Railway Suicides in Denmark. Crisis. 44(2): 169–72. doi: 10.1027/0227-5910/a000827

Giraud C. (2021). Suicide Prevention in the Square Mile: Presentation given at the National Suicide Prevention Annual Conference.

Havârneanu GM, Burkhardt JM, Paran F. (2015). A systematic review of the literature on safety measures to prevent railway suicides and trespassing accidents. Accid Anal Prev. 81: 30–50. doi: 10.1016/j.aap.2015.04.012

Ichikawa M, Inada H, Kumeji M. (2014). Reconsidering the effects of blue-light installation for prevention of railway suicides. Journal of affective disorders. 152: 183–5. doi: 10.1016/j.jad.2013.09.006

Jain N, Wijnen B, Lohumi I, et al. (2024). Economic burden of suicides and suicide attempts in low- and middle-income countries: a systematic review of costing studies. Expert Rev Pharmacoecon Outcomes Res. 24(9): 1067–79. doi: 10.1080/14737167.2024.2388132

Joyner L, Cliffe B, Mackenzie J-M, et al. (2024a). The Use of Smart Surveillance Technologies for Suicide Prevention in PublicSpaces: A Professional Stakeholder Survey from the United Kingdom.

Joyner L, Cliffe B, Mackenzie J-M, et al. (2024b). Summary: The Use of Smart Surveillance Technologies for Suicide Prevention in PublicSpaces: A Professional Stakeholder Survey from the United Kingdom.

Katsampa D, Mackenzie J-M, Crivatu I, et al. (2022). Intervening to prevent suicide at railway locations: findings from a qualitative study with front-line staff and rail commuters. BJPsych open. 8(2): e62. doi: 10.1192/bjo.2022.27

Kiseleva M, Hounsome J, Mann M, et al. (2024). What is the effectiveness of interventions other than physical means restriction to reduce suicide ideation, attempts, and deaths at public locations? Rapid review protocol. OSF. doi: 10.17605/OSF.IO/3XGRA

Kolves K, Leske S, De Leo D. (2023). From suicide surveillance to restricting access to means: A time series study of suicide prevention at the Story Bridge. Aust N Z J Psychiatry. 57(8): 1184–6. doi: 10.1177/00048674231177960

Kupferman S. (2015). How the Bloor Viaduct’s Luminous Veil finally got itself illuminated. Toronto Life. Available at: https://torontolife.com/city/toronto-luminous-veil-bloor-viaduct-lighting/ [Accessed 24th February 2025].

Lee J, Lee CM, Park NK. (2016). Application of sensor network system to prevent suicide from the bridge. Multimedia Tools and Applications. 75(22): 14557–68. doi: doi:10.1007/s11042-015-3134-z

Li X, Onie S, Liang M, et al. (2022). Towards Building a Visual Behaviour Analysis Pipeline for Suicide Detection and Prevention. Sensors (Basel, Switzerland). 22(12). doi: 10.3390/s22124488

Local Government Association. (2022). A new crisis café to prevent suicide at a high-risk location. Available at: https://www.local.gov.uk/case-studies/hertfordshire-county-council-and-stevenage-borough-council-new-crisis-cafe-prevent [Accessed 10th December 2024].

Lockley A, Cheung YTD, Cox G, et al. (2014). Preventing suicide at suicide hotspots: a case study from Australia. Suicide & life-threatening behavior. 44(4): 392–407. doi: 10.1111/sltb.12080

Lockwood C, Munn Z, Porritt K. (2015). Qualitative research synthesis: methodological guidance for systematic reviewers utilizing meta-aggregation. Int J Evid Based Healthc. 13(3): 179–87. doi: 10.1097/XEB.0000000000000062

Matsubayashi T, Sawada Y, Ueda M. (2013). Does the installation of blue lights on train platforms prevent suicide? A before-and-after observational study from Japan. J Affect Disord. 147(1-3): 385–8. doi: 10.1016/j.jad.2012.08.018

Matsubayashi T, Sawada Y, Ueda M. (2014). Does the installation of blue Lights on train platforms shift suicide to another station?: Evidence from Japan. Journal of affective disorders. 169: 57–60. doi: 10.1016/j.jad.2014.07.036

Moola S, Munn Z, Tufanaru C, et al. (2020). Chapter 7: Systematic reviews of etiology and risk. In: Aromataris E & Munn Z (eds.) JBI Manual for Evidence Synthesis. JBI.

National Heart Lung and Blood Institute. (2013). Quality Assessment Tool for Before-After (Pre-Post) studies with no control group. Available at: https://www.nhlbi.nih.gov/health-topics/study-quality-assessment-tools [Accessed 22th January 2025].

National Institute for Health and Care Research Public Health Intervention Responsive Studies Teams. (2024). Bridge Watch Evaluability Assessment, Version 3_1.

Network Rail. (2018). Annual Report and Accounts 2018. Available at: https://www.networkrail.co.uk/wp-content/uploads/2018/07/NRL-2018-ARA-Full.pdf.

Network Rail. (2019). Annual Report and Accounts 2019. Available at: https://www.annualreports.com/HostedData/AnnualReportArchive/n/network-rail_2019.pdf.

Network Rail. (2020). Annual Report and Accounts 2020. Available at: https://www.networkrail.co.uk/wp-content/uploads/2020/07/Annual-report-and-accounts-2020.pdf.

Network Rail. (2024a). Small Talk Saves Lives. Available at: https://www.networkrail.co.uk/communities/safety-in-the-community/railway-safety-campaigns/suicide-prevention-campaigns/ [Accessed 10th January 2025].

Network Rail. (2024b). Suicide prevention on the railway. Available at: https://www.networkrail.co.uk/communities/safety-in-the-community/suicide-prevention-on-the-railway/ [Accessed 10th January 2025].

Ngo NV, Gregor SD, Beavan G, et al. (2022). The Role of Bystanders in the Prevention of Railway Suicides in New South Wales, Australia. Crisis. 43(5): 412–8. doi: 10.1027/0227-5910/a000804

O’Connor RC, Kirtley OJ. (2018). The integrated motivational-volitional model of suicidal behaviour. Philos Trans R Soc Lond B Biol Sci. 373(1754). doi: 10.1098/rstb.2017.0268

O’Neill S, Potts C, Bond R, et al. (2021). An analysis of the impact of suicide prevention messages and memorials on motorway bridges. Suicide & life-threatening behavior. 51(4): 657–64. doi: 10.1111/sltb.12736

Okolie C, Hawton K, Lloyd K, et al. (2020a). Means restriction for the prevention of suicide on roads. Cochrane Database Syst Rev. 9(9): CD013738. doi: 10.1002/14651858.CD013738

Okolie C, Wood S, Hawton K, et al. (2020b). Means restriction for the prevention of suicide by jumping. Cochrane Database Syst Rev. 2(2): CD013543. doi: 10.1002/14651858.CD013543

Onie S, Li X, Glastonbury K, et al. (2023). Understanding and detecting behaviours prior to a suicide attempt: A mixed-methods study. The Australian and New Zealand journal of psychiatry. 57(7): 1016–22. doi: 10.1177/00048674231152159

Owens C, Derges J, Abraham C. (2019). Intervening to prevent a suicide in a public place: a qualitative study of effective interventions by lay people. BMJ open. 9(11): e032319. doi: 10.1136/bmjopen-2019-032319

Pirkis J, Too LS, Spittal MJ, et al. (2015). Interventions to reduce suicides at suicide hotspots: a systematic review and meta-analysis. Lancet Psychiatry. 2(11): 994–1001. doi: 10.1016/S2215-0366(15)00266-7

Prosser I. (2022). Pulling together for better mental health in the rail industry. Office of Rail and Road. Available at: https://www.orr.gov.uk/search-news/pulling-together-better-mental-health-rail-industry [Accessed 20 March 2025].

Public Health England. (2015). Preventing suicides in public places: A practice resource, Public Health England.

Public Health Scotland. (2022a). National guidance on action to address suicides at locations of concern. Available at: https://publichealthscotland.scot/publications/national-guidance-on-action-to-address-suicides-at-locations-of-concern/national-guidance-on-action-to-address-suicides-at-locations-of-concern/where-to-get-help/ [Accessed 16th September 2024].

Public Health Scotland. (2022b). Rapid literature review in reducing suicides at locations of concern.

Public Health Wales. (2024). Deaths by suspected suicide 2022-2023.

Rail Safety and Standard Board. (2020). Trespass & Welfare Officers – Network Rail & South Western Railway joint project Wessex Route. Available at: https://www.rssb.co.uk/en/safety-and-health/trespass-and-suicide/trespass-and-suicide-one-stop-shop/case-studies-and-resources-on-preventing-trespass-and-suicide/trespass-and-welfare-officers--nr-and-swr-joint-project-wessex-route [Accessed 10th January 2025].

Ross V, Koo YW, Kolves K. (2020). A suicide prevention initiative at a jumping site: A mixed-methods evaluation. EClinicalMedicine. 19: 100265. doi: 10.1016/j.eclinm.2020.100265

Samaritans. (2024a). The economic cost of suicide in the UK. Samaritans. (2024b). Guidance for reporting on suicide in public places.

Schünemann HJ, Brennan S, Akl EA, et al. (2023). The development methods of official GRADE articles and requirements for claiming the use of GRADE - A statement by the GRADE guidance group. J Clin Epidemiol. 159: 79–84. doi: 10.1016/j.jclinepi.2023.05.010

Shin S, Pirkis J, Clapperton A, et al. (2024a). Effectiveness of partial restriction of access to means in jumping suicide: lessons from four bridges in three countries. Epidemiology and psychiatric sciences. 33: e38. doi: 10.1017/S2045796024000428

Shin S, Pirkis J, Spittal MJ, et al. (2024b). Change in incidents of suicidal acts after intervention on a bridge in South Korea. Social psychiatry and psychiatric epidemiology. doi: 10.1007/s00127-024-02744-9

Sinyor M, Men VY, Chan PPM, et al. (2024). Long-Term Impact of the Bloor Viaduct Suicide Barrier on Suicides in Toronto: A Time-Series Analysis: Effet à long terme de la barrière anti-suicide du viaduc Bloor sur les suicides à Toronto : une analyse chronologique. Canadian Journal of Psychiatry. doi: 10.1177/07067437241293978

Stack S. (2015). Crisis Phones - Suicide Prevention Versus Suggestion/Contagion Effects. Crisis. 36(3): 220–4. doi: 10.1027/0227-5910/a000313

Too LS, Ross A, Pirkis J, et al. (2020). The Impact of the “Pause. Call. Be Heard” Campaign on Help-Seeking and Suicidal Behaviors Within Rail Environment in Victoria, Australia. Suicide & life-threatening behavior. 50(2): 490–501. doi: 10.1111/sltb.12604

Too LS, Spittal MJ, Bugeja L, et al. (2015). An investigation of neighborhood-level social, economic and physical factors for railway suicide in Victoria, Australia. Journal of affective disorders. 183: 142–8. doi: 10.1016/j.jad.2015.05.006

Torok M, Passioura J, Konings P, et al. (2023). A Spatial Analysis of Suicide Displacement at a High-Risk Cliff-Based Location Following Installation of a Means Restriction Initiative. Prevention science : the official journal of the Society for Prevention Research. 24(7): 1292–301. doi: 10.1007/s11121-023-01504-6

Waalen A, Bera K, Bera R. (2020). Suicide hotspots, interventions, and future areas of work at a Californian university. Death studies. 44(9): 569–77. doi: 10.1080/07481187.2019.1595221

World Health Organization. (2014). Preventing suicide: A global imperative, Luxembourg.

Yogesan DS, Onie S, De Belen RA, et al. (2023). Evaluating a Human Detection Model in a Behaviour Analysis Pipeline for Suicide Prevention. Annu Int Conf IEEE Eng Med Biol Soc. 2023: 1–4. doi: 10.1109/EMBC40787.2023.10339992

Zalsman G, Hawton K, Wasserman D, et al. (2016). Suicide prevention strategies revisited: 10-year systematic review. The lancet. Psychiatry. 3(7): 646–59. doi: 10.1016/S2215-0366(16)30030-X

